# A year of Covid-19 GWAS results from the GRASP portal reveals potential SARS-CoV-2 modifiers

**DOI:** 10.1101/2021.06.08.21258507

**Authors:** Florian Thibord, Melissa V. Chan, Ming-Huei Chen, Andrew D. Johnson

## Abstract

Host genetic variants influence the susceptibility and severity of several infectious diseases, and the discovery of novel genetic associations with Covid-19 phenotypes could help developing new therapeutic strategies to reduce its burden.

Between May 2020 and June 2021, we used Covid-19 data released periodically by UK Biobank and performed 65 Genome-Wide Association Studies (GWAS) in up to 18 releases of Covid-19 susceptibility (N=18,481 cases in June 2021), hospitalization (N=3,260), severe outcomes (N=1,244) and death (N=1,104), stratified by sex and ancestry.

In coherence with previous studies, we observed 2 independent signals at the chr3p21.31 locus (rs73062389-A, OR=1.21, P=4.26×10^−15^ and rs71325088-C, OR=1.62, P=2.25×10^−9^) modulating susceptibility and severity, respectively, and a signal influencing susceptibility at the *ABO* locus (rs9411378-A, OR=1.10, P=3.30×10^−12^), suggesting an increased risk of infection in non-O blood groups carriers. Additional signals at the *APOE* (associated with severity and death) *LRMDA* (susceptibility in non-European) and chr2q32.3 (susceptibility in women) loci were also identified but did not replicate in independent datasets. We then devised an original approach to extract variants exhibiting an increase in significance over time. When applied to the susceptibility, hospitalization and severity analyses, this approach revealed the known *DPP9, RPL24* and *MAPT* loci, amongst thousands of other signals. Finally, this significance trajectory analysis was applied to the larger Covid-19hgi meta-analyses, where additional loci of interest, related to the immune system, were identified.

These results, freely available on the GRASP portal, provide new insights on the genetic mechanisms involved in Covid-19 phenotypes.

## Introduction

The severe acute respiratory syndrome – coronavirus 2 (SARS-CoV-2) is responsible for the coronavirus disease 2019 (Covid-19) which affects individuals with variable severity, ranging from asymptomatic patients to mild respiratory symptoms, hypercytokinemia, pneumonia, thrombosis and even death.^1,2^ Understanding the mechanisms leading to heterogeneous symptoms and susceptibility is essential in order to develop efficient treatments and improve patient care. Host genetic diversity has been shown to influence the effects of infection to several viruses,^3^ such as variations in CCR5 [MIM: 601373] leading to HIV resistance^4^ [MIM: 609423] or *IRF7* [MIM: 605047] deficiency affecting *Influenza susceptibility* [MIM: 614680].^5^

In order to discover human genetic determinants to Covid-19 susceptibility and severity, several biobanks and research groups worldwide collaborated to perform Genome-Wide Association Studies (GWAS) and meta-analyses of the GWAS. In June 2020, a study involving 1,980 Covid-19 infected patients with respiratory failure was the first to reveal genome-wide significant (P < 5 × 10^−8^) associations at the 3p21.31 locus, encompassing *SLC6A20* [MIM: 605616] and several chemokine receptors, and at the *ABO* [MIM: 110300] locus on chromosome 9.^6^ These 2 signals were later validated in independent analyses for both Covid-19 susceptibility and severity^7,8^ while additional significant associations were observed at loci involved in immune response or inflammation, such as *IFNAR2* [MIM: 602376], *DPP9* [MIM: 608258], *TYK2* [MIM: 176941], *CCHCR1* [MIM: 605310] and *OAS1* [MIM: 164350]. Notably, these findings implicate common and uncommon variants, while studies trying to identify associations of rare variants have been unsuccessful so far.^9^ The largest effort is currently led by the Covid-19 host genetics initiative (Covid-19hgi),^10^ which completed meta-analyses of results shared by 61 studies as of June 15^th^ 2021, and plan to release new results as additional data is made available. A major contributor to this group is the UK Biobank (UKB)^11^ which periodically releases the results of Covid-19 tests and related deaths, as well as health care data for its nearly 500,000 consented participants, to approved researchers.

We created a public Covid-19 GWAS results portal (https://grasp.nhlbi.nih.gov/Covid19GWASResults.aspx) in order to provide rapid deep annotation for emerging genetics results and facilitate open access to the scientific community. We contribute to this resource by performing GWAS on each Covid-19 data release from the UK Biobank, including sex-specific, ancestry-specific, and trans-ethnic Covid-19 related GWAS, along with a deep set of annotations for top variants (with P < 1 × 10^−5^). For each release, up to 65 GWAS have been generated including Covid-19 susceptibility, Covid-19 hospitalization, severe Covid-19 with respiratory failure, and Covid-19 death. Here we describe the results of these GWAS, 612 in total as of June 18^th^ 2021, and report the evolution of signals associated with these Covid-19 phenotypes over the consecutive datasets released by UKB since May 2020. The latter approach, tracking the evolution of genetic signatures iteratively in UKB, suggests a valuable new analytic approach in genetic biobank studies where emerging true signals may be identified before they reach genome-wide significance based on their trajectories of significance.

## Materials and Methods

### UKB data

Analyses are based on v.3 of the UKB imputed dataset,^12^ which provide genomic data for 487,320 participants from multiple ethnicities, including 459,250 of European ancestry (EUR), 7,644 of African ancestry (AFR), 9,417 of South Asian ancestry (SAS) and 11,009 of other ancestries (OTHERS). Participants were enrolled at ages ranging from 37 to 73 and are 51.16% female. UKB started to release Covid-19 test results of its participants on March 15^th^ 2020, and periodically update this resource as new cases are reported. Furthermore, information about Covid-19 related death was made available from June 2020, while inpatient data and primary care data was first added during the summer of 2020 and are periodically updated. Details regarding the definition and selection of cases with Covid-19 susceptibility, Covid-19 hospitalization, Covid-19 severity, and Covid-19 death are available in **Table S1**.

Depending on the Covid-19 phenotype analyzed (susceptibility, hospitalization, severity, or death), up to 3 different subsets of participants were used as controls. For Covid-19 susceptibility, cases with positive test results were analyzed against either participants tested with negative results (labelled Tested), or against all participants without a positive test (labelled Population). For analyses of Covid-19 hospitalization, patients hospitalized due to Covid-19 were tested against non-hospitalized participants with a positive test (Positive), or non-hospitalized participants with a test (Tested), or against all non-hospitalized participants (Population). For analyses of Covid-19 severity, patients requiring invasive respiratory support or patients who died from complications were tested against non-severe participants with a positive test (Positive), or non-severe participants with a test (Tested) or all non-severe participants (Population). For analyses of Covid-19 death, patients with Covid-19 death were tested against participants with a positive test (Positive), or participants with a test (Tested) or against all participants (Population).

### Analyses

Each GWAS was conducted with SAIGE v0.38,^13^ which controls for population stratification, relatedness and case-control imbalance, and adjusted for baseline age (at enrollment), sex and 10 genetic principal components. For the results uploaded to the GRASP portal, variants were filtered on imputation quality (r^2^ > 0.3), minor allele count (MAC > 2), and minor allele frequency (MAF > 0.0001). However, for the results presented in this manuscript, we applied a more stringent filter, and considered only well-imputed variants (r^2^ > 0.8) and common variants (MAF > 0.01). After applying this filter, the lambda (genomic control factor) ranged from 0.988 to 1.027 in all 65 analyses of the last data release analyzed (**Table S2**), indicating no systematic inflation. For analyses prior to the June 18^th^ 2020 release, we conducted analyses on participants of European ancestry only, and started adding new analyses stratified by sex and ancestry from June 18^th^ 2020 onward. GWAS were stratified for EUR, AFR, SAS, and OTHERS ancestries, and an additional trans-ancestry GWAS combining all participants (labelled ALL) as well as GWAS combining non-European (nEUR) participants were performed.

Associations are either reported as odd ratios (OR) and 95% confidence intervals or as beta coefficients (β) and associated standard errors (SE). Linkage disequilibrium (LD) was estimated by squared correlation (r^2^) using UKB EUR imputed data. Given the large number of variants that may be significantly (or suggestively) associated at a locus, we assigned significantly (or suggestively) associated variants to a common locus if they were separated by less than 1Mb, and only reported the lead variant at that locus. To test 2 observed effects are equal, we used the Z statistic: 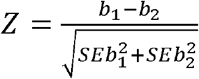, with *b*_1_ and *b*_2_ corresponding to the observed effects and SEb_1_ and SEb_2_ the associated standard errors. In addition, haplotype analyses were performed with the LDlink LDhap tool,^14^ while regional association plots were generated with locuszoom.^15^

### Annotation

For each analysis hosted on the portal, we provide comprehensive annotation for top results (P < 1 × 10^−5^) using ANNOVAR^16^ and the RESTful API service provided by CADD v1.6.^17^ We also retrieve known phenotype associations extracted from the GRASP^18^ and EBI GWAS catalogs,^19^ and known eQTLs extracted from GTeX v8^20^ and other eQTL resources compiled from nearly 150 datasets (built upon the work of Zhang *et al*,^21^ detailed in **Table S3**).

### Significance Trajectory Analyses

All 65 analyses were updated as soon as new data was released from UKB. As a result, we obtained results for these analyses at different time points, which differed by the addition of new cases, thus increasing power in most recent analyses. With an increase in power, *bona fide* signals associated with Covid-19 phenotypes should increase in significance in each consecutive data release analyzed. We designed a workflow to extract these signals with positive significance trajectory in each analysis: for each variant (i) in the most recent data release analyzed, P < 10^−4^; (ii) in the first data release analyzed, P must be greater than in the most recent data release analyzed; (iii) P cannot decrease in significance by more than one order of magnitude between 2 consecutive releases; (iv) P must have increased in significance by more than 1 order of magnitude at least once between 2 consecutive releases; (v) the latest P observed must have decreased by more than 1 order of magnitude since the minimal P observed. This set of rules should ensure to extract variants which increased in significance since we started performing these analyses, while allowing some stagnation, which might happen due to random sampling and/or low case increase between 2 consecutive releases.

### The Covid-19hgi datasets

For replication purposes and significance trajectory analyses, we used the publicly available Covid-19hgi meta-analyses summary statistics (freeze 6) for Covid-19 susceptibility (labelled C2, N = 112,612 cases), hospitalization (B2, N = 24,274) and severity (A2, N = 8,779 cases). These datasets are currently the largest analyses of Covid-19 phenotypes available. Publicly available summary statistics do not include the 23andMe dataset, and a version of the summary statistics without UKB was also made available for the B2 and C2 analyses. As there was no overlap of samples with our analyses, we used these summary statistics to replicate our novel findings from susceptibility and hospitalization analyses, as well as our suggestive findings from the significance trajectory analyses. To replicate findings from our severe analyses, we used the A2 summary statistics without UKB from the freeze 5 (the A2 results without UKB from freeze 6 were not available). In addition, we retrieved Covid-19hgi summary statistics (with UKB) from all publicly available freezes to perform significance trajectory analyses with the Covid-19hgi datasets (freezes 2 to 6 for B2 and C2; freezes 3 to 6 for A2).

### Data Availability

Permission was obtained to post UKB summary statistics under an approved application (ID 28525). The association results are available on the portal, as well as annotated top results. In addition to UKB summary statistics, results from other efforts are also hosted on the portal. Authors of Covid-19 GWAS publications have been contacted to seek approval before hosting the results of their analyses on the GRASP Covid-19 portal. Summary statistics at this time include multiple releases of the Covid-19hgi group, severe hospitalization results from Ellinghaus *et al*^6^ and the GenOMICC study,^7^ as well as hospitalization status and time to end Covid-19 symptoms from the COLCORONA study,^22^ with all top results being re-annotated in the common framework mentioned above.

## Results

### UKB Covid-19 demographics

Using the latest UKB data releases available at this point, with susceptibility and hospitalization phenotypes analyzed on June 18^th^ 2021, and severe Covid-19 and death analyzed on May 9^th^ 2021, we retrieved 86,435 participants with a Covid-19 diagnostic, of which 18,481 tested positive. According to inpatient care data, 3,260 positive cases were hospitalized, while 1,244 patients with severe Covid-19 diagnostic received respiratory support and/or died from complications (**Table 1**). Since May 7^th^ 2020, we analyzed up to 18 UKB data releases regarding Covid-19 susceptibility, 8 data releases concerning Covid-19 related deaths, 6 for Covid-19 hospitalization and 5 for Covid-19 severity. Amongst Covid-19 positive participants, we observed a global increase in the percentage of female cases, starting at 45.3% at the first release analyzed, and reaching 52.8% in the last, while men were more likely to be infected (P = 8.06 × 10^−6^), hospitalized (P = 1.59 × 10^−45^), or develop severe complications (P = 1.51 × 10^−34^) and die from Covid-19 (P = 2.12 × 10^−27^) (**Table S4**). There was also a decrease in the mean age of positive cases, ranging from 57.02 to 53.57, with a significant drop after the 2020 summer (**Table 2**), with younger individuals more likely to be infected (P < 10^−300^) while increase in age was associated with hospitalization (P = 4.54 × 10^−90^), severity (P = 5.63 × 10^−108^) and death (P = 1.11 < 10^−122^) (**Table S4**). Positive cases were mainly of European ancestry, representing 85.5% of all Covid-19 positive participants in the first analysis and growing to 89.6% in the last. South Asian ancestry, African ancestry, and other ancestry represent 4.4%, 3% and 3% of positive cases, respectively, in this UKB data release. However, compared to European participants, non-European participants were enriched amongst cases (P = 3.09 × 10^−33^ for AFR, P = 1.10 × 10^−91^ for SAS, P = 1.96 × 10^−6^ for OTHERS), as well as hospitalized (P = 2.05 × 10^−53^ for AFR, P = 1.89 × 10^−24^ for SAS, P = 6.69 × 10^−9^ for OTHERS), severe cases (P = 1.02 × 10^−17^ for AFR, P = 5.93 × 10^−11^ for SAS, P = 0.004 for OTHERS) and deceased participants (P = 2.17 × 10^−14^ for AFR, P = 6.14 × 10^−9^ for SAS, P = 0.009 for OTHERS) (**Table S4**).

**Table 1.**
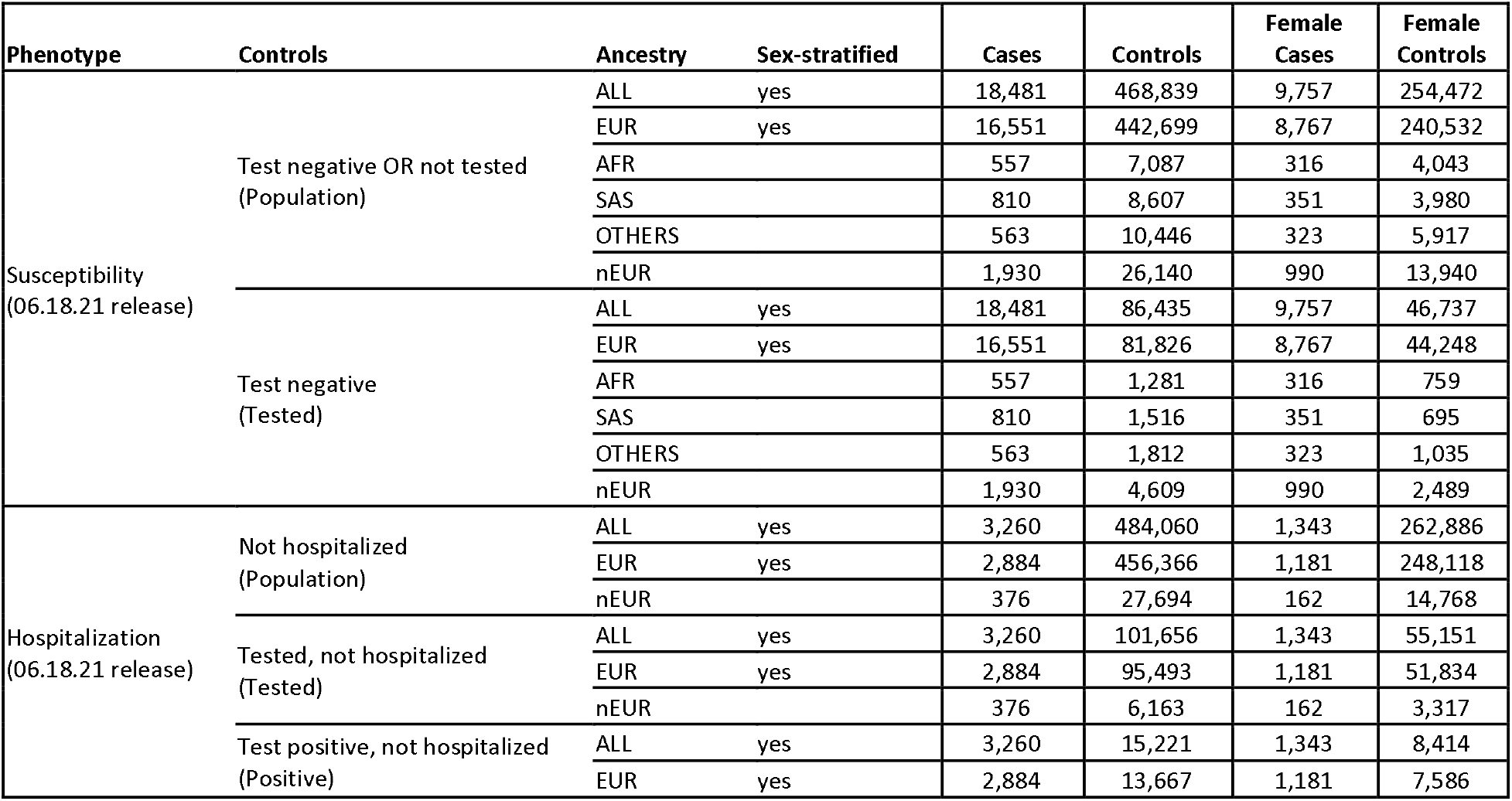
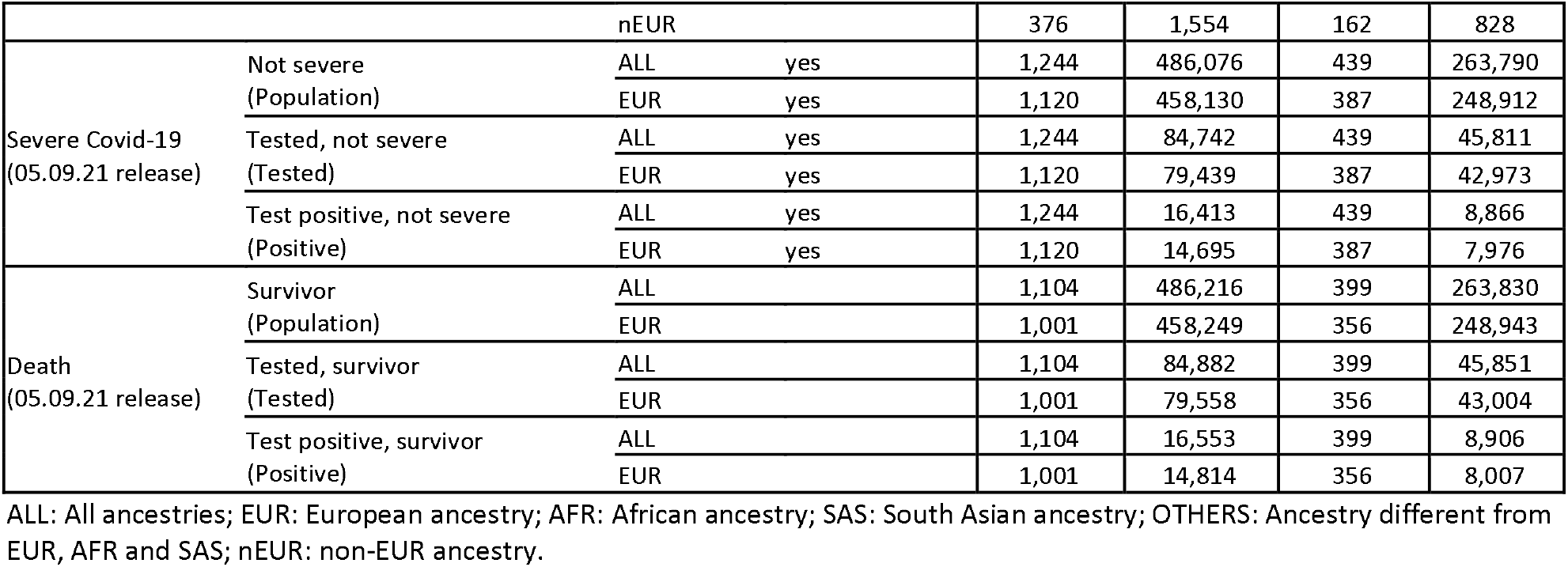
Sample sizes for each analysis, as of 06.18.21

**Table 2.** Composition of the consecutive UK Biobank Covid-19 data releases

| Release Date | Cases (tested positive) | Controls | % Tested Positive Females | Age Cases<sup>a</sup> |
| --- | --- | --- | --- | --- |
| 05.07.20<sup>b</sup> | 1,029 | 486,291 | 45.29 | 57.02 (9.12) |
| 05.25.20<sup>b</sup> | 1,270 | 486,050 | 46.93 | 56.57 (9.25) |
| 06.05.20 | 1,412 | 485,908 | 47.31 | 56.45 (9.23) |
| 06.18.20 | 1,485 | 485,835 | 47.21 | 56.47 (9.19) |
| 07.14.20 | 1,670 | 485,650 | 46.59 | 56.89 (9.14) |
| 08.04.20 | 1,723 | 485,597 | 46.66 | 56.77 (9.11) |
| 09.08.20 | 1,821 | 485,499 | 46.73 | 56.66 (9.10) |
| 10.16.20 | 3,050 | 484,270 | 48.69 | 55.19 (9.09) |
| 11.03.20 | 4,372 | 482,948 | 49.38 | 54.79 (8.95) |
| 11.25.20 | 5,868 | 481,452 | 49.52 | 54.56 (8.88) |
| 12.04.20 | 7,435 | 479,885 | 50.46 | 54.36 (8.80) |
| 01.04.21 | 8,722 | 478,598 | 50.50 | 54.28 (8.79) |
| 01.22.21 | 13,401 | 473,919 | 52.27 | 53.85 (8.66) |
| 02.02.21 | 14,802 | 472,518 | 52.44 | 53.81 (8.64) |
| 02.24.21 | 15,738 | 471,582 | 52.47 | 53.77 (8.63) |
| 04.04.21 | 16,586 | 470,734 | 52.68 | 53.70 (8.62) |
| 05.09.21 | 17,657 | 469,663 | 52.70 | 53.64 (8.62) |
| 06.18.21 | 18,481 | 468,839 | 52.79 | 53.57 (8.60) |

<sup>a</sup> Mean baseline Age (standard deviation), at recruitment
<sup>b</sup> For the first 2 releases, the numbers reflect all ancestries, but only cases and controls of European ancestry were considered for analyses

### Genome-wide significant results

Between May 2020 and June 2021, we observed several genome-wide significant (P < 5 × 10^−8^) signals across the 65 analyses. However, many signals were only punctually significant, and only a handful of signals remained significant in subsequent data releases analyzed. For instance, the analysis of Covid-19 susceptibility in participants of European ancestry, with untested or negatively tested participants as controls (noted Population controls), revealed 8 signals reaching genome-wide significance at some point (**Figure 1**), of which only 2 remained significant in the last data release analyzed (on 06.18.21): the chr3p21.31 locus encompassing SLC6A20 and several chemokine receptors (rs73062389-A, MAF = 0.06, OR = 1.22 [1.16; 1.28], P = 7.60 × 10^−16^), and the *ABO* locus on chromosome 9 (rs9411378-A, MAF = 0.22, OR = 1.10 [1.07; 1.14], P = 8.78 × 10^−12^). These two loci were previously reported to modulate Covid-19 susceptibility in several studies.^6–8,10^

**Figure 1.**
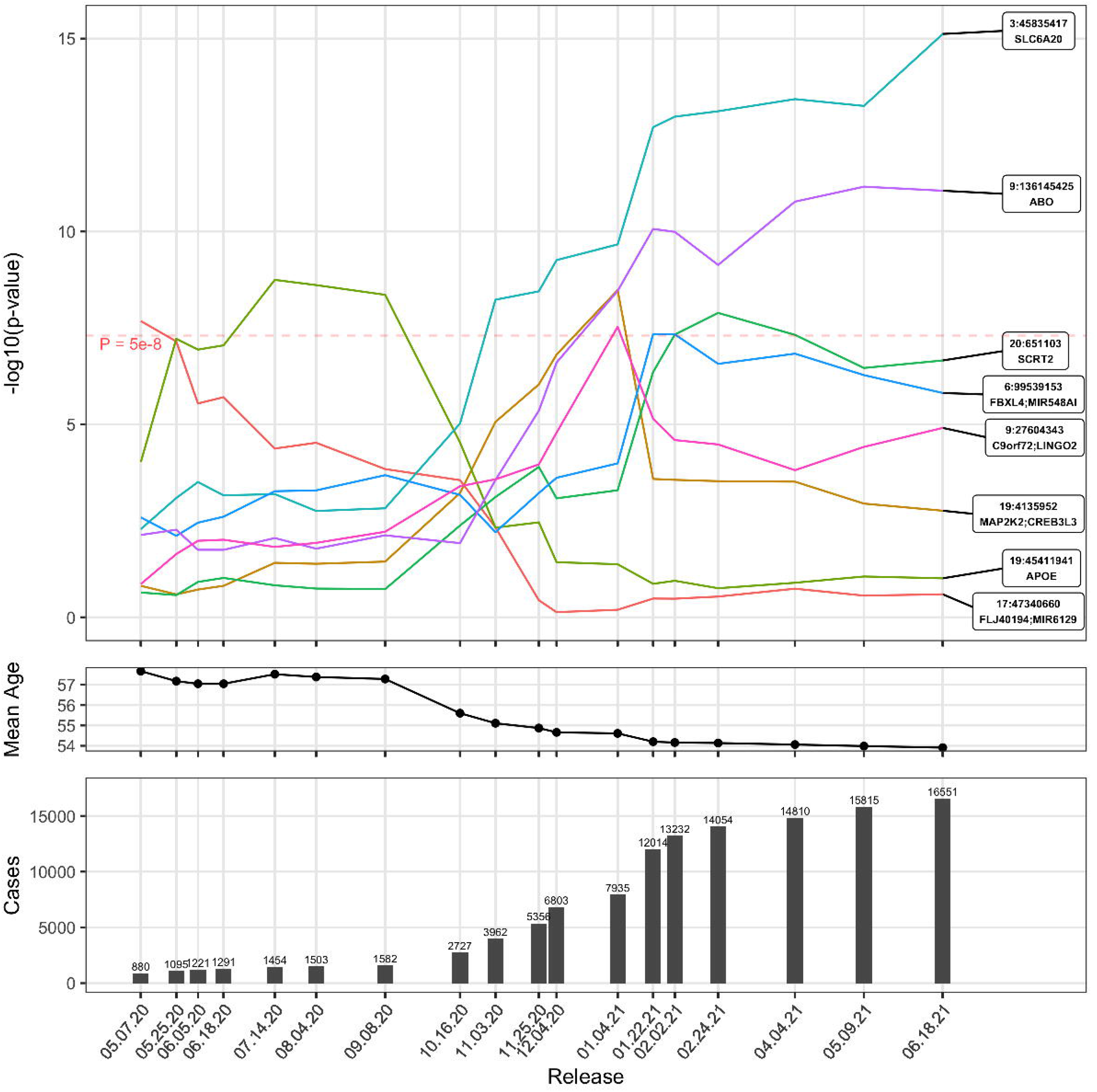
Evolution of significant signals associated with covid-19 susceptibility in UKB participants of European ancestry, using Population as controls. The top panel represents the evolution in significance of signals reaching genome wide significance at least once across all data releases analyzed in Europeans for Covid-19 susceptibility. The middle panel represent the mean age of cases in each data release. The bottom panel represent the number of cases in each data release. A representation of these signals across the Covid-19hgi releases is available as Figure S50.

Across all 65 analyses, 5 loci reached genome-wide significance in the last data releases analyzed (on 06.18.21 for susceptibility and hospitalization, and 05.09.21 for severity and death): the chr3p21.31 locus, *ABO, APOE* [MIM: 107741], *LRMDA* [MIM: 614537] and an intergenic signal at the chr2q32.3 locus (**Table 3**). All associations with P < 10^−5^, from all 65 analyses, are available in **Table S5**. In addition, all signals reaching genome-wide significance in any data release analyzed are presented in **Figures S1-S46**. The signals reported in the following correspond to results using Population controls, but after systematically applying the Z-test for the equality of regression coefficients we did not observe a significant difference of effect when using a different set of control, such as Tested controls (P > 0.05, **Figure S47**).

**Table 3.**
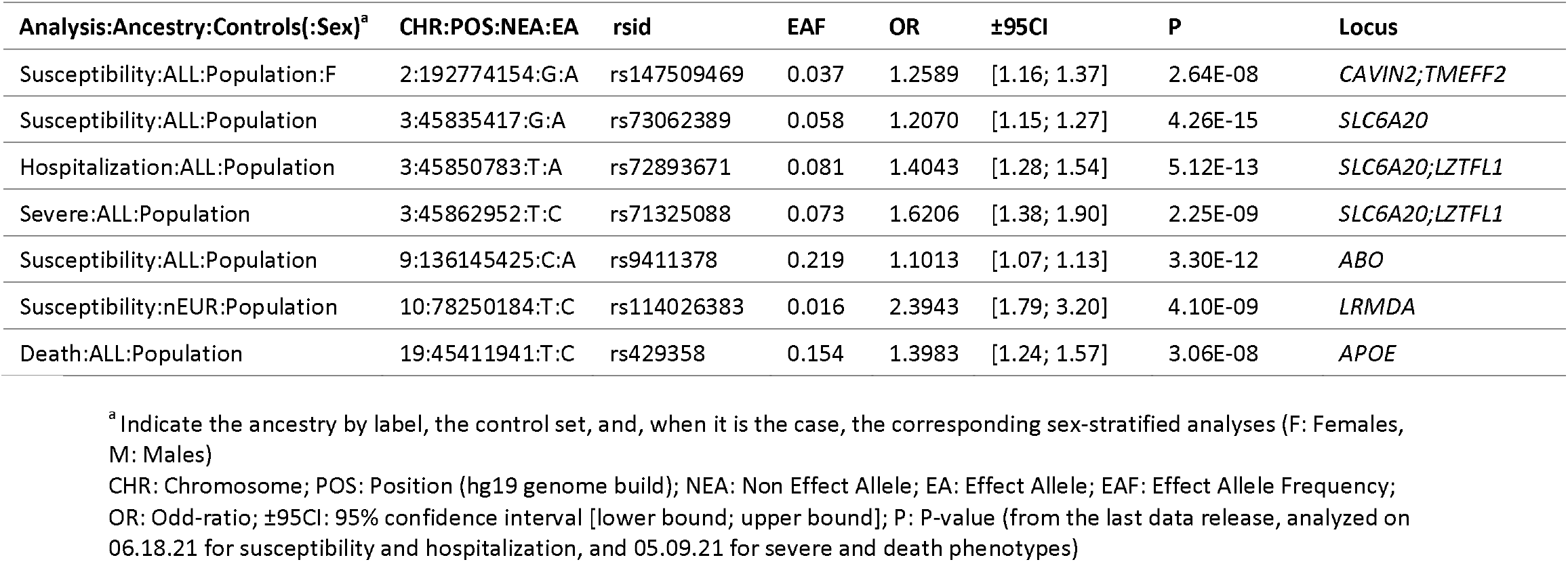
Lead variants associated with Covid-19 phenotypes

| Analysis:Ancestry:Controls(:Sex)<sup>a</sup> | CHR:POS:NEA:EA | rsid | EAF | OR | ±95CI | P | Locus |
| --- | --- | --- | --- | --- | --- | --- | --- |
| Susceptibility:ALL:Population:F | 2:192774154:G:A | rs147509469 | 0.037 | 1.2589 | [1.16; 1.37] | 2.64E-08 | *CAVIN2;TMEFF2* |
| Susceptibility:ALL:Population | 3:45835417:G:A | rs73062389 | 0.058 | 1.2070 | [1.15; 1.27] | 4.26E-15 | *SLC6A20* |
| Hospitalization:ALL:Population | 3:45850783:T:A | rs72893671 | 0.081 | 1.4043 | [1.28; 1.54] | 5.12E-13 | *SLC6A20;LZTFL1* |
| Severe:ALL:Population | 3:45862952:T:C | rs71325088 | 0.073 | 1.6206 | [1.38; 1.90] | 2.25E-09 | *SLC6A20;LZTFL1* |
| Susceptibility:ALL:Population | 9:136145425:C:A | rs9411378 | 0.219 | 1.1013 | [1.07; 1.13] | 3.30E-12 | *ABO* |
| Susceptibility:nEUR:Population | 10:78250184:T:C | rs114026383 | 0.016 | 2.3943 | [1.79; 3.20] | 4.10E-09 | *LRMDA* |
| Death:ALL:Population | 19:45411941:T:C | rs429358 | 0.154 | 1.3983 | [1.24; 1.57] | 3.06E-08 | *APOE* |

<sup>a</sup> Indicate the ancestry by label, the control set, and, when it is the case, the corresponding sex-stratified analyses (F: Females, M: Males)
CHR: Chromosome; POS: Position (hg19 genome build); NEA: Non Effect Allele; EA: Effect Allele; EAF: Effect Allele Frequency; OR: Odd-ratio; ±95CI: 95% confidence interval [lower bound; upper bound]; P: P-value (from the last data release, analyzed on 06.18.21 for susceptibility and hospitalization, and 05.09.21 for severe and death phenotypes)

#### The 3p21.31 locus

This locus was associated with Covid-19 susceptibility, hospitalization and severity, in EUR and ALL ancestries, regardless of the set of controls employed. However, as mentioned in previous works,^8,10^ the lead variant associated with susceptibility was not in LD with the lead variants for hospitalization nor severity (r2 < 0.01) suggesting 2 distinct signals modulating Covid-19 susceptibility and severity (**Figure S48**).

#### ABO

The *ABO* locus was associated with Covid-19 susceptibility for ALL and EUR participants, using either Population or Tested controls. Using haplotype analyses with blood group tagging variants, we determined the blood groups tagged by the lead variant associated with susceptibility (rs9411378). For the 5 main blood groups, we used the following tagging variants^23^: rs41302905 (tagging for O2), rs8176743 (tagging for B), rs1053878 (tagging for A2), rs8176719 (tagging for O1) and rs2519093 (tagging for A1). As a result, we observed that the Covid-19 susceptibility variant was fully tagging A1 and A2 haplotypes, and partially tagging B haplotypes (20% of B haplotypes) (**Figure S49**), suggesting an increased risk of infection for A1 and A2 carriers.

#### APOE

The *APOE* variant tagging for the APOE-ε4 haplotype was initially significantly associated with Covid-19 susceptibility in EUR (rs429358-C, MAF = 0.15, OR = 1.38 [1.24; 1.53], P = 1.80 × 10^−9^, on 07.14.20). This signal was notably the first report of a genetic determinant for Covid-19 susceptibility.^24^ This previous report was based on UKB data but this signal was not replicated in an independent dataset, and this association was greatly attenuated after the summer, when the number of Covid-19 cases started to rise significantly and the mean age of infected participants decreased (**Figure 1**). The interaction between the age of participants and the APOE variant was significant (P = 1.78 × 10^−9^), which was further confirmed using a 2df test^25^ with age and genotype (P = 2.65 × 10^−9^), suggesting that a subset of older participant carriers of this variant was more at risk of Covid-19 infection. Remarkably, this signal became genome-wide significant in the 05.09.21 data release analyses of death in ALL and EUR, and in the analysis of Severe Covid-19 in EUR, which may suggest a mechanism leading to severe and lethal complications after infection. However, there is no evidence of association in the larger Covid-19hgi meta-analysis of Covid-19 severity (A2, freeze 5 without UKB, P = 0.27).

#### Results from ancestry- and sex-stratified analyses

In other ancestry-stratified GWAS (AFR, SAS and OTHERS), no signal was found genome-wide significant in the last data release analyzed. In the GWAS combining all non-European participants (nEUR), with Population controls, one signal was found significant at the *LRMDA* locus on chromosome 10 (rs114026383-C, MAF = 0.02, OR = 2.39 [1.79; 3.20], P = 4.10 × 10^−9^). According to gnomAD (v2.1.1),^26^ this variant is mostly carried by individuals of African ancestry (MAF = 0.04) and mainly absent in other ancestries. In the GWAS of African ancestry participants, this signal is close to genome wide significance (P = 1.55 × 10^−7^). Interestingly, *LRMDA* variants have been found associated to lung function^27^ [MIM: 608852] and HIV viral load in an unadjusted GWAS,^28^ but there was no evidence of LD between these variants and rs114026383 (r^2^ < 0.01). Furthermore, this association did not replicate in the Covid-19hgi susceptibility analysis (C2) restricted to African ancestry participants (P=0.27).

In addition, the chr3p21.31 susceptibility variant (rs73062389) was less frequent in non Europeans, and we did not observe an association with this variant in any of the non European ancestry-stratified analyses (MAF = 0.007 and P = 0.22 in AFR; MAF = 0.021 and P = 0.47 in SAS; MAF = 0.038 and P = 0.79 in OTHERS). Similarly, there was no association at the ABO lead variant (rs9411378) in non European ancestry-stratified analyses (P = 0.18 in AFR; P = 0.22 in SAS; P = 0.34 in OTHERS).

In sex-stratified analyses, using Population controls, the chr3p21.31 susceptibility signal was significant in women (rs73062389, P = 1.06 × 10^−8^ in ALL) and moderatly associated in men (P = 2.10 × 10^−6^ in ALL), whereas the ABO signal was significant in men (rs9411378, P = 5.10 × 10^−10^ in ALL) and moderatly associated in women (P = 3.30 × 10^−5^ in ALL). The chr3p21.31 lead variant in the hospitalization analysis (rs72893671) was more significant in men (P = 6.80 × 10^−11^ in ALL) than in women (P = 3.68 × 10^−4^ in ALL). Despite these differences in significance between men and women for these 3 main signals, we did not observe a significant difference of effect when using the Z-test for the equality of regression coefficients (P > 0.05 for all 3 signals). Additionally, a variant reached genome-wide significance in the analysis of Covid-19 susceptibility of women of ALL ancestry at the chr2q32.3 locus, while no association was observed for this variant in men (P = 0.47). However this association was not supported by the Covid-19hgi C2 analysis (P = 0.58), even though the Covid-19hgi meta-analyses were not sex-stratified.

### Signals with a positive significance trajectory in UKB

The significance trajectory of the most robust signals, at the chr3p21.31 and ABO loci, mostly increased after the surge of new cases following the 09.08.20 data release (**Figure 1**). After this date, the chr3p21.31 signal increased at each new data release and reached genome-wide significance in the 11.03.20 release, while the *ABO* signal also increased and reached significance in the 01.04.21 release. In order to identify signals that may become significant in future releases, we extracted variants displaying a similar positive trajectory in significance over time, and reaching at least P < 10^−4^ in the last data release analyzed.

After applying our filters to each of the 65 analyses, the number of variants with a positive significance trajectory summed up to 61,430. For each analysis, we extracted the lead variant at each locus, which resulted into 11,821 lead variants (**Table S6**), with some duplicates, as lead variants from a same locus can appear in several analyses (*i*.*e*. the chr3p21.31 locus, which appears in 49 out of the 65 analyses). Notably, some of these loci previously reached genome-wide significance in the Covid-19hgi meta-analyses^10^: the *DPP9* locus [MIM: 608258] associated with hospitalization, the *NXPE3* / *RPL24* locus [MIM: 604180] associated with susceptibility, and the *MAPT* locus [MIM: 157140] associated with severe Covid-19.

In order to identify which variants with a positive trajectory reached at least nominal significance in the Covid-19hgi analyses, we further focused on loci with positive significance trajectory from the susceptibility, hospitalization and severe analyses, using ALL ancestry and Population controls, which have the closest phenotype definition to the susceptibility (C2), hospitalization (B2) and severity (A2) meta-analyses from Covid-19hgi, respectively. Signals with a positive trajectory from Covid-19 susceptibility and hospitalization GWAS were sought for replication in the corresponding C2 and B2 meta-analyses from Freeze 6 without UKB, while signals from the severe Covid-19 GWAS were sought for replication in the A2 meta-analysis from Freeze 5 without UKB. We identified 205 loci with a positive trajectory in the susceptibility analysis, 193 with hospitalization, and 168 with severity. Out of these 566 signals, 430 were present in the corresponding Covid-19hgi meta-analyses, of which 30 (7%) had a concordant effect direction and P < 0.05 (**Table S7**). Notably, we observed a stringent gap in P-values from the Covid-19hgi meta-analyses results between the top 7 signals, previously identified and reaching P < 10^−16^ (chr3p21.31, *ABO, DPP9*, and *RPL24*), and the remaining 23 signals which did not reach P < 10^−3^ (**Table 4**). Furthermore, amongst the variants missing in the Covid-19hgi meta-analyses figured the lead variant at the *MAPT* locus (rs532052263), which had a positive trajectory in the UKB severe analysis. However, we observed significant LD between this variant and the known signal associated with severe Covid-19^10^ at this locus (rs8080583, r^2^ = 0.75), suggesting they both belong to the same signal.

**Table 4.** Signals with a positive significance trajectory in UKB, sorted by their P-value in their equivalent Covid-19hgi meta-analyses (top 10)

| Phenotype | CHR:POS:NEA:EA | UKB |  |  |  |  | COVID-19hgi (without UKB) |  |  |  | Locus |
| --- | --- | --- | --- | --- | --- | --- | --- | --- | --- | --- | --- |
|  |  | rsid | MAF | BETA | SE | P | ANALYSIS | BETA | SE | P |  |
| Hospitalization | 3:45848760:G:C | rs17712877 | 0.07 | 0.340 | 0.050 | 9.28E-12 | B2 | 0.387 | 0.023 | 8.01E-64 | SLC6A20;LZTFL1 |
| Susceptibility | 9:136149229:T:C | rs505922 | 0.32 | 0.071 | 0.012 | 1.04E-09 | C2 | 0.074 | 0.005 | 2.44E-60 | ABO |
| Susceptibility | 3:45835417:G:A | rs73062389 | 0.06 | 0.188 | 0.024 | 4.26E-15 | C2 | 0.158 | 0.010 | 1.08E-54 | SLC6A20 |
| Severe | 3:45889921:A:T | rs35081325 | 0.07 | 0.468 | 0.081 | 6.65E-09 | A2 | 0.633 | 0.044 | 5.29E-46 | LZTFL1 |
| Hospitalization | 19:4723670:C:A | rs2277732 | 0.30 | 0.113 | 0.028 | 4.64E-05 | B2 | 0.131 | 0.014 | 4.26E-22 | DPP9 |
| Hospitalization | 9:136149500:T:C | rs529565 | 0.32 | 0.107 | 0.027 | 7.12E-05 | B2 | 0.106 | 0.012 | 7.10E-18 | ABO |
| Susceptibility | 3:101503765:G:A | rs4342086 | 0.35 | -0.046 | 0.012 | 6.62E-05 | C2 | -0.039 | 0.005 | 8.22E-17 | NXPE3 |
| Hospitalization | 4:25464527:T:C | rs12507905 | 0.13 | 0.178 | 0.037 | 1.69E-06 | B2 | 0.053 | 0.016 | 1.04E-3 | ANAPC4;LOC101929161 |
| Hospitalization | 17:1441448:A:G | rs6502772 | 0.18 | 0.130 | 0.033 | 9.56E-05 | B2 | 0.056 | 0.018 | 1.42E-3 | PITPNA |
| Hospitalization | 1:77954397:G:C | rs12034334 | 0.18 | 0.133 | 0.033 | 6.71E-05 | B2 | 0.051 | 0.016 | 1.51E-3 | AK5 |

<sup>a</sup> Indicate the phenotype, the ancestry, the control set, and, when it is the case, the corresponding sex-stratified analyses (F: Females, M: Males) CHR: Chromosome; POS: Position (hg19 genome build); NEA: Non Effect Allele; EA: Effect Allele; MAF: Minor Allele Frequency; SE: Standard Error; P: P-value (from the last data release, analyzed on 06.18.21 for susceptibility and hospitalization, and 05.09.21 for severe and death phenotypes)

### Signals with a positive significance trajectory in Covid-19hgi meta-analyses

Finally, given the larger sample size and statistical power of the Covid-19hgi studies, we applied the significance trajectory analyses to these datasets, where this approach might be more effective to identify signals of interest. For the B2 and C2 analyses, we analyzed freezes 2 to 6, and we analyzed freezes 3 to 6 for the A2 analysis (all including the UKB dataset, but not the 23andMe dataset). For this significance trajectory analysis, the same set of rules used for the UKB analyses was applied, except that we only extracted variants reaching P < 10^−5^ in the last freeze (instead of P < 10^−4^ in UKB), since these meta-analyses are better powered than the UKB analyses. Over all 3 analyses, we identified a total of 9,697 variants with a positive trajectory (**Table S8**). After clusterizing these variants into loci and extracting lead variants at each locus, we obtained 51 lead variants for the A2 analysis, 89 for B2 and 85 for C2 (**Table S9**).

Out of these 225 loci, 34 reached genome-wide significance, and only 3 known genome wide significant signals were not detected by the significance trajectory analysis: at the *CCHCR1* locus [MIM: 605310] (in both B2 and A2 analyses) and at the *MUC5B* locus [MIM: 600770] (in the B2 analysis), as both these loci decreased in significance by more than one order of magnitude at some point. Furthermore, 2 loci that reached genome-wide significance only when the 23andMe datasets were included in the Covid-19hgi meta-analyses were also detected by our approach: at the *SFTPD* locus [MIM: 178635] and at the *SLC22A31* locus (both in the B2 analysis). Additionaly, we detected signals at the *ACE2* [MIM: 300335] and *TMPRSS2* loci [MIM: 602060], 2 proteins involved in SARS-CoV-2 infection. A rare *ACE2* variant was previously observed as significantly associated in the C2 Covid-19hgi meta-analysis (rs190509934, MAF = 0.003, P = 3.30 × 10^−17^), but the significance trajectory analysis also revealed a common variant (rs4646120, MAF = 0.49, P = 2.30 × 10^−6^) associated with Covid-19 susceptibility. At the TMPRSS2 locus, a common exonic variant was detected in the A2 analysis (rs12329760, MAF = 0.27, P = 8.18 × 10^−6^), and other intronic variants in LD with the exonic variant were detected in the B2 analysis (rs915823, P = 1.67 × 10^−6^, LD r^2^= 0.83). Finally, several signals detected in this approach might be of interest as they are located in, or near, genes playing a role in immune response. For instance, intronic variants in the mucin genes *MUC4* [MIM: 158372] (rs2260685, MAF = 0.46, P = 1.76 × 10^−6^) and *MUC16* [MIM: 606154] (rs73005873, MAF = 0.36, P = 7.81 × 10^−8^) were identified in the C2 analysis, and 2 intergenic variants close to interferon regulatory factors *IRF1* [MIM: 147575] (rs4143335, MAF = 0.14, P = 5.96 × 10^−6^) and *IRF2* [MIM: 147576] (rs6854508, MAF = 0.17, P = 9.26 × 10^−6^) were identified in the B2 analysis.

## Discussion

This project was initiated with the major aim to share results of Covid-19 host genetics analyses freely and rapidly on the GRASP portal, during a pandemic where new insights to improve patient care and develop better treatments were greatly needed. A few months after the first Covid-19 case was identified in the UK, we started to perform GWAS on each dataset released by UKB between May 2020 and June 2021, and thus far examined genetic signals associated with Covid-19 phenotypes across 18 data releases. This unique context allowed us to track the evolution of genetic associations over time, an approach rarely applied. While some works have examined the statistical properties of phased and nested case-control studies,^29,30^ few studies have proposed using statistical significance trajectories over time in genetic analyses.

As a first observation, the majority of signals observed in the first stages of the project did not sustain over time. For instance, the first genome-wide significant association observed in the GWAS of Covid-19 susceptibility in Europeans with rs34338189 as lead variant is not even nominally significant in the last release (P = 0.25). Another genome-wide significant signal was observed at the *APOE* locus, with a significance that increased in the first 4 data releases before decreasing continually in the subsequent releases. The lead variant at the *APOE* locus is coding in part for the APOE-ε4 haplotype, known to increase the risk of Alzheimer’s disease [MIM: 607822], dementia, dyslipidemia and cardiovascular diseases [MIM: 617347] and is speculated to cause inflammation and cytokine storms.^31^ Notably, the significant association of the *APOE* signal with severe Covid-19 and death in most recent analyses could support this proposition, but this association has not been replicated in an independent dataset. The evolution of the *APOE* signal over time could also be due to an initial higher prevalence of Covid-19 in nursing homes,^32^ where dementia patients were at higher risk of being infected and spreading the virus due to living arrangements, and a poor understanding of transmission dynamics and appropriate safety guidelines early in the pandemic. Overall, the evolution of these signals suggests a change in the composition of cases over time, such as the diminution of age and increase of positively tested women in the later data releases, as well as the introduction of variant SARS-CoV2 strains. This change was most significant after the 2020 summer when a surge in new Covid-19 infections occurred.

The most robust findings from our study are the association of the chr3p21.31 and ABO loci with Covid-19 susceptibility and a distinct signal at the chr3p21.31 locus associated with Hospitalization and severe Covid-19. These observations corroborate several previous reports,^6– 8,10^. In order to detect additional suggestive signals, that may reach genome-wide significance in future analyses, we developed an original strategy to identify associations displaying increased significance over time, and identified thousands of variants with a positive significance trajectory. Amongst these, 3 signals at the *DPP9, NXPE3*, and *MAPT* loci were previously established variants associated with Covid-19 phenotypes, notably by the Covid-19hgi analyses,^10^ thus demonstrating the potential of this approach. However, determining which variants actually modulate Covid-19 phenotypes amongst thousands of candidates, without an independent and well-powered dataset such as Covid-19hgi to confirm the validity of a signal, is still challenging. Still, this approach might be valuable to reduce the scope of search. To further reduce the scope of candidates, we applied this significance trajectory analysis to the larger meta-analyses provided by the Covid-19hgi. Given the higher statistical power of these analyses, we also applied a more stringent threshold, which allowed us to extract only variants reaching P < 10^−5^ in the last freeze. As a result, a lower and thus more workable number of loci were extracted, among which several have a role in immune function, such as mucins, interferon regulators, interleukin receptors, or HLA system related genes, which might be of interest for the study of genetic mechanisms involved in Covid-19 phenotypes. However, these associations are still only suggestive, and will need to be further validated either in future releases or in independent datasets.

The associations we observed changing through the pandemic could reflect random effects or changes in statistical power, but some of the results suggest changes due to potential gene-environment interactions such as age, underlying health conditions (*APOE*) or sex makeup of cases exposed to or engaging in risk behavior. This indicates the general approach of iterative analysis and significance trajectory analyses for genetics during pandemics may have benefits in uncovering pathophysiologic clues. Additionally, other factors like predominant virus strains and changing treatment strategies through a pandemic might interact with host genetics, and be better understood by iterative analyses. To enable such approaches in pandemics and other critical research domains, rapid and extensive results sharing are important catalysts, as demonstrated by the UKBB, COLCORONA, GenOMICC and Covid-19hgi.

In summary, our host genomic analyses of Covid-19 may contribute to improve the comprehension of mechanisms involved in the infection and complications due to Covid-19. Our study also has some limitations. Most importantly, our data and the work of others support large health disparities between EUR and non-EUR individuals related to COVID-19 throughout the ongoing pandemic. Despite an over-representation proportionally among cases and those with severe and fatal outcomes, the non-EUR component of UKB and Covid-19hgi is a proportionally small sample limiting our statistical power to address population-specific genetic variants contributing to health outcomes. However, moving forward we feel that having a diverse set of results with different phenotype definitions, sex-specific, ancestry-specific, and including external group summary statistics, all in a common genome reference and annotation framework may maximize the chance for new studies to cross-replicate or meta-analyze results as Covid-19 genetic studies continue to grow.

## Data Availability

The datasets generated during this study are available at the GRASP Covid-19 portal

https://grasp.nhlbi.nih.gov/Covid19GWASResults.aspx

## Description of Supplemental Data

Supplemental Data include 50 figures and 9 tables.

## Acknowledgments

All authors were supported by NIH Intramural Research Program funds. The views expressed in this manuscript are those of the authors and do not necessarily represent the views of the National Heart, Lung, and Blood Institute; the National Institutes of Health; or the U.S. Department of Health and Human Services. This research has been conducted using the UK Biobank Resource under Application Number 28525. UK Biobank was established by the Wellcome Trust, Medical Research Council, Department of Health, Scottish government, and Northwest Regional Development Agency. It has also had funding from the Welsh assembly government and the British Heart Foundation. All UKB analyses for this manuscript were conducted on the NIH Biowulf high performance computing cluster (https://hpc.nih.gov/). The Genotype-Tissue Expression (GTEx) Project was supported by the Common Fund of the Office of the Director of the National Institutes of Health, and by NCI, NHGRI, NHLBI, NIDA, NIMH, and NINDS. We also thank the NHLBI IT team for their help in keeping the GRASP portal up to date, David-Alexandre Tregouet for his helpful comments regarding the ABO haplotype analyses, and the Covid-19 Host Genetics Initiative for sharing the results of their analyses.

## Web Resources

GRASP Covid-19 resource: https://grasp.nhlbi.nih.gov/Covid19GWASResults.aspx

GTeX: https://gtexportal.org/home/

CADD: https://cadd.gs.washington.edu/

EBI GWAS catalog: https://www.ebi.ac.uk/gwas/

GRASP catalog: https://grasp.nhlbi.nih.gov/Overview.aspx

Covid-19hgi meta-analyses results: https://www.covid19hg.org/results/r5/

GnomAD: https://gnomad.broadinstitute.org/

LocusZoom: https://locuszoom.org/

LDlink: https://ldlink.nci.nih.gov

## Data and Code Availability

The datasets generated during this study are available at the GRASP Covid-19 portal: https://grasp.nhlbi.nih.gov/Covid19GWASResults.aspx

## References

1. Connors, J.M., and Levy, J.H. (2020). COVID-19 and its implications for thrombosis and anticoagulation. Blood 135, 2033–2040.

2. Tang, D., Comish, P., and Kang, R. (2020). The hallmarks of COVID-19 disease. PLoS Pathog. 16, e1008536.

3. Kwok, A.J., Mentzer, A., and Knight, J.C. (2021). Host genetics and infectious disease: new tools, insights and translational opportunities. Nat. Rev. Genet. 22, 137–153.

4. Dragic, T., Litwin, V., Allaway, G.P., Martin, S.R., Huang, Y., Nagashima, K.A., Cayanan, C., Maddon, P.J., Koup, R.A., Moore, J.P., et al. (1996). HIV-1 entry into CD4+ cells is mediated by the chemokine receptor CC-CKR-5. Nature 381, 667–673.

5. Ciancanelli, M.J., Huang, S.X.L., Luthra, P., Garner, H., Itan, Y., Volpi, S., Lafaille, F.G., Trouillet, C., Schmolke, M., Albrecht, R.A., et al. (2015). Life-threatening influenza and impaired interferon amplification in human IRF7 deficiency. Science 348, 448–453.

6. Ellinghaus, D., Degenhardt, F., Bujanda, L., Buti, M., Albillos, A., Invernizzi, P., Fernández, J., Prati, D., Baselli, G., Asselta, R., et al. (2020). Genomewide Association Study of Severe Covid-19 with Respiratory Failure. N. Engl. J. Med. 0, null.

7. Pairo-Castineira, E., Clohisey, S., Klaric, L., Bretherick, A.D., Rawlik, K., Pasko, D., Walker, S., Parkinson, N., Fourman, M.H., Russell, C.D., et al. (2020). Genetic mechanisms of critical illness in Covid-19. Nature 1–1.

8. Shelton, J.F., Shastri, A.J., Ye, C., Weldon, C.H., Filshtein-Sonmez, T., Coker, D., Symons, A., Esparza-Gordillo, J., Aslibekyan, S., and Auton, A. (2021). Trans-ancestry analysis reveals genetic and nongenetic associations with COVID-19 susceptibility and severity. Nat. Genet. 1–8.

9. Kosmicki, J.A., Horowitz, J.E., Banerjee, N., Lanche, R., Marcketta, A., Maxwell, E., Bai, X., Sun, D., Backman, J.D., Sharma, D., et al. (2021). Pan-ancestry exome-wide association analyses of COVID-19 outcomes in 586,157 individuals. Am. J. Hum. Genet. 0,.

10. COVID-19 Host Genetics Initiative (2021). Mapping the human genetic architecture of COVID-19. Nature.

11. Sudlow, C., Gallacher, J., Allen, N., Beral, V., Burton, P., Danesh, J., Downey, P., Elliott, P., Green, J., Landray, M., et al. (2015). UK Biobank: An Open Access Resource for Identifying the Causes of a Wide Range of Complex Diseases of Middle and Old Age. PLOS Med. 12, e1001779.

12. Bycroft, C., Freeman, C., Petkova, D., Band, G., Elliott, L.T., Sharp, K., Motyer, A., Vukcevic, D., Delaneau, O., O’Connell, J., et al. (2018). The UK Biobank resource with deep phenotyping and genomic data. Nature 562, 203–209.

13. Zhou, W., Nielsen, J.B., Fritsche, L.G., Dey, R., Gabrielsen, M.E., Wolford, B.N., LeFaive, J., VandeHaar, P., Gagliano, S.A., Gifford, A., et al. (2018). Efficiently controlling for case-control imbalance and sample relatedness in large-scale genetic association studies. Nat. Genet. 50, 1335–1341.

14. Machiela, M.J., and Chanock, S.J. (2015). LDlink: a web-based application for exploring population-specific haplotype structure and linking correlated alleles of possible functional variants. Bioinforma. Oxf. Engl. 31, 3555–3557.

15. Boughton, A.P., Welch, R.P., Flickinger, M., VandeHaar, P., Taliun, D., Abecasis, G.R., and Boehnke, M. (2021). LocusZoom.js: Interactive and embeddable visualization of genetic association study results. Bioinforma. Oxf. Engl. btab186.

16. Wang, K., Li, M., and Hakonarson, H. (2010). ANNOVAR: functional annotation of genetic variants from high-throughput sequencing data. Nucleic Acids Res. 38, e164–e164.

17. Rentzsch, P., Witten, D., Cooper, G.M., Shendure, J., and Kircher, M. (2019). CADD: predicting the deleteriousness of variants throughout the human genome. Nucleic Acids Res. 47, D886–D894.

18. Eicher, J.D., Landowski, C., Stackhouse, B., Sloan, A., Chen, W., Jensen, N., Lien, J.-P., Leslie, R., and Johnson, A.D. (2015). GRASP v2.0: an update on the Genome-Wide Repository of Associations between SNPs and phenotypes. Nucleic Acids Res. 43, D799–804.

19. Buniello, A., MacArthur, J.A.L., Cerezo, M., Harris, L.W., Hayhurst, J., Malangone, C., McMahon, A., Morales, J., Mountjoy, E., Sollis, E., et al. (2019). The NHGRI-EBI GWAS Catalog of published genome-wide association studies, targeted arrays and summary statistics 2019. Nucleic Acids Res. 47, D1005–D1012.

20. Aguet, F., Brown, A.A., Castel, S.E., Davis, J.R., He, Y., Jo, B., Mohammadi, P., Park, Y., Parsana, P., Segrè, A.V., et al. (2017). Genetic effects on gene expression across human tissues. Nature 550, 204–213.

21. Zhang, X., Gierman, H.J., Levy, D., Plump, A., Dobrin, R., Goring, H.H.H., Curran, J.E., Johnson, M.P., Blangero, J., Kim, S.K., et al. (2014). Synthesis of 53 tissue and cell line expression QTL datasets reveals master eQTLs. BMC Genomics 15, 532.

22. Dubé, M.-P., Lemaçon, A., Barhdadi, A., Lemieux Perreault, L.-P., Oussaïd, E., Asselin, G., Provost, S., Sun, M., Sandoval, J., Legault, M.-A., et al. (2021). Genetics of symptom remission in outpatients with COVID-19. Sci. Rep. 11, 10847.

23. Goumidi, L., Thibord, F., Wiggins, K.L., Li-Gao, R., Brown, M.R., van Hylckama Vlieg, A., Souto, J.-C., Soria, J.-M., Ibrahim-Kosta, M., Saut, N., et al. (2021). Association between ABO haplotypes and the risk of venous thrombosis: impact on disease risk estimation. Blood 137, 2394–2402.

24. Kuo, C.-L., Pilling, L.C., Atkins, J.L., Masoli, J.A.H., Delgado, J., Kuchel, G.A., and Melzer, D. (2020). ApoE e4e4 Genotype and Mortality With COVID-19 in UK Biobank. J. Gerontol. A. Biol. Sci. Med. Sci. 75, 1801–1803.

25. Kraft, P., Yen, Y.-C., Stram, D.O., Morrison, J., and Gauderman, W.J. (2007). Exploiting gene-environment interaction to detect genetic associations. Hum. Hered. 63, 111–119.

26. Karczewski, K.J., Francioli, L.C., Tiao, G., Cummings, B.B., Alföldi, J., Wang, Q., Collins, R.L., Laricchia, K.M., Ganna, A., Birnbaum, D.P., et al. (2020). The mutational constraint spectrum quantified from variation in 141,456 humans. Nature 581, 434–443.

27. Soler Artigas, M., Loth, D.W., Wain, L.V., Gharib, S.A., Obeidat, M., Tang, W., Zhai, G., Zhao, J.H., Smith, A.V., Huffman, J.E., et al. (2011). Genome-wide association and large-scale follow up identifies 16 new loci influencing lung function. Nat. Genet. 43, 1082–1090.

28. Ekenberg, C., Tang, M.-H., Zucco, A.G., Murray, D.D., MacPherson, C.R., Hu, X., Sherman, B.T., Losso, M.H., Wood, R., Paredes, R., et al. (2019). Association Between Single-Nucleotide Polymorphisms in HLA Alleles and Human Immunodeficiency Virus Type 1 Viral Load in Demographically Diverse, Antiretroviral Therapy-Naive Participants From the Strategic Timing of AntiRetroviral Treatment Trial. J. Infect. Dis. 220, 1325–1334.

29. Van der Tweel, I., and van Noord, P.A. (2000). Sequential analysis of matched dichotomous data from prospective case-control studies. Stat. Med. 19, 3449–3464.

30. Boessen, R., van der Baan, F., Groenwold, R., Egberts, A., Klungel, O., Grobbee, D., Knol, M., and Roes, K. (2013). Optimizing trial design in pharmacogenetics research: comparing a fixed parallel group, group sequential, and adaptive selection design on sample size requirements. Pharm. Stat. 12, 366–374.

31. Numbers, K., and Brodaty, H. (2021). The effects of the COVID-19 pandemic on people with dementia. Nat. Rev. Neurol. 17, 69–70.

32. Chen, M.K., Chevalier, J.A., and Long, E.F. (2021). Nursing home staff networks and COVID-19. Proc. Natl. Acad. Sci. U. S. A. 118,.

