## Supplemental Figures for "A year of Covid-19 GWAS results from the GRASP portal reveals potential SARS-CoV-2 modifiers"

Figures S1-S46 : Evolution of genome wide significant signals associated with Covid-19 phenotypes (analysis described as ancestry:controls(:sex))

Figure S47 : Comparison of genome-wide significant signals in analyses using Population and Tested controls

Figure S48 : Regional association plots of Covid-19 susceptibility and severity in Europeans (Population as controls), at the chr3p21.31 locus

Figure S49: Haplotype analysis of ABO blood groups, including the variant associated with Covid-19 susceptibility (from the LDlink tool LDhap)

Figure S50: Evolution of the signals from Figure 1 (Susceptibility signals in EUR, from UKB) in Covid-19hgi C2 analyses

Supplemental Figure 1: Covid-19 Susceptibility in ALL:Pop

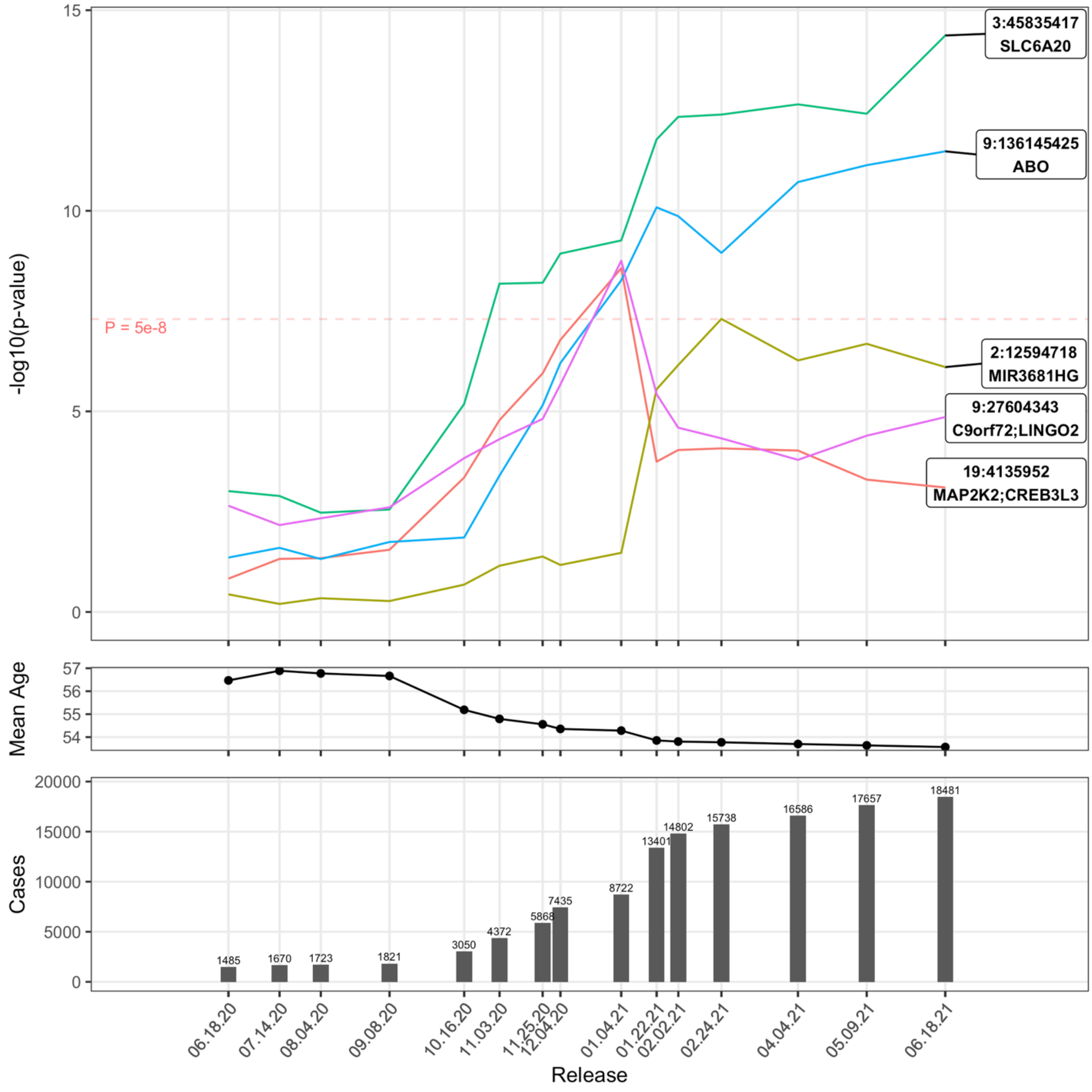

Supplemental Figure 2: Covid-19 Susceptibility in ALL:Pop:F

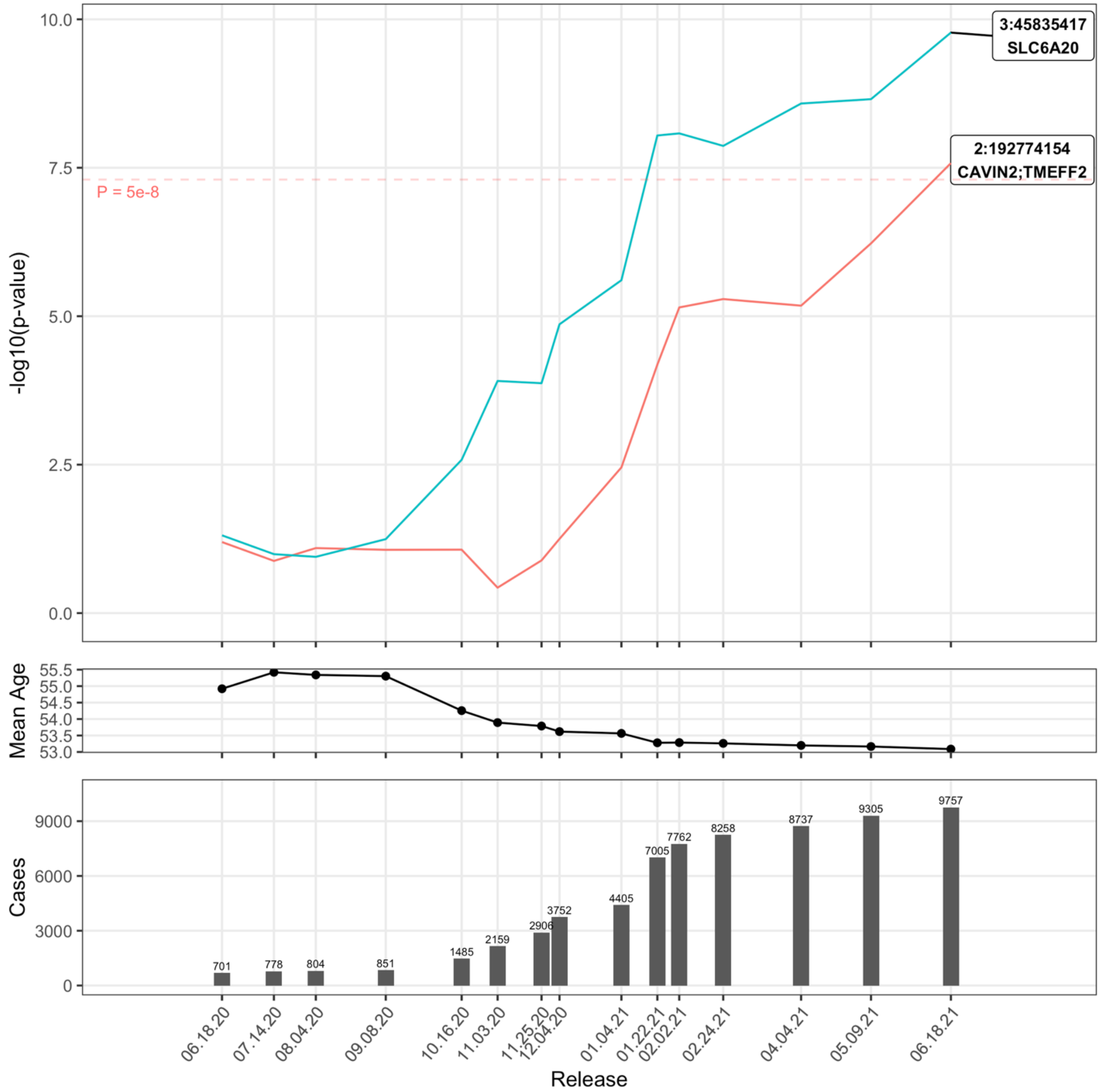

Supplemental Figure 3: Covid-19 Susceptibility in ALL:Pop:M

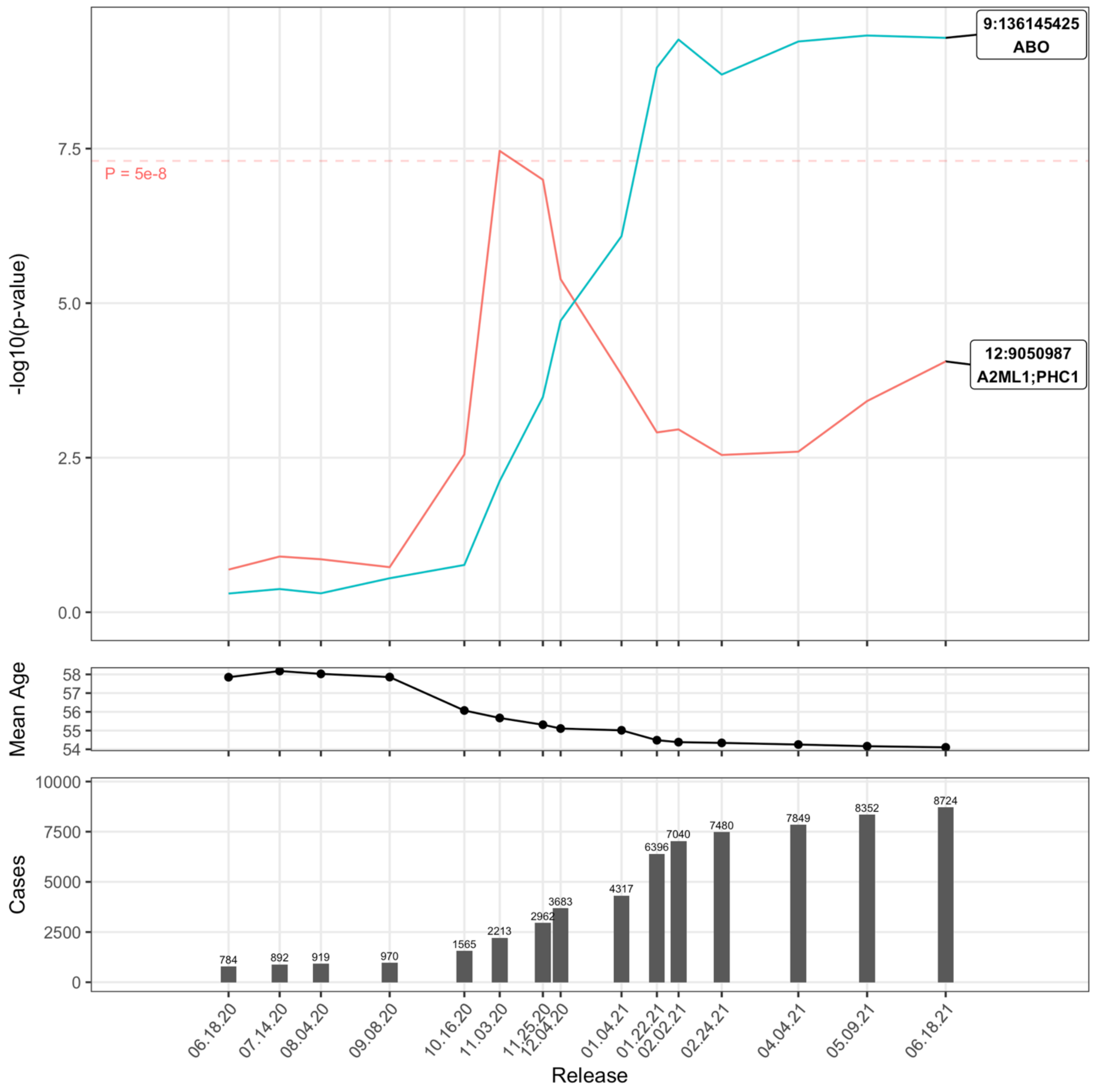

Supplemental Figure 4: Covid-19 Susceptibility in ALL:Tested

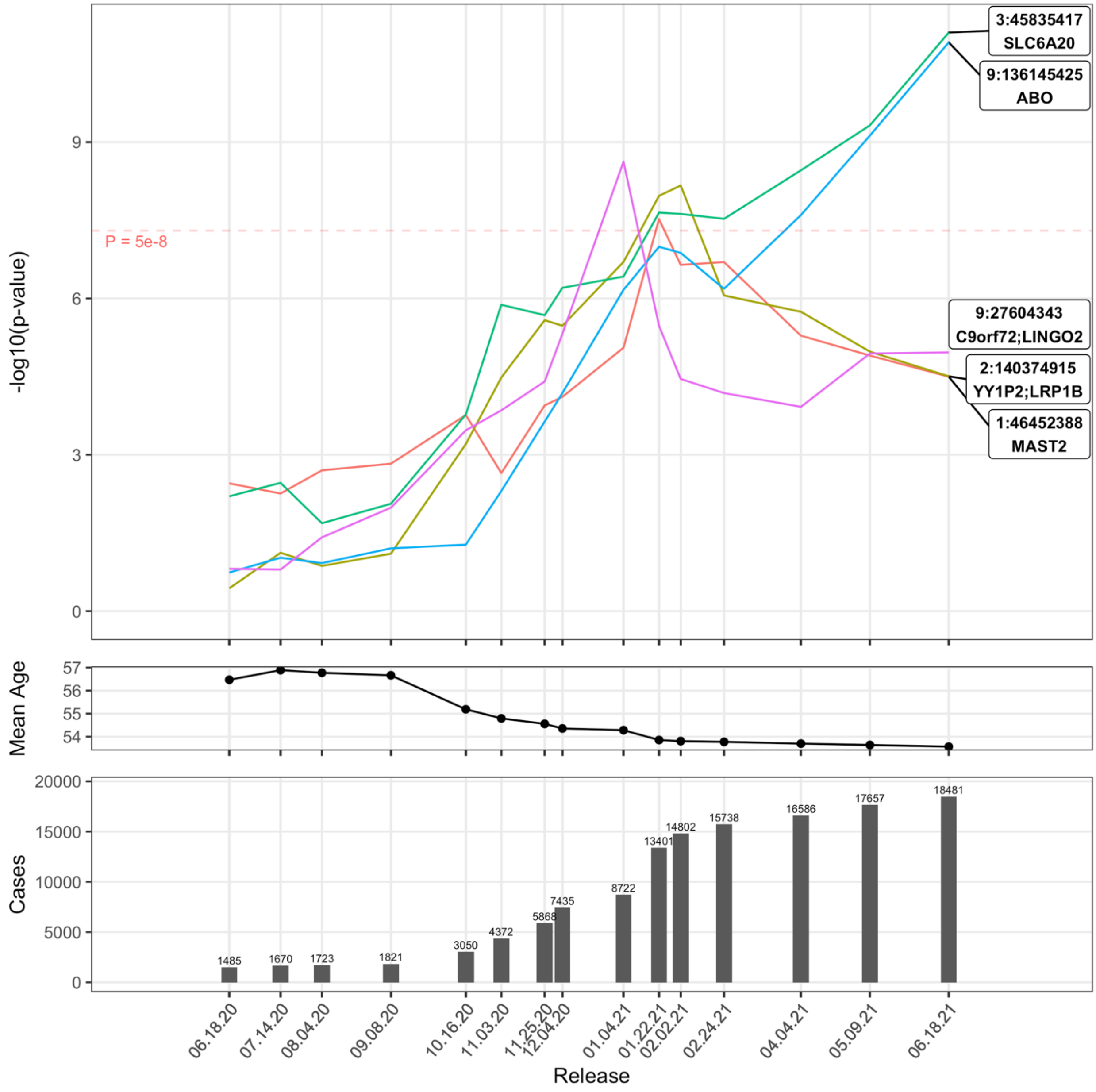

Supplemental Figure 5: Covid-19 Susceptibility in ALL:Tested:F

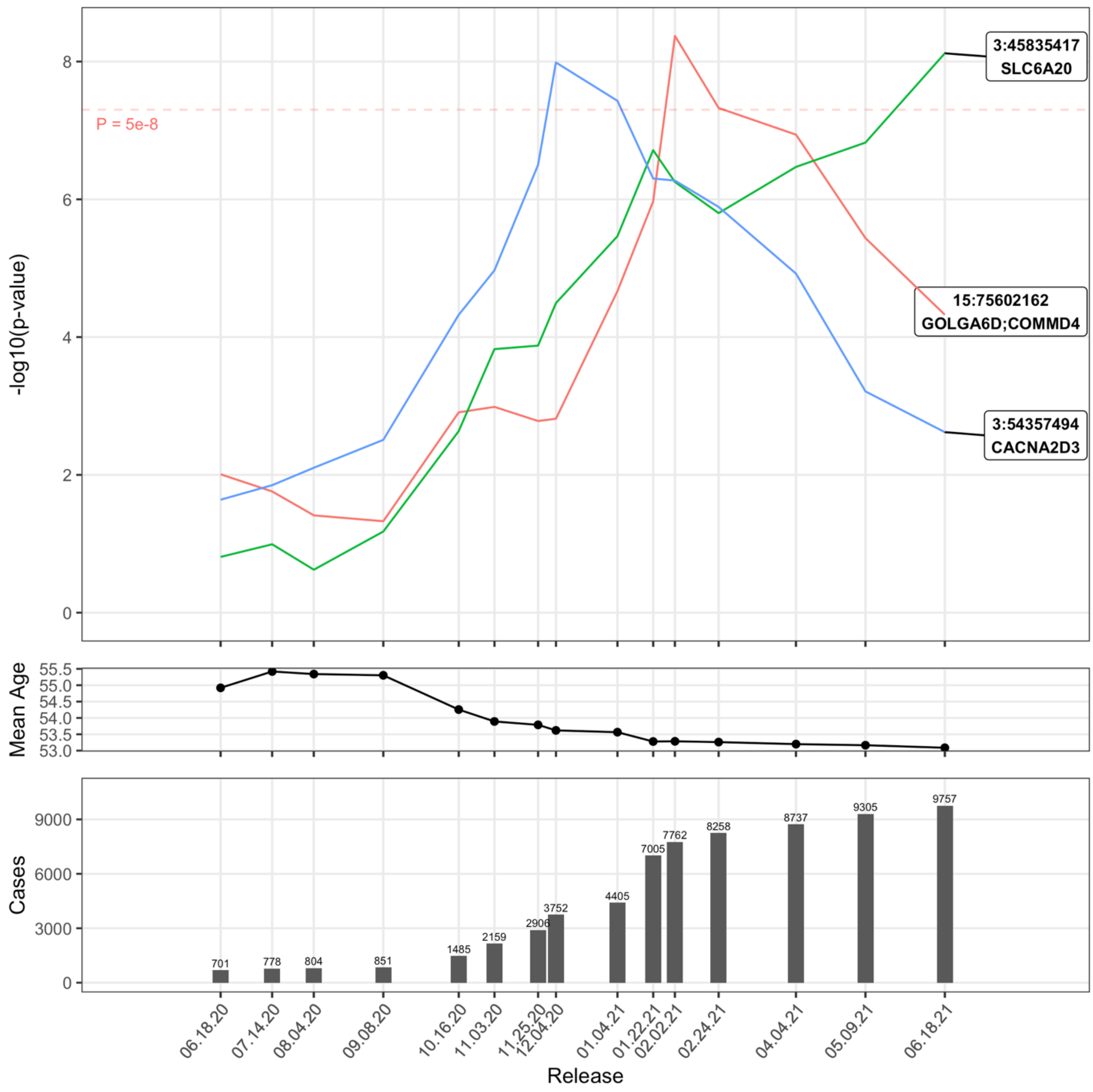

Supplemental Figure 6: Covid-19 Susceptibility in ALL:Tested:M

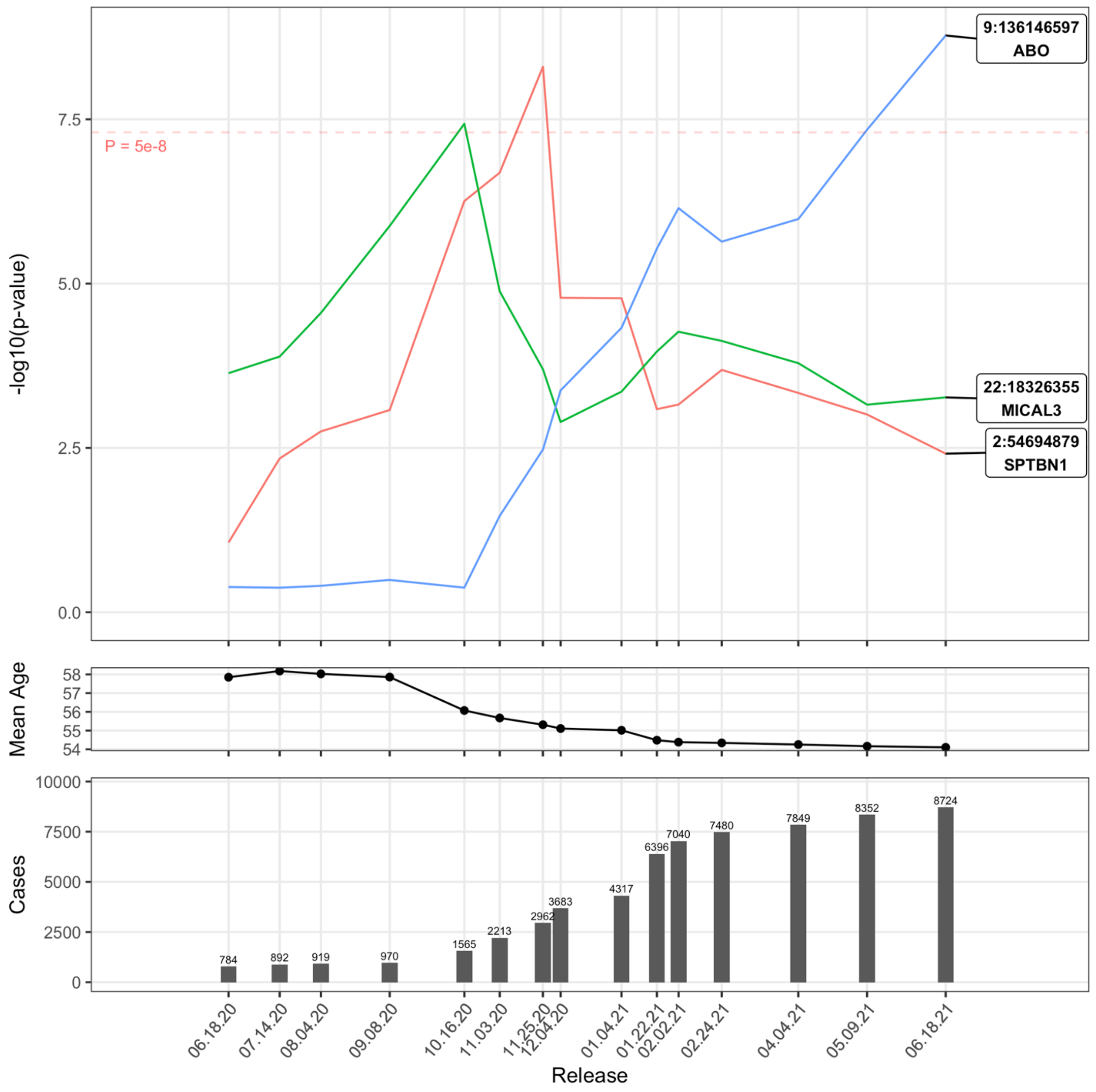

Supplemental Figure 7: Covid-19 Susceptibility in EUR:Pop

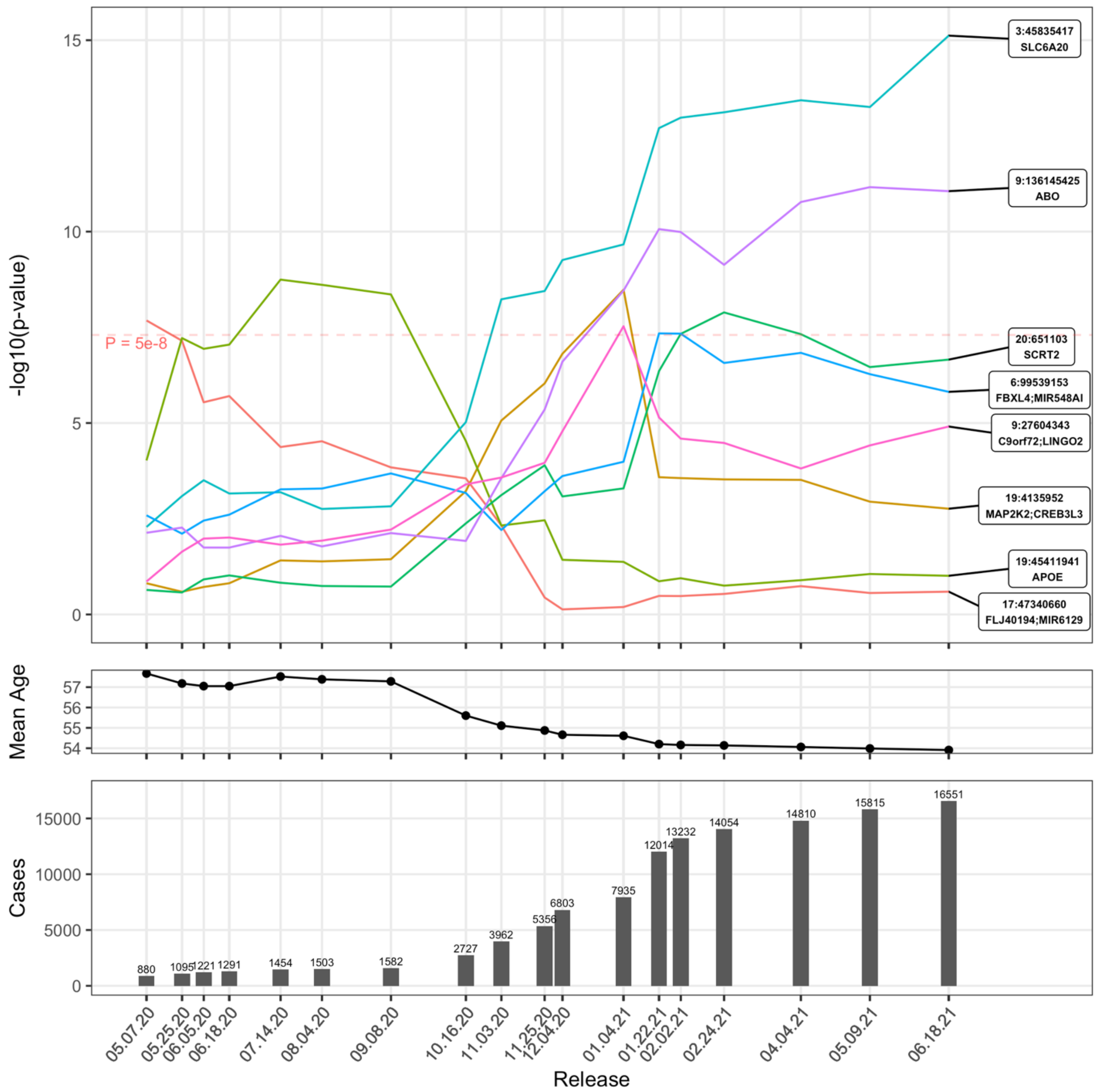

Supplemental Figure 8: Covid-19 Susceptibility in EUR:Pop:F

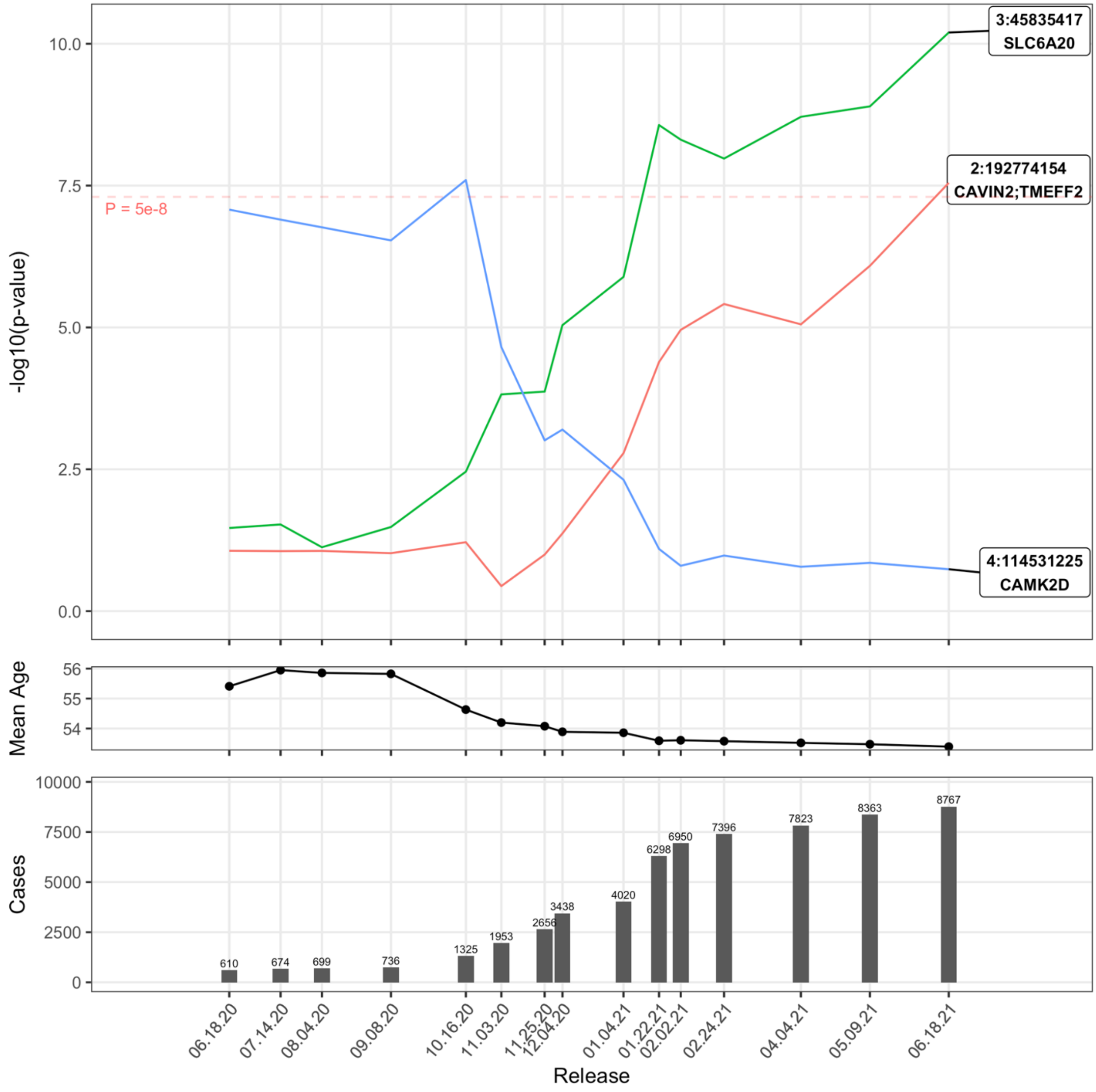

Supplemental Figure 9: Covid-19 Susceptibility in EUR:Pop:M

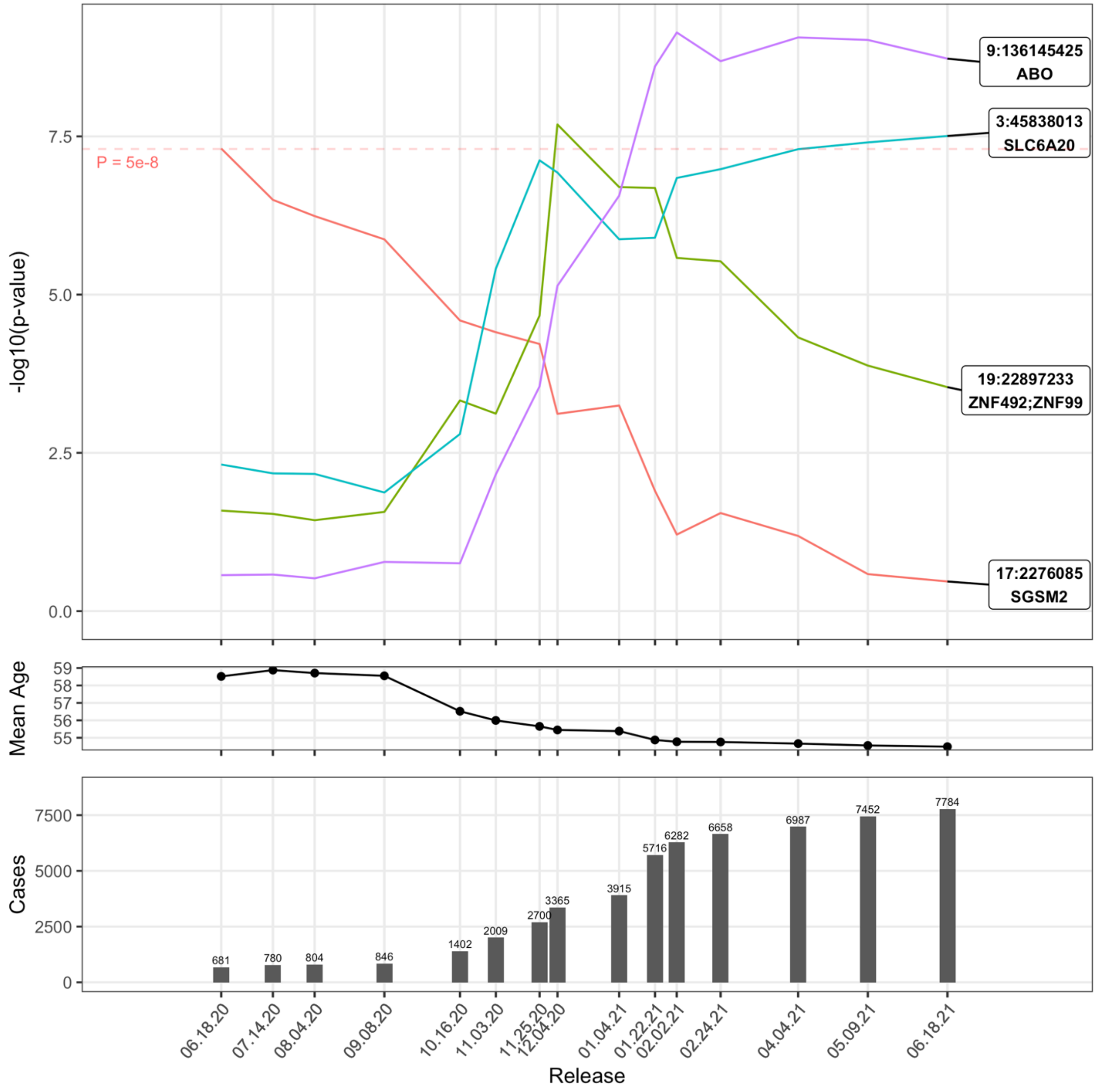

Supplemental Figure 10: Covid-19 Susceptibility in EUR:Tested

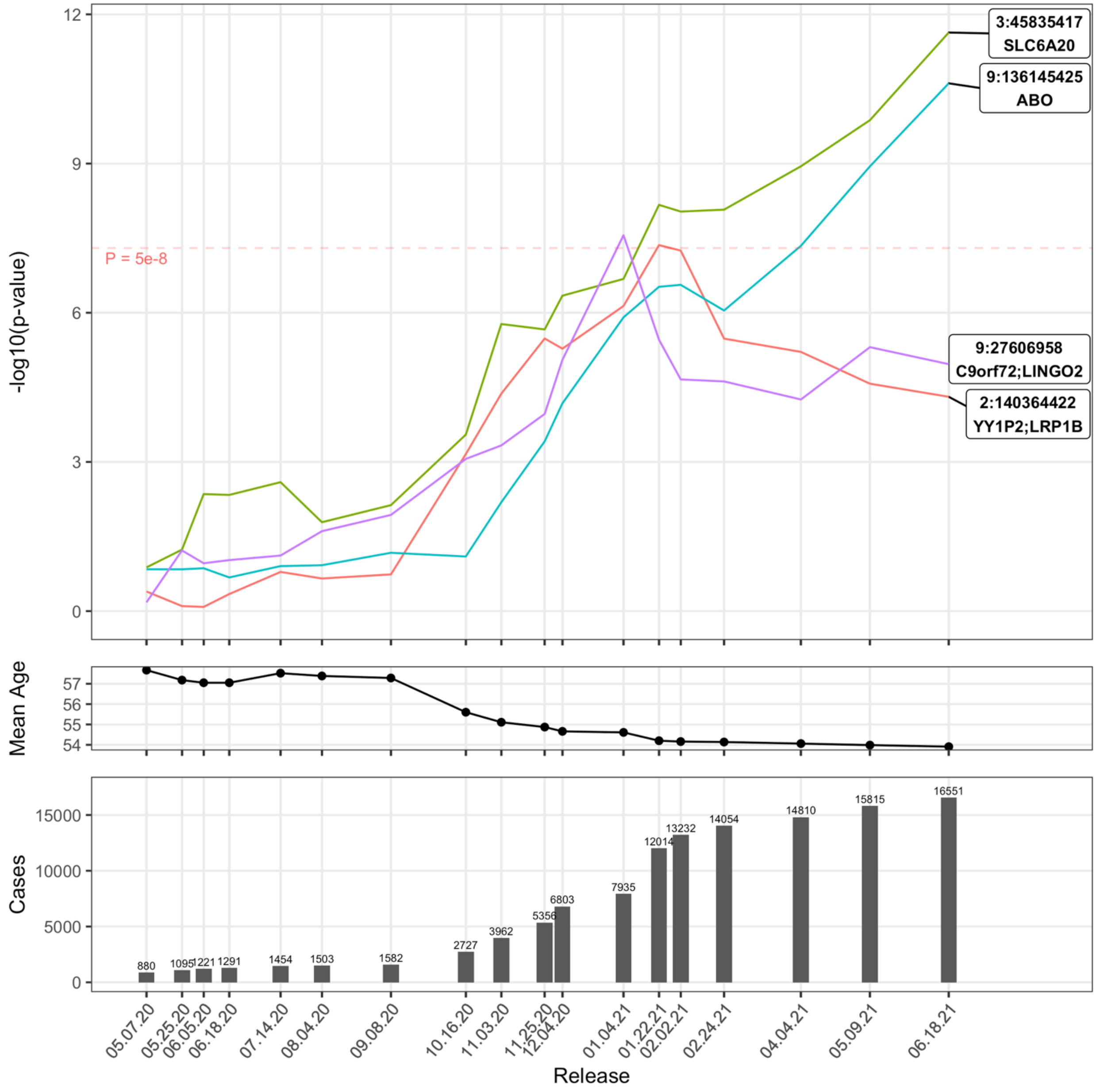

Supplemental Figure 11: Covid-19 Susceptibility in EUR:Tested:F

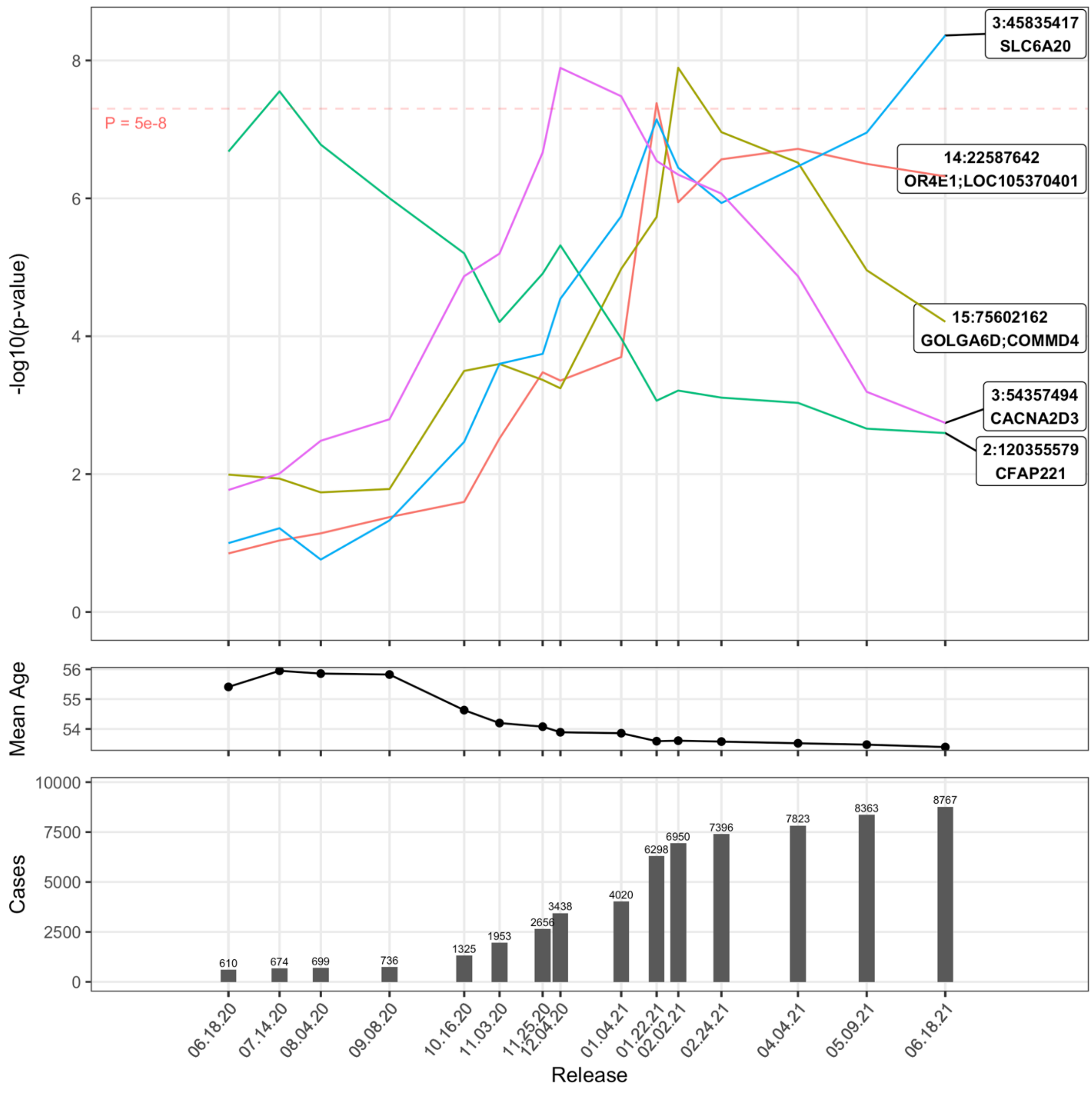

Supplemental Figure 12: Covid-19 Susceptibility in EUR:Tested:M

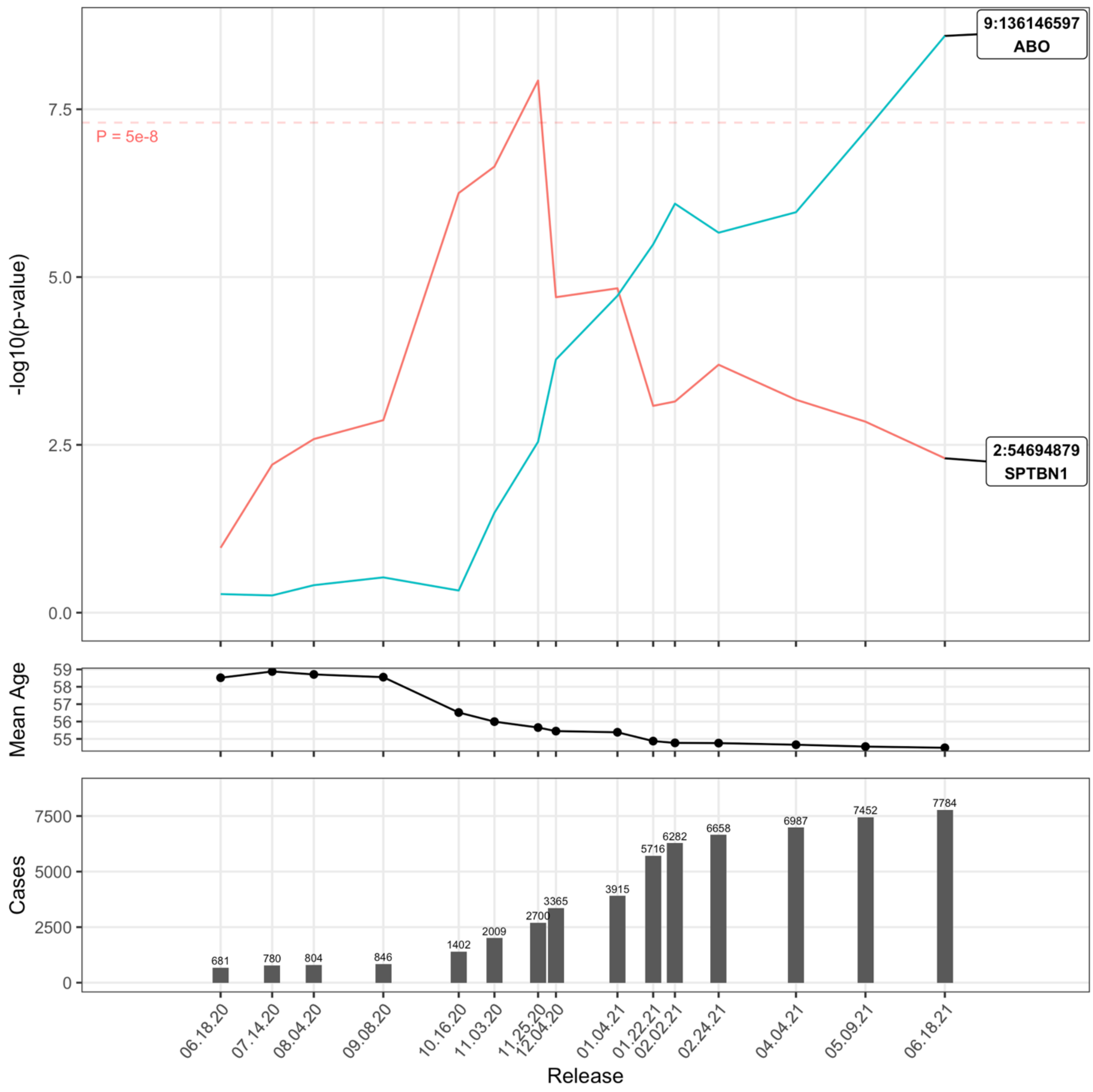

Supplemental Figure 13: Covid-19 Susceptibility in AFR:Pop

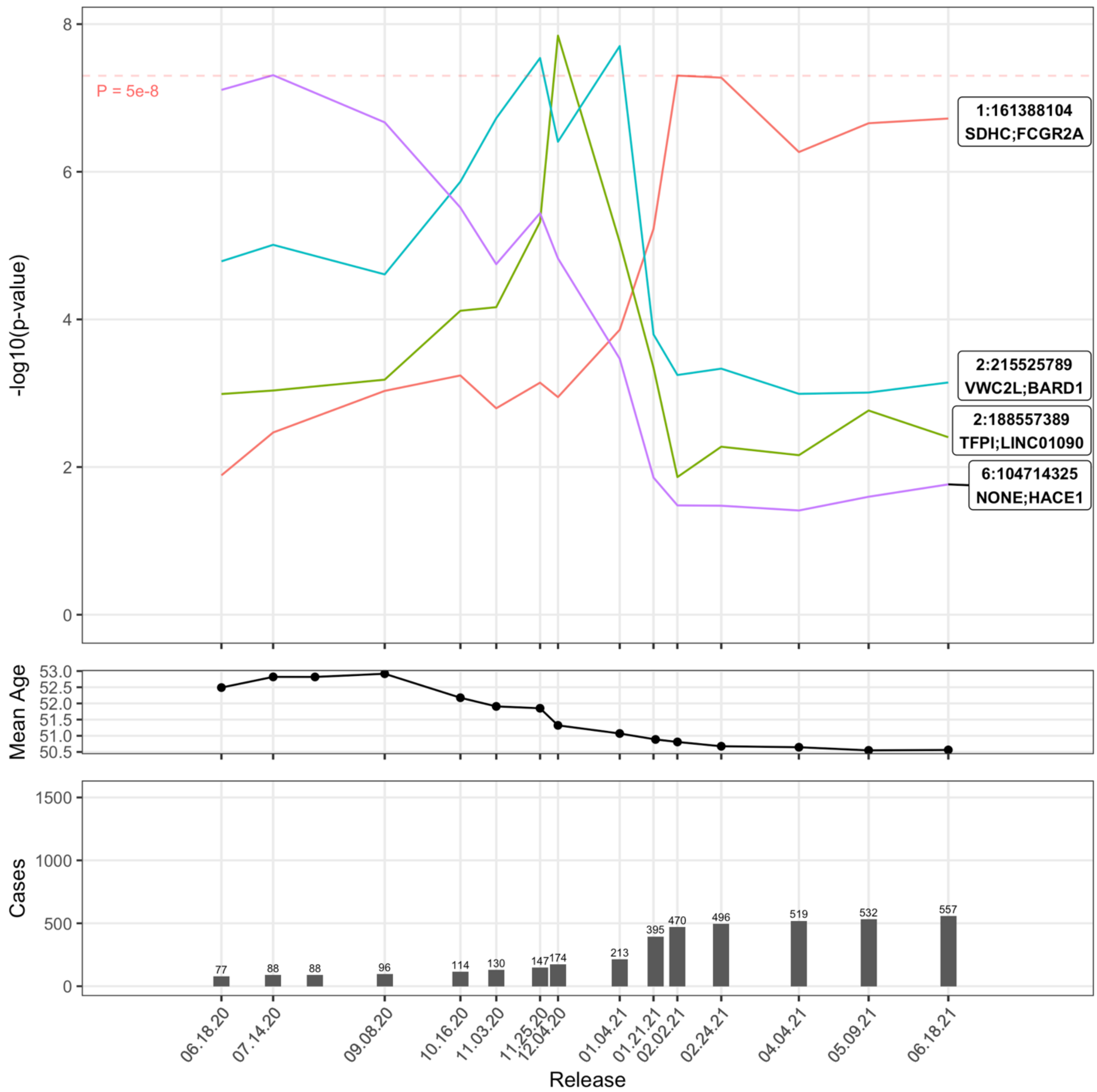

Supplemental Figure 14: Covid-19 Susceptibility in AFR:Tested

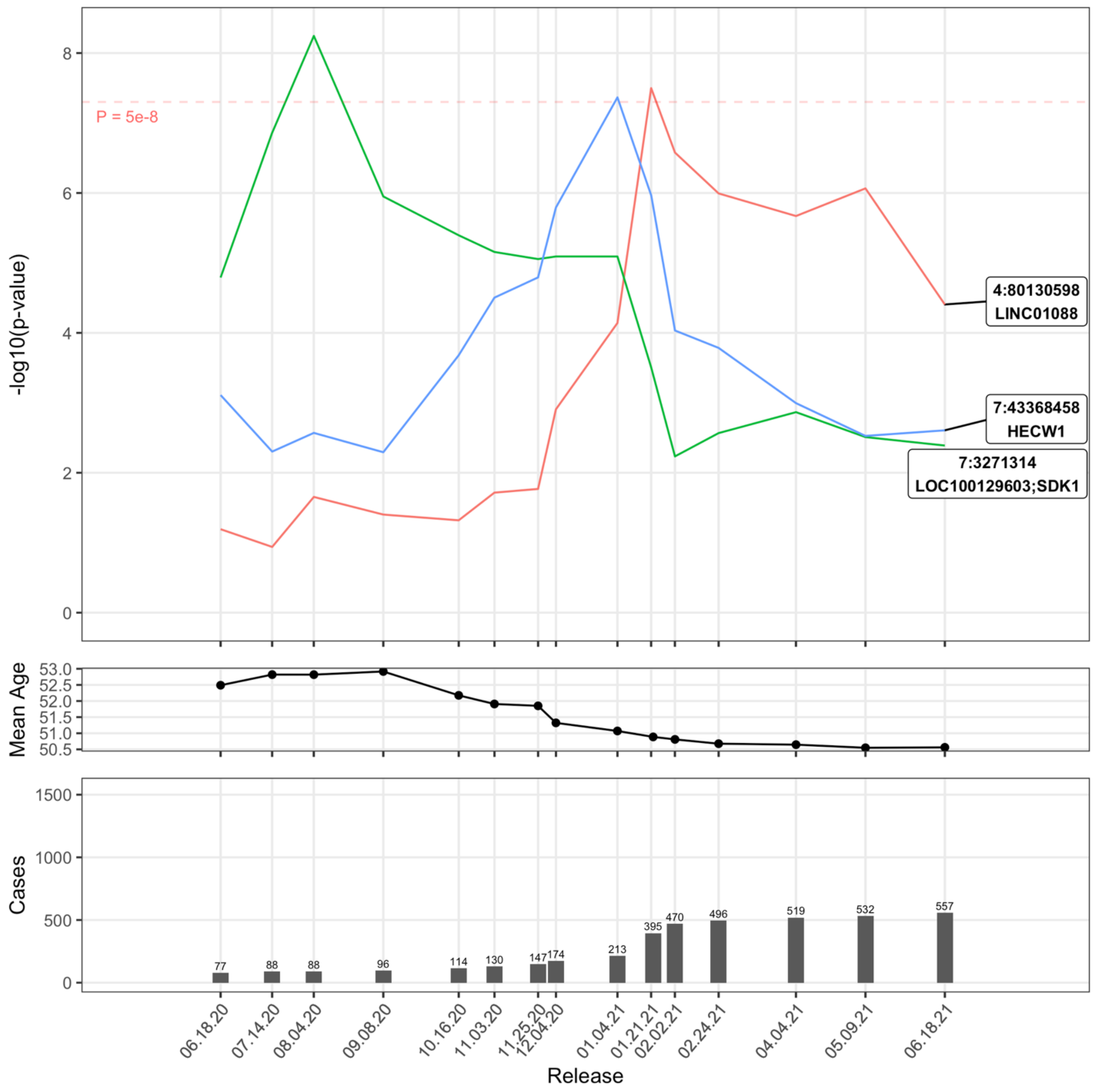

Supplemental Figure 15: Covid-19 Susceptibility in SAS:Pop

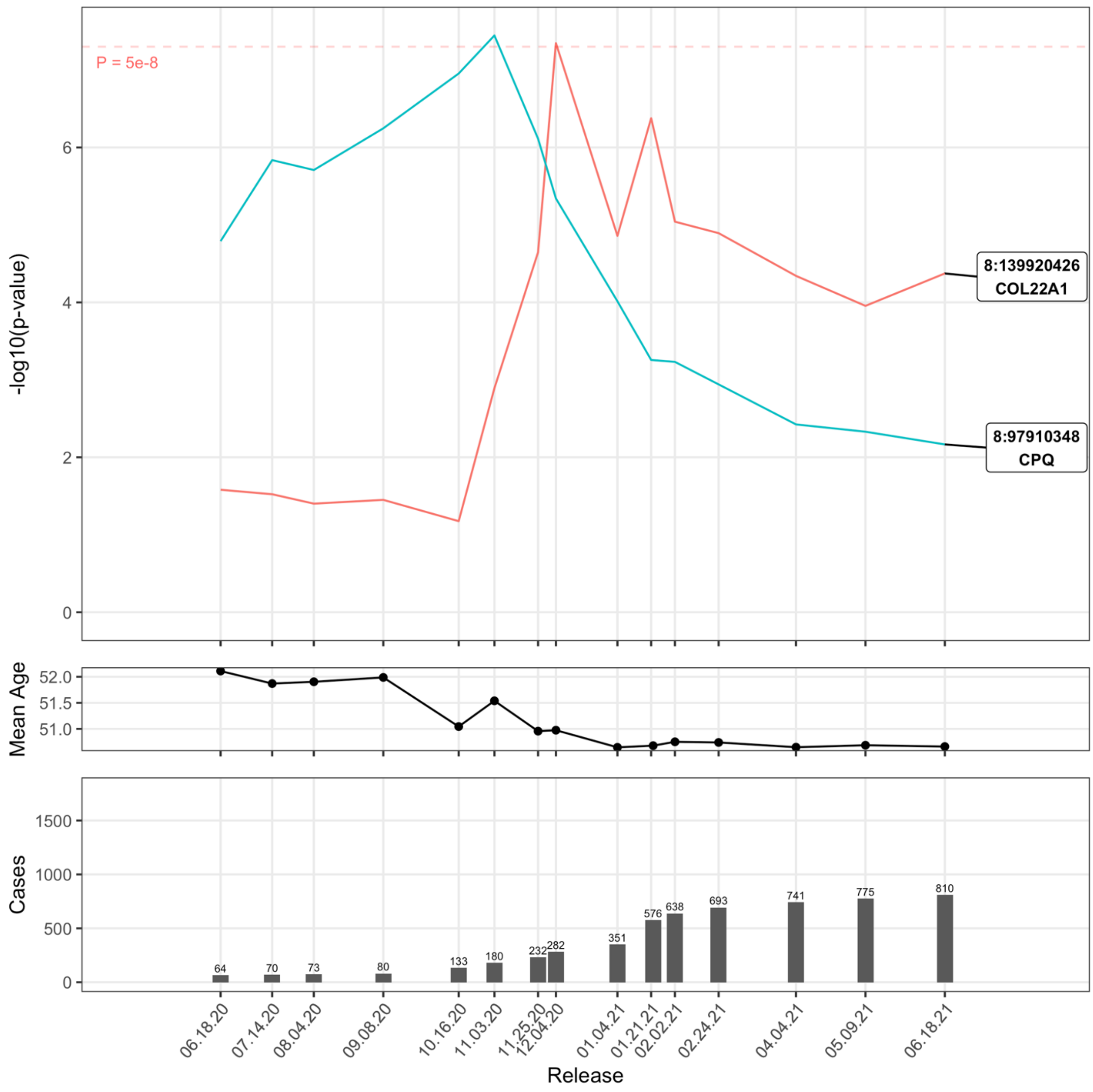

Supplemental Figure 16: Covid-19 Susceptibility in SAS:Tested

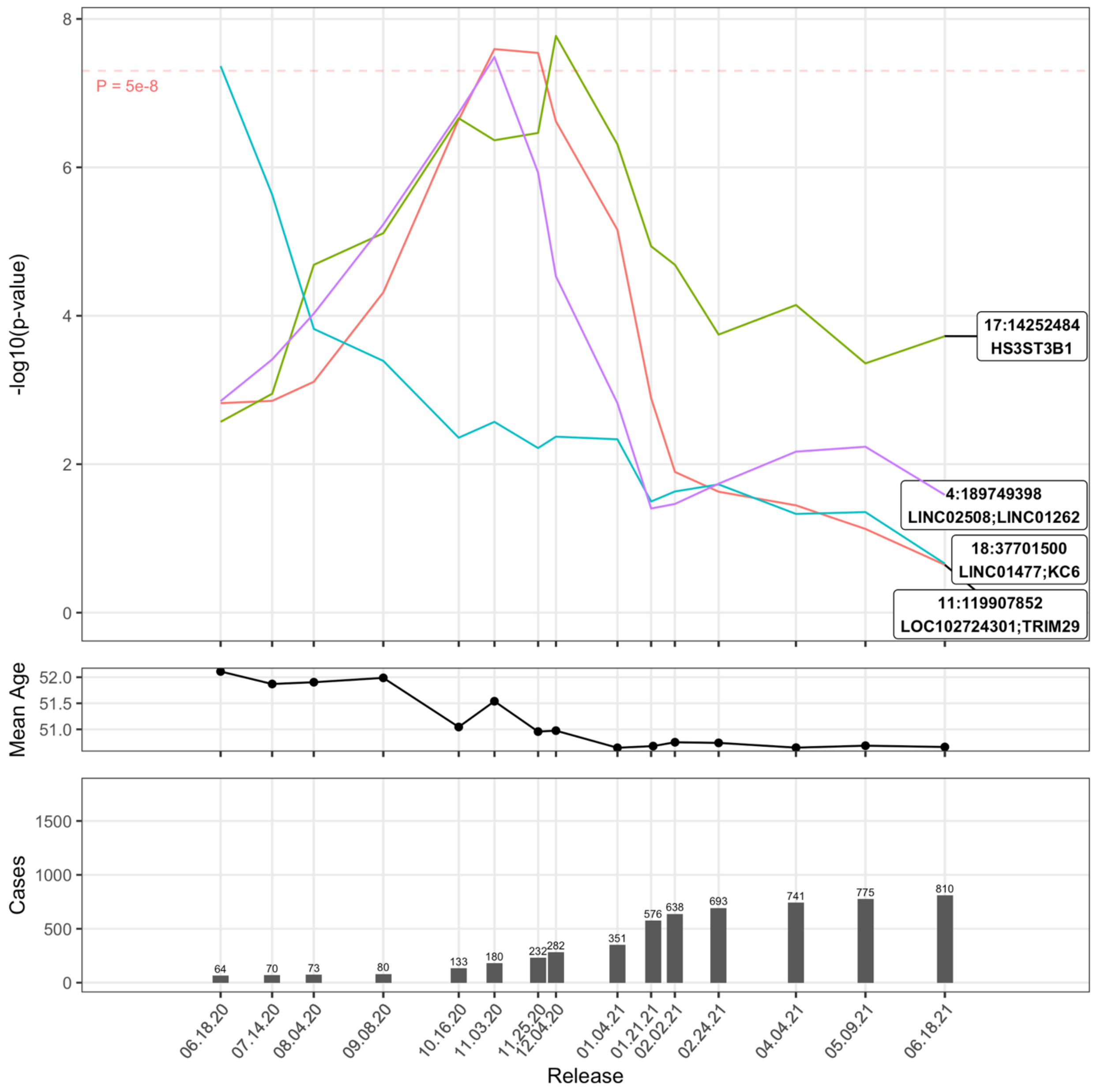

Supplemental Figure 17: Covid-19 Susceptibility in OTHERS:Tested

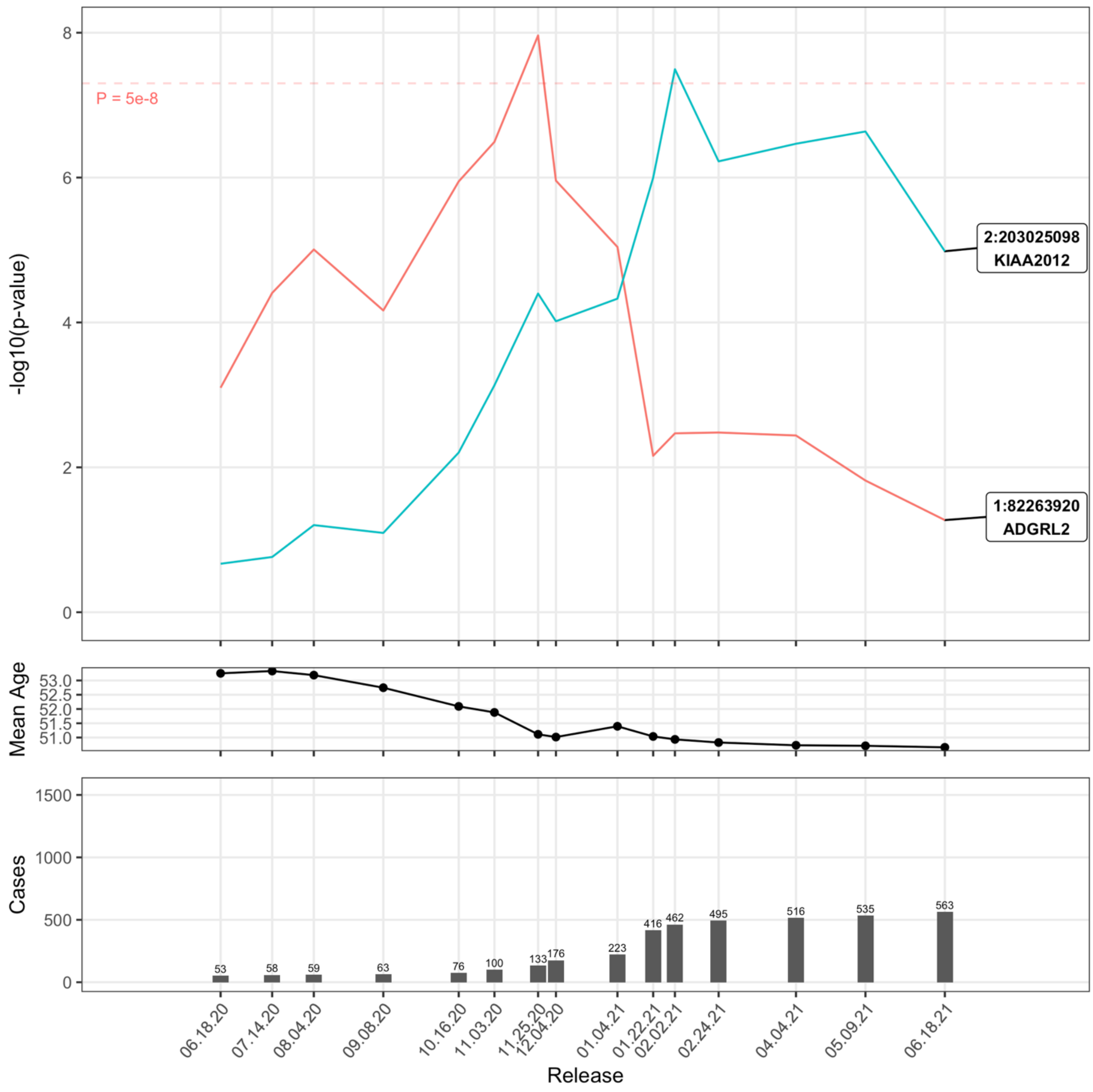

Supplemental Figure 18: Covid-19 Susceptibility in nEUR:Pop

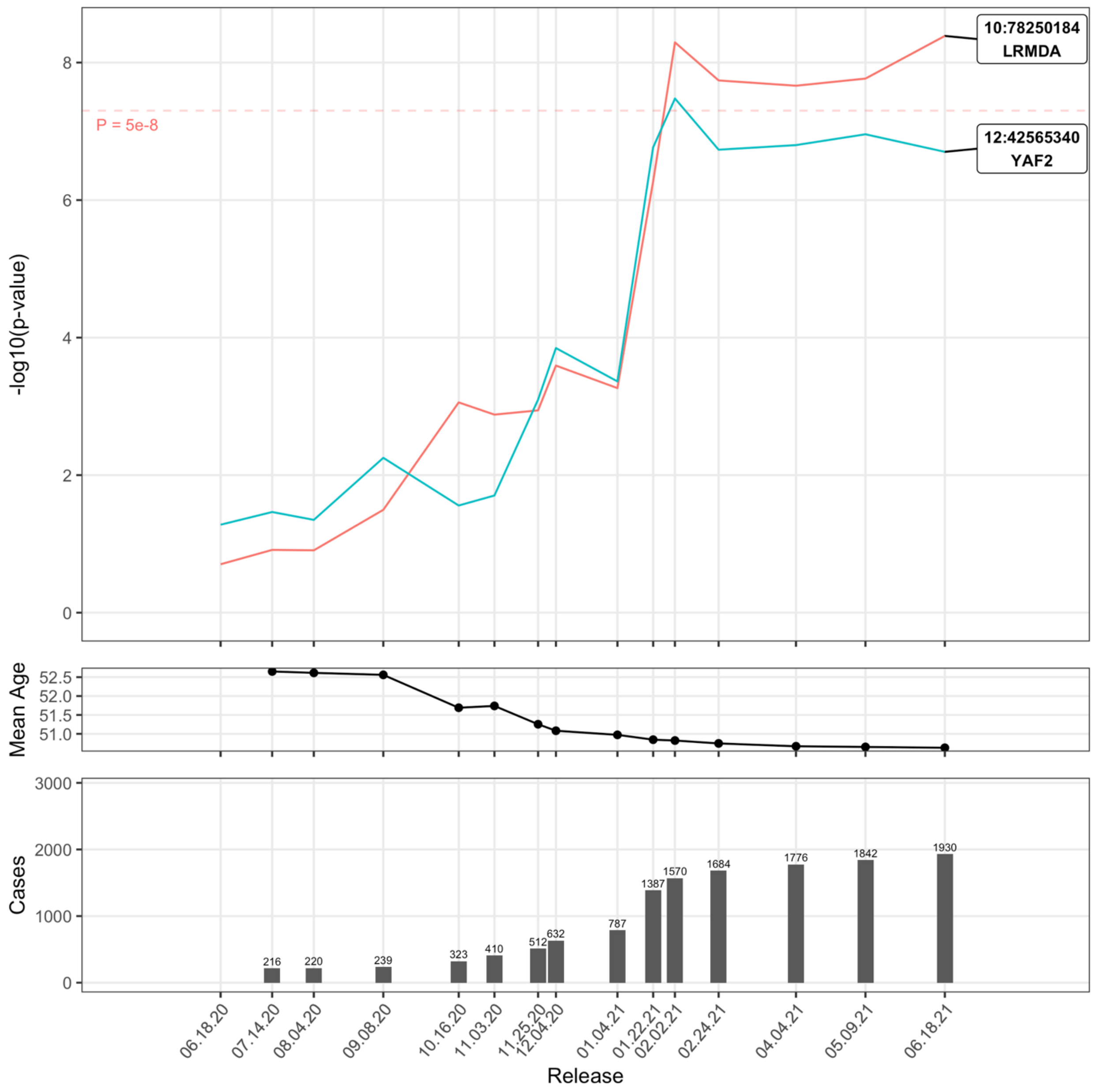

Supplemental Figure 19: Covid-19 Hospitalization in ALL:Pop

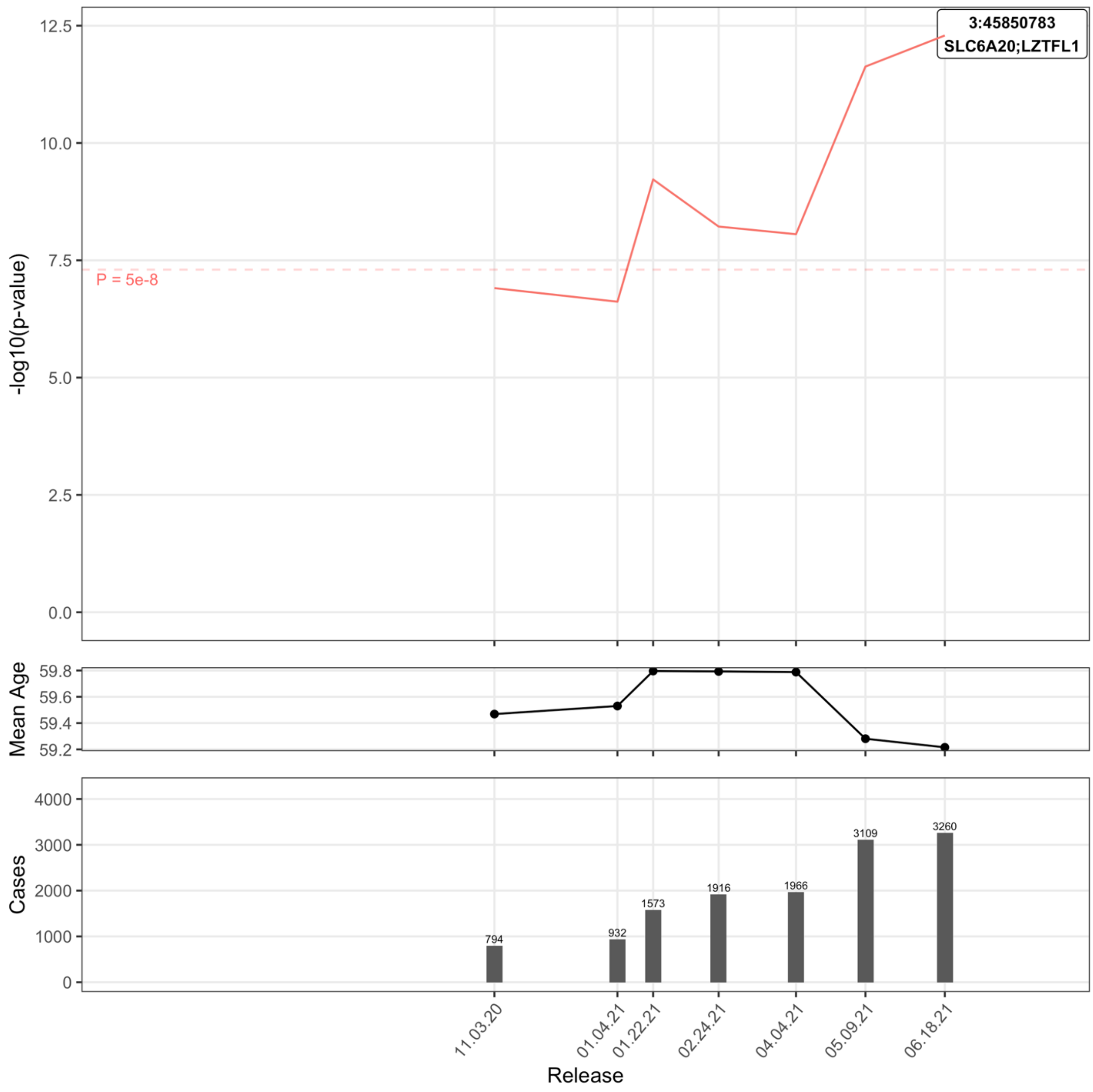

Supplemental Figure 20: Covid-19 Hospitalization in ALL:Pop:M

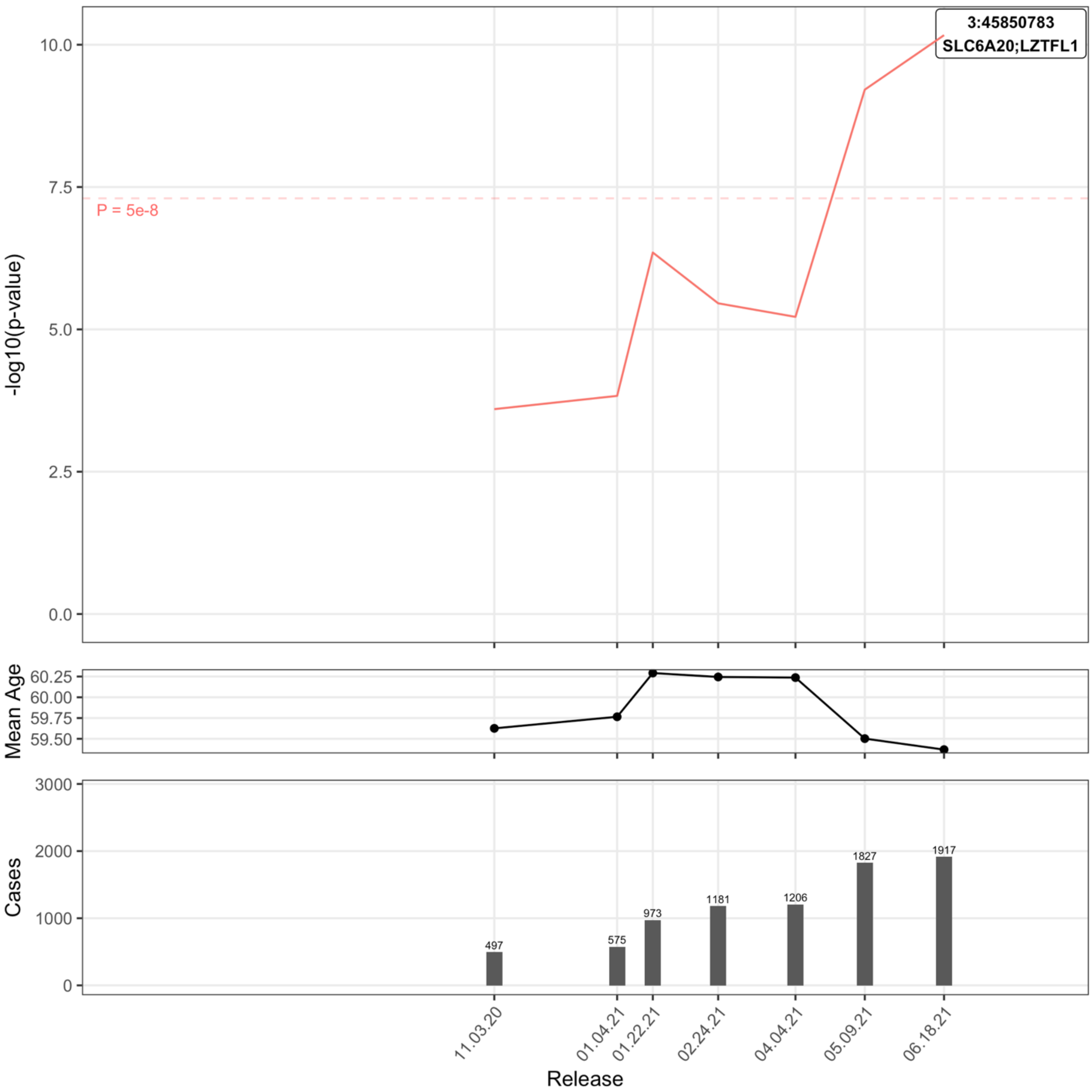

Supplemental Figure 21: Covid-19 Hospitalization in ALL:Tested

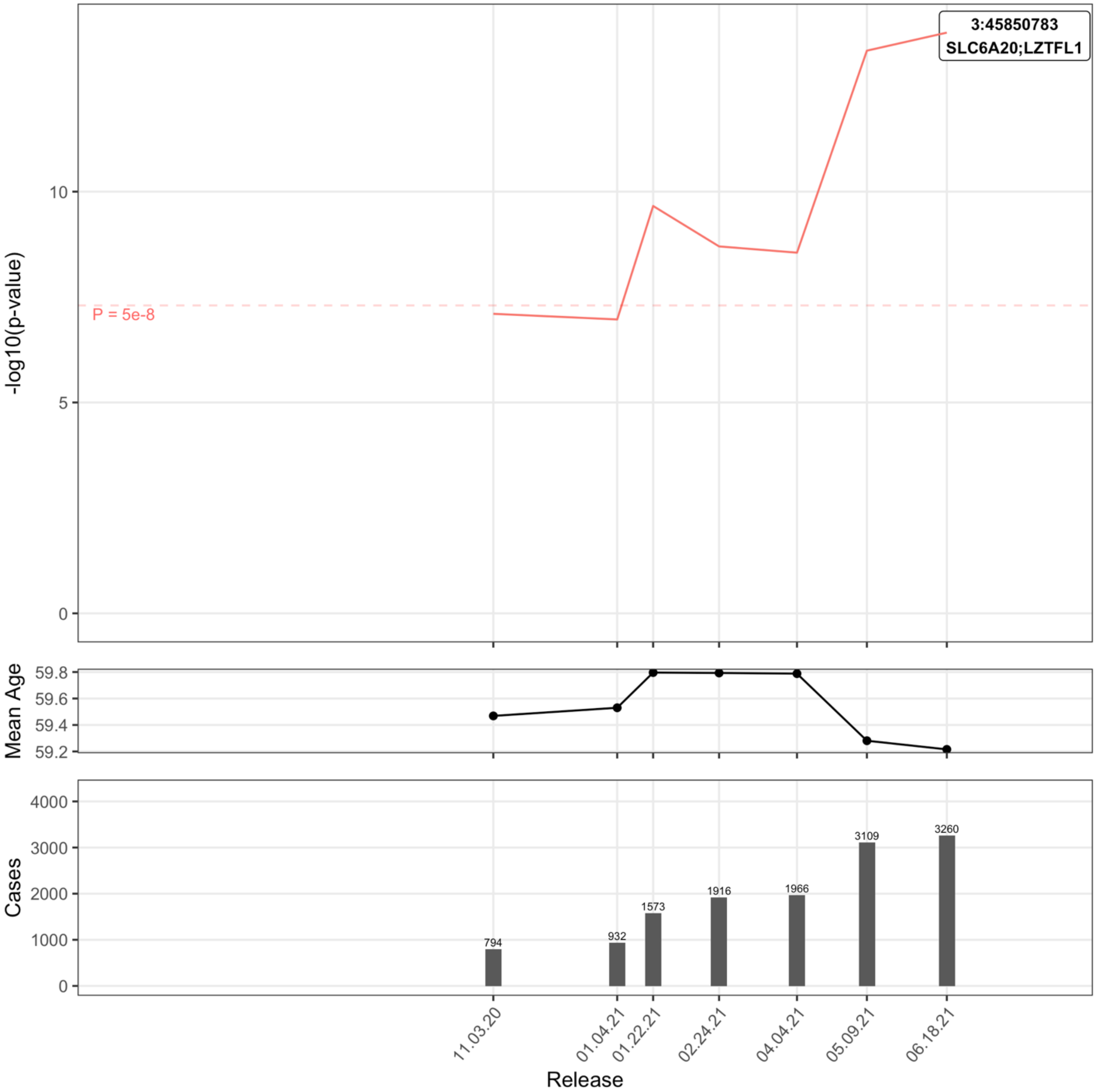

Supplemental Figure 22: Covid-19 Hospitalization in ALL:Tested:M

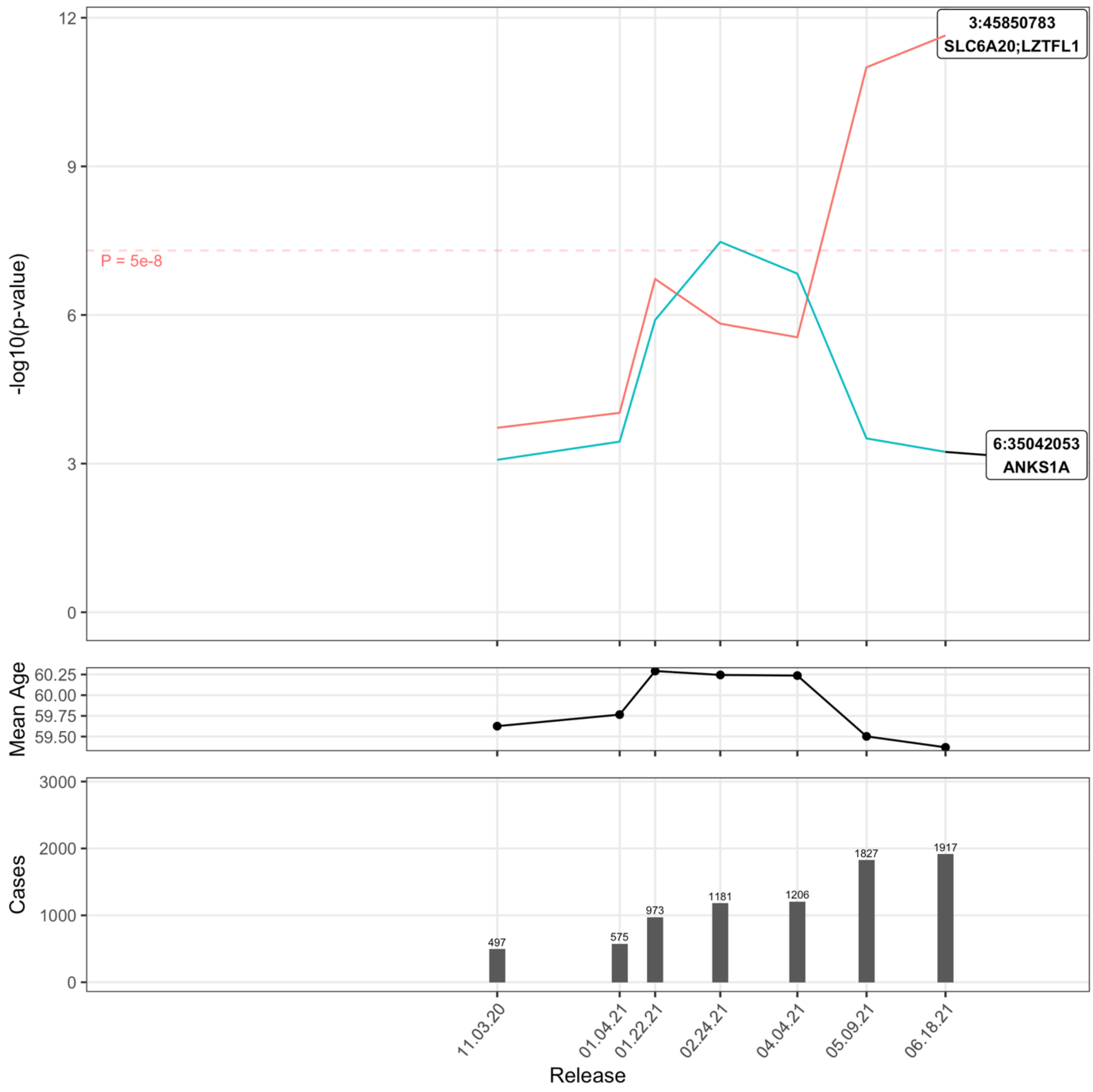

Supplemental Figure 23: Covid-19 Hospitalization in ALL:Positive

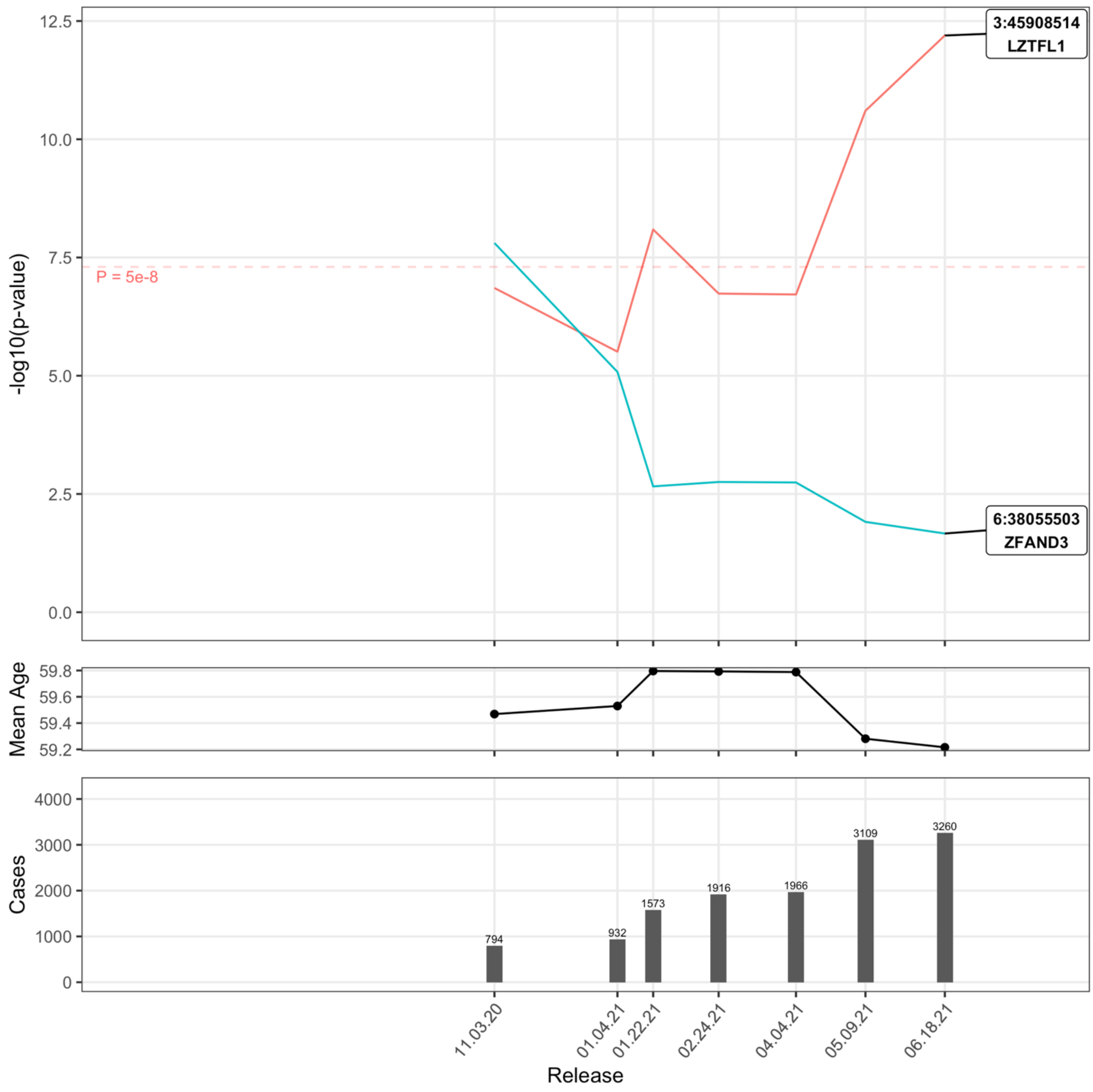

Supplemental Figure 24: Covid-19 Hospitalization in ALL:Positive:M

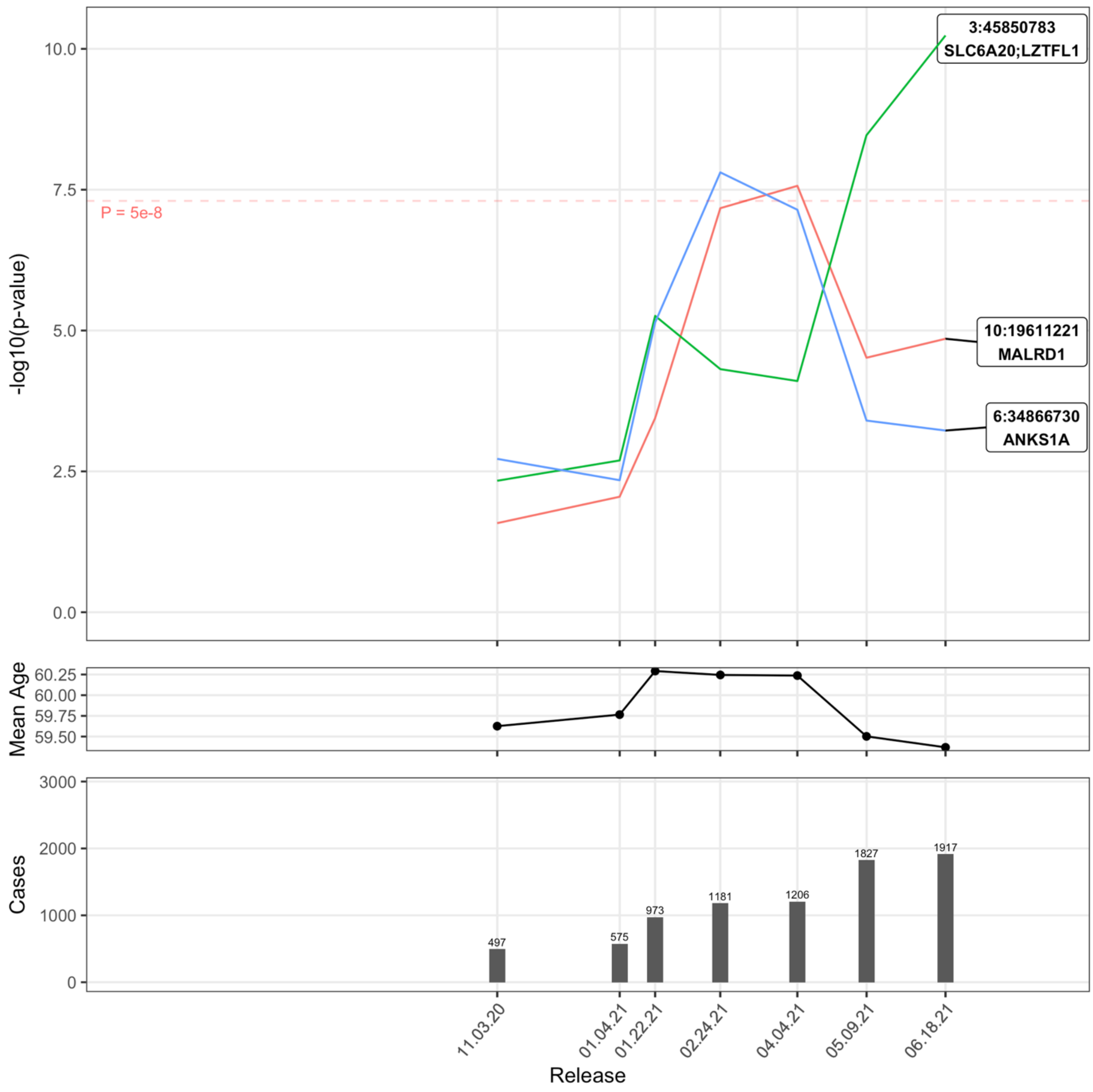

Supplemental Figure 25: Covid-19 Hospitalization in EUR:Pop

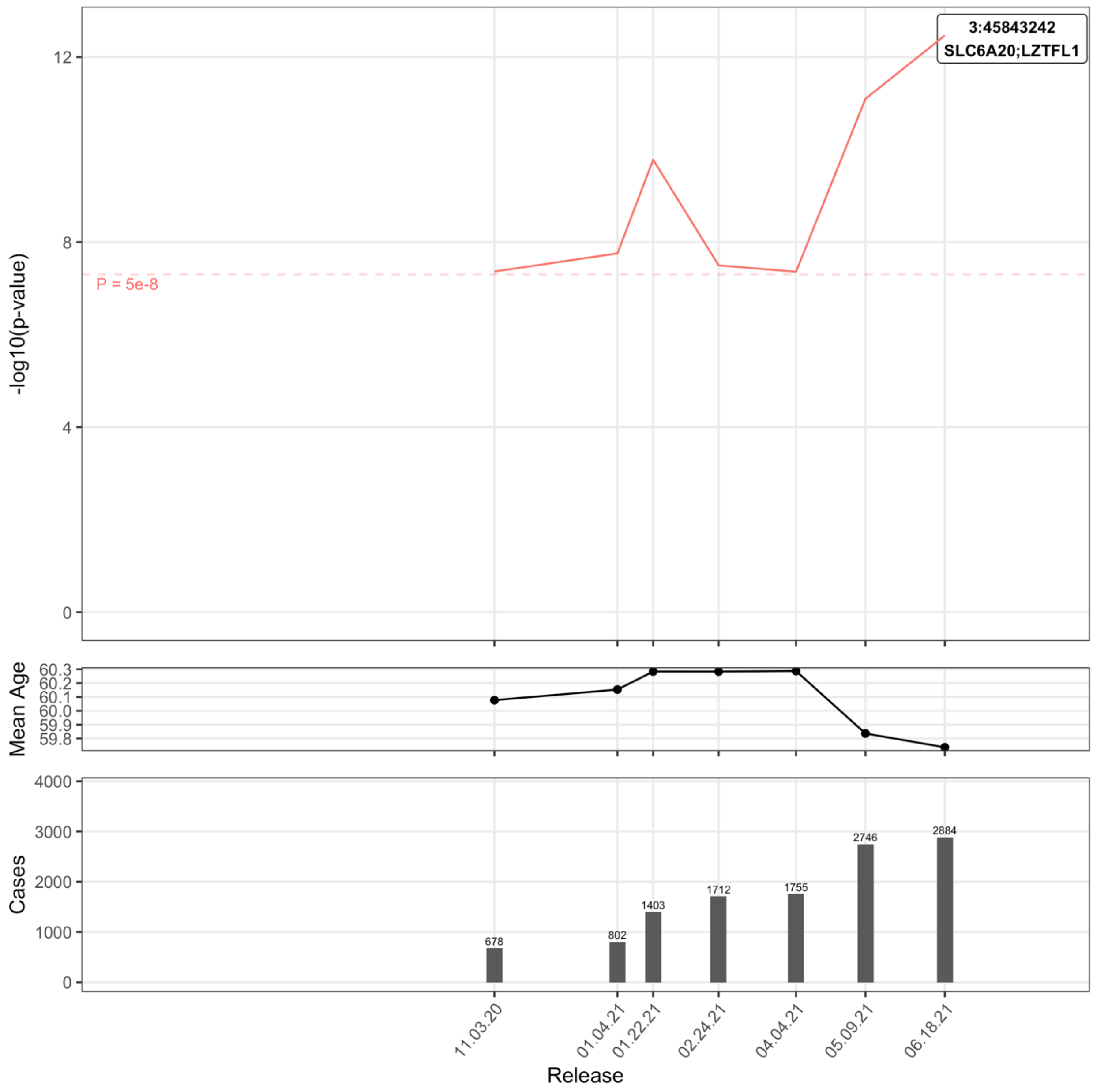

Supplemental Figure 26: Covid-19 Hospitalization in EUR:Pop:M

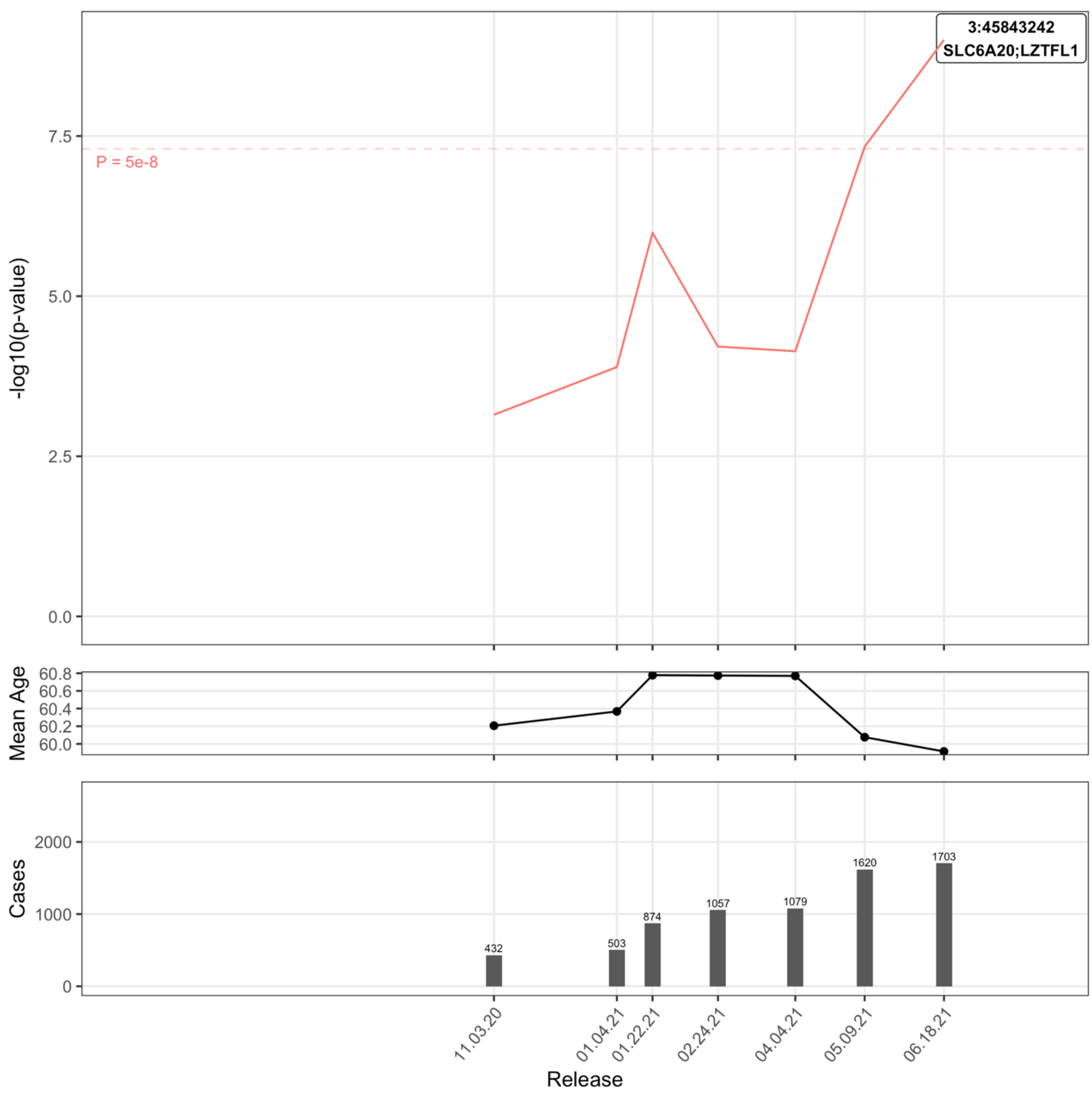

Supplemental Figure 27: Covid-19 Hospitalization in EUR:Tested

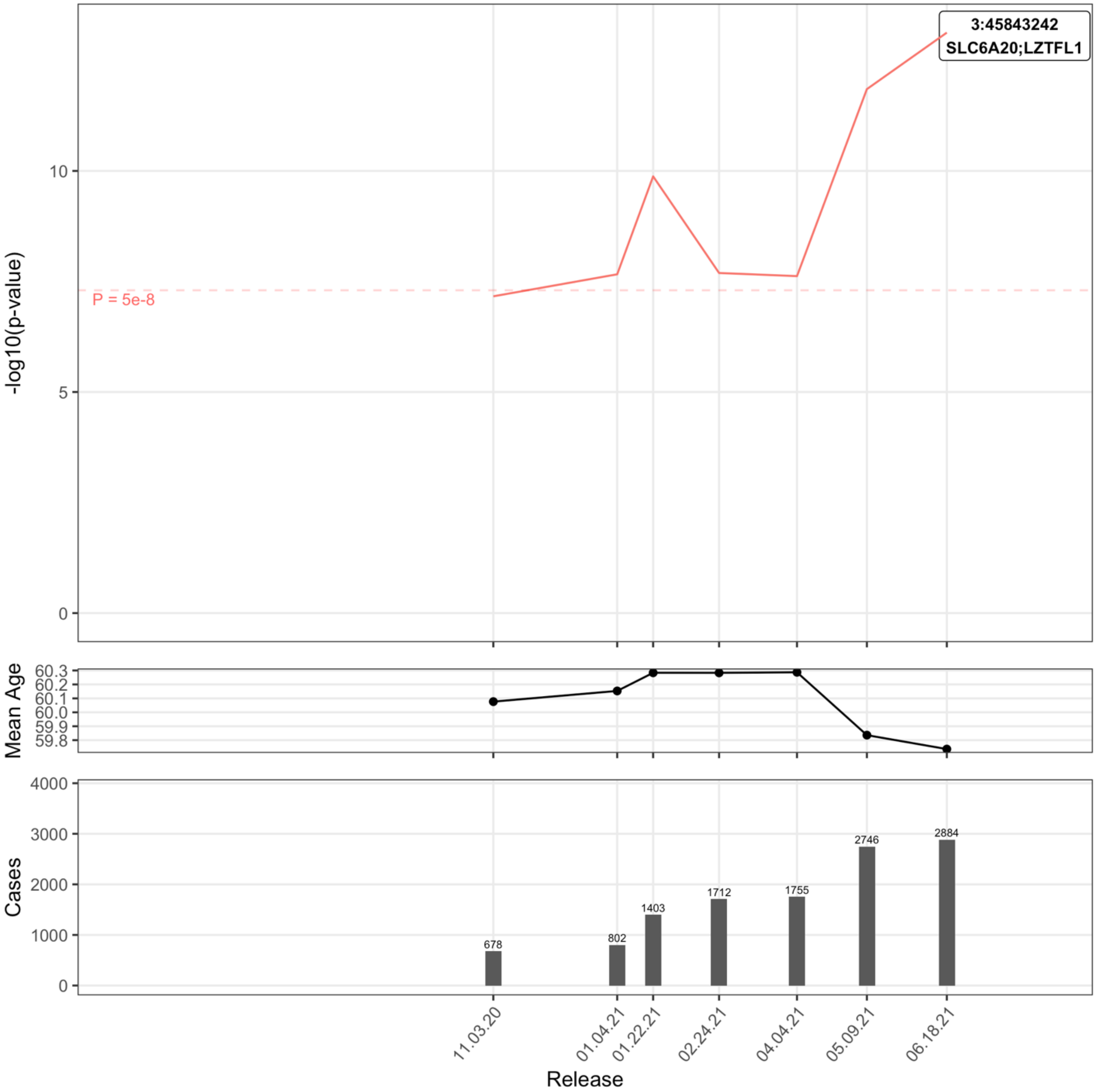

Supplemental Figure 28: Covid-19 Hospitalization in EUR:Tested:M

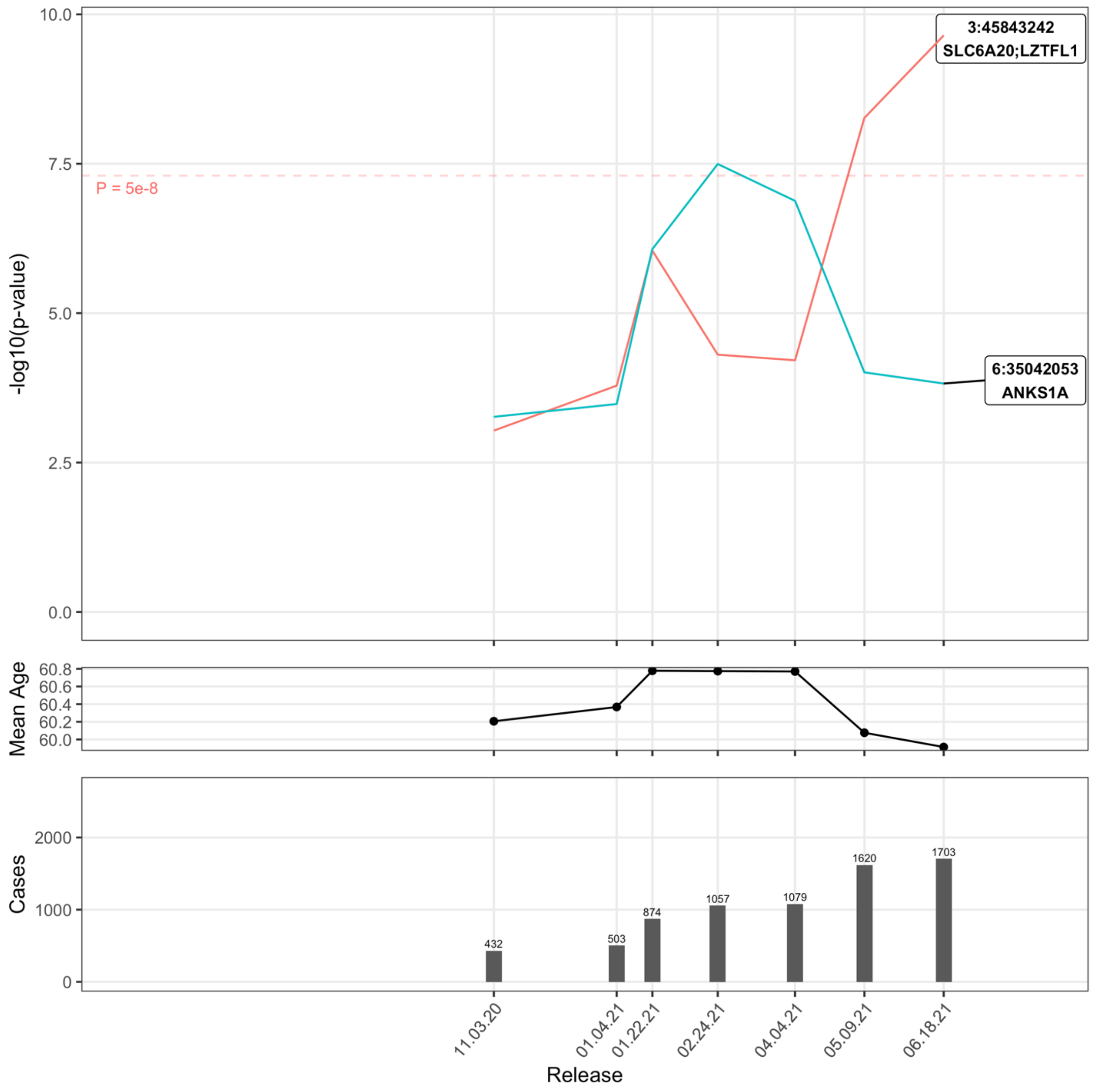

Supplemental Figure 29: Covid-19 Hospitalization in EUR:Positive

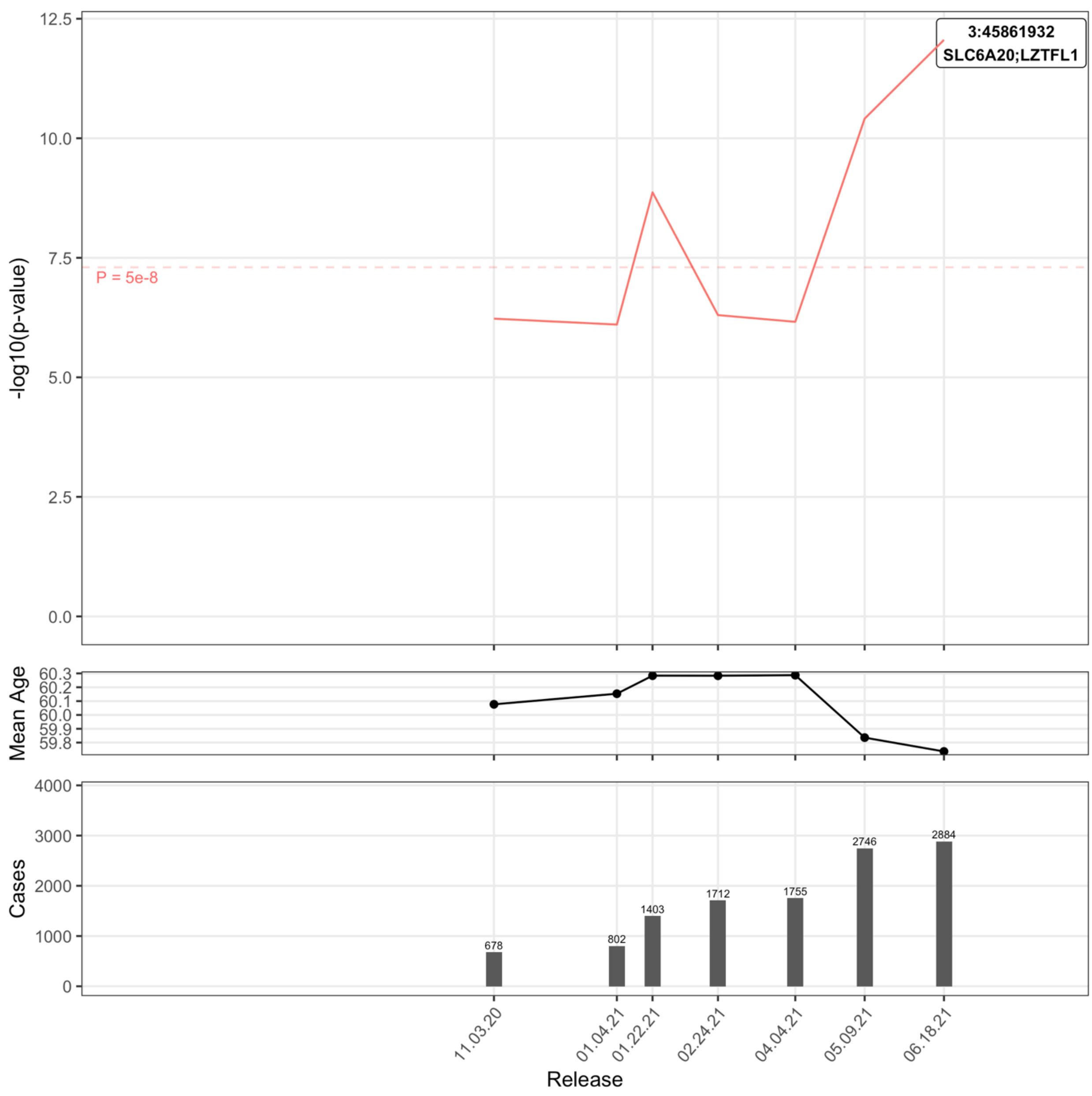

Supplemental Figure 30: Covid-19 Hospitalization in EUR:Positive:F

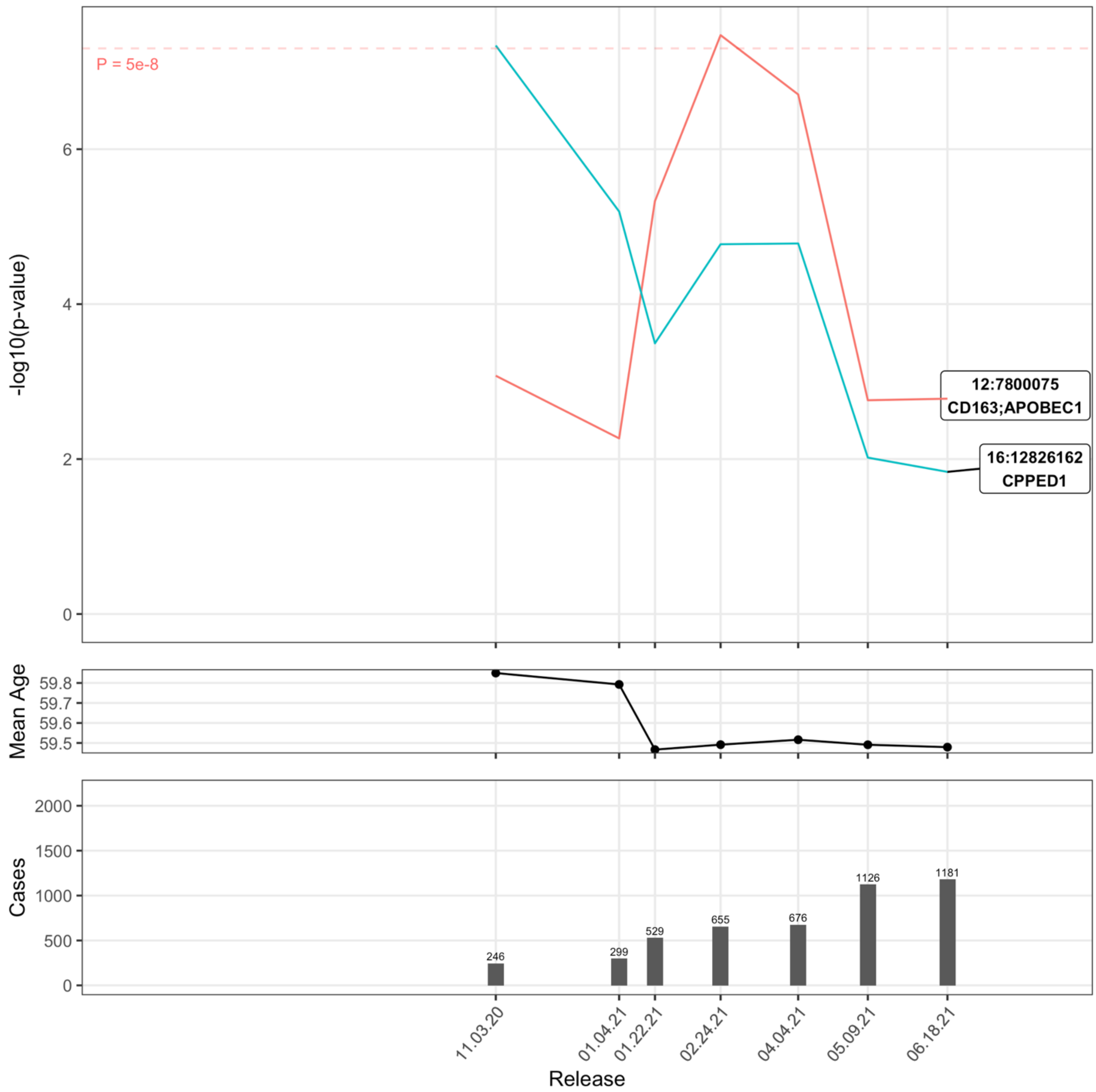

Supplemental Figure 31: Covid-19 Hospitalization in EUR:Positive:M

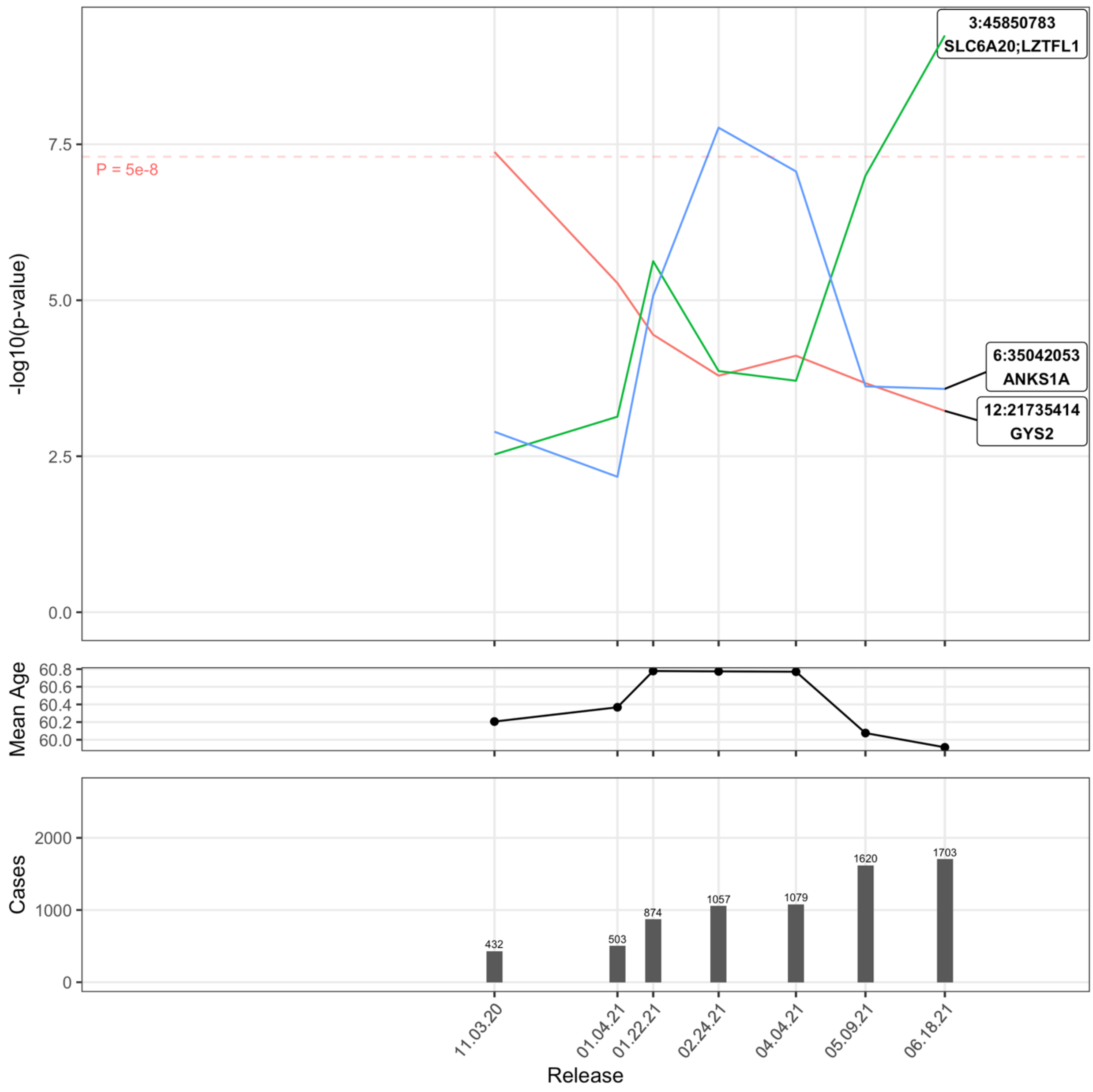

Supplemental Figure 32: Severe Covid-19 in ALL:Pop

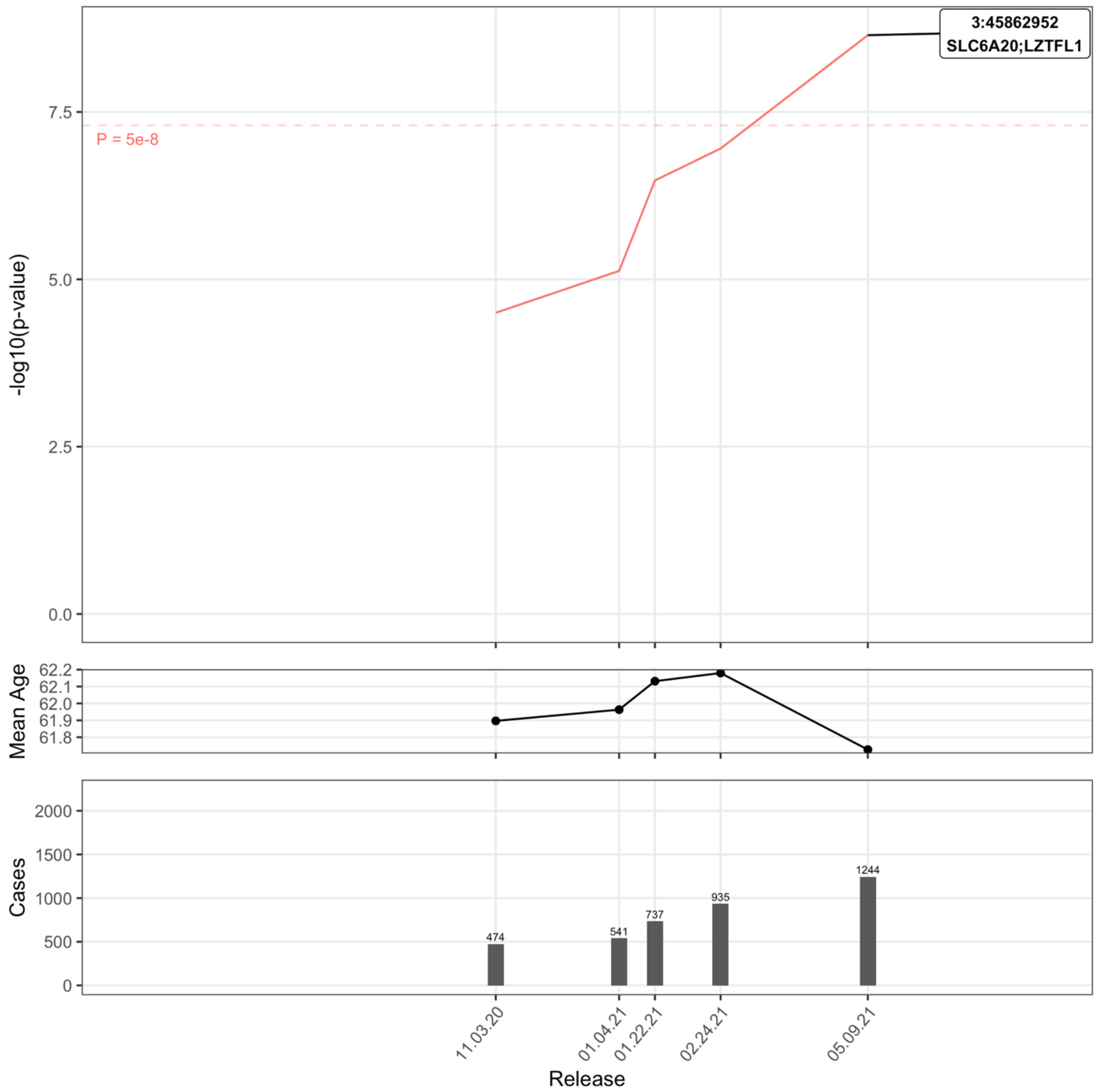

Supplemental Figure 33: Severe Covid-19 in ALL:Pop:M

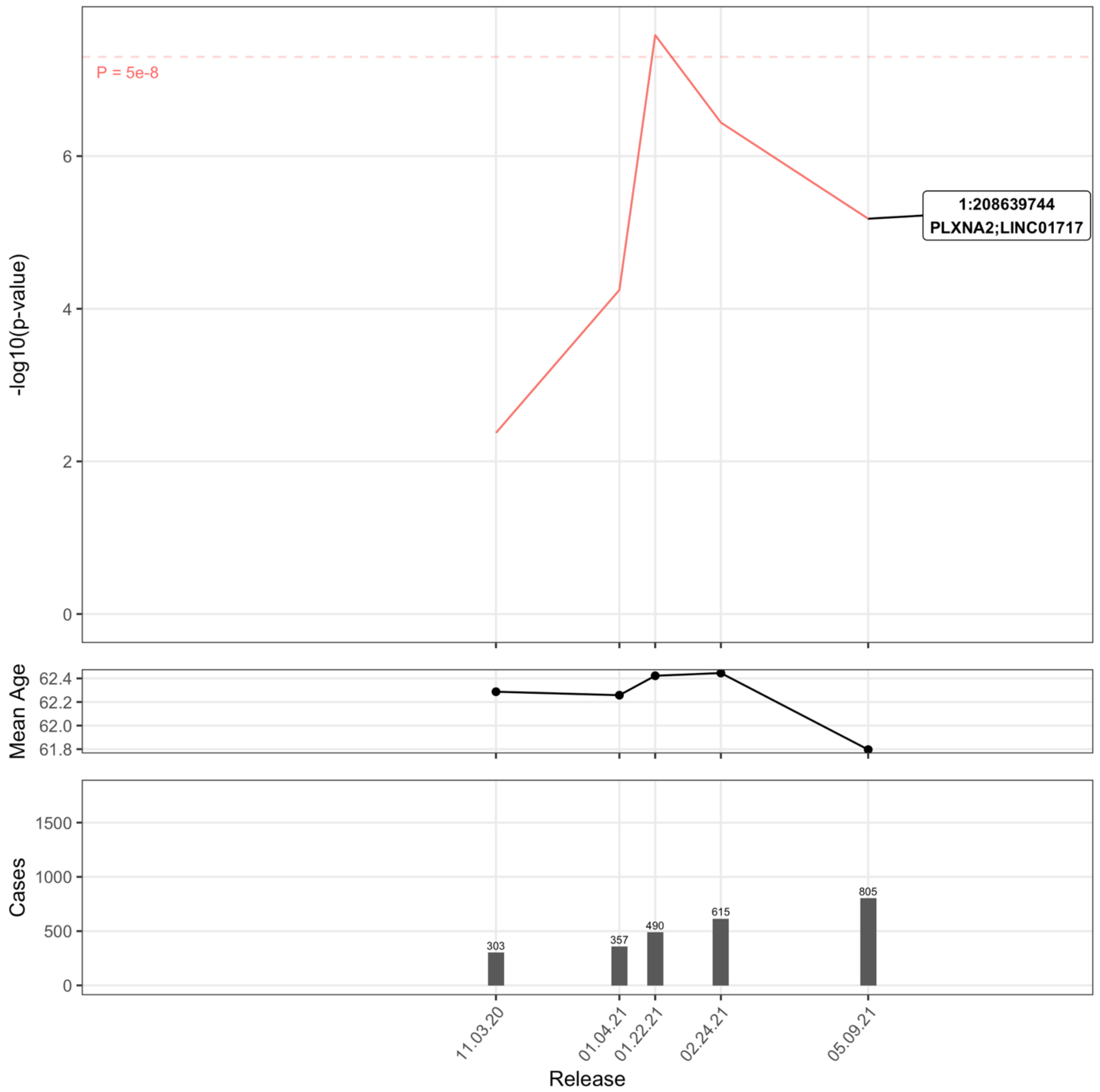

Supplemental Figure 34: Severe Covid-19 in ALL:Tested

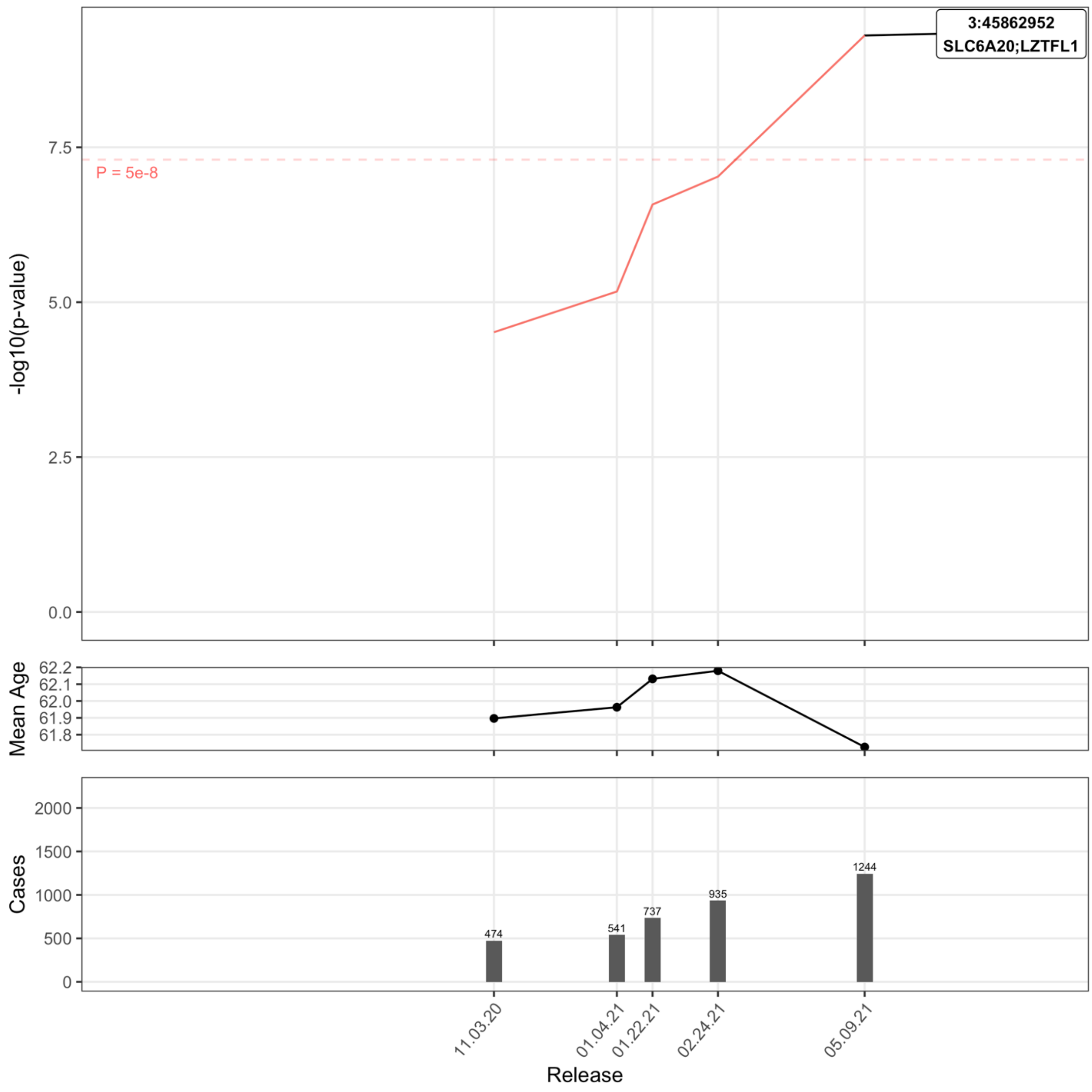

Supplemental Figure 35: Severe Covid-19 in ALL:Tested:M

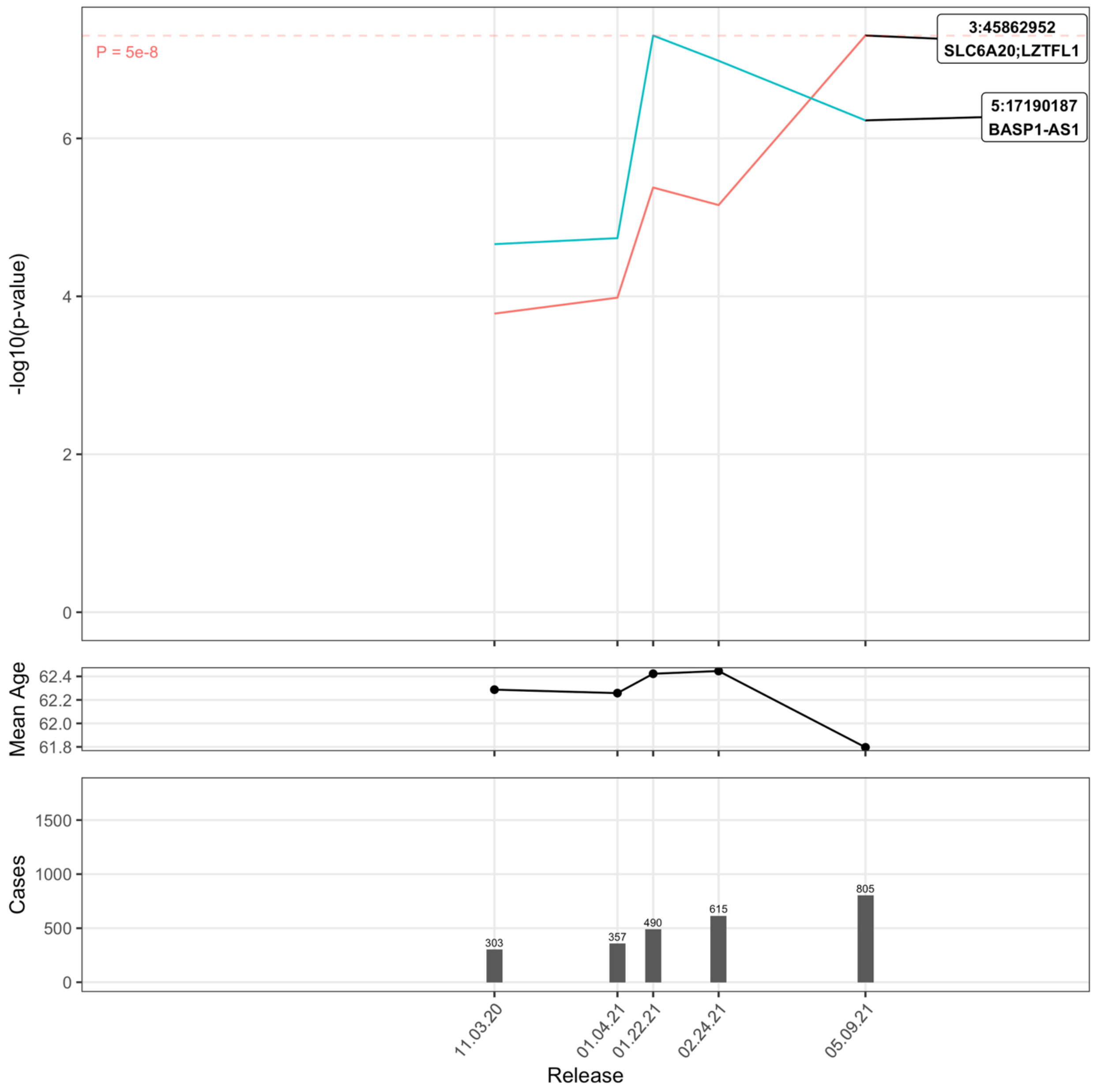

Supplemental Figure 36: Severe Covid-19 in ALL:Positive

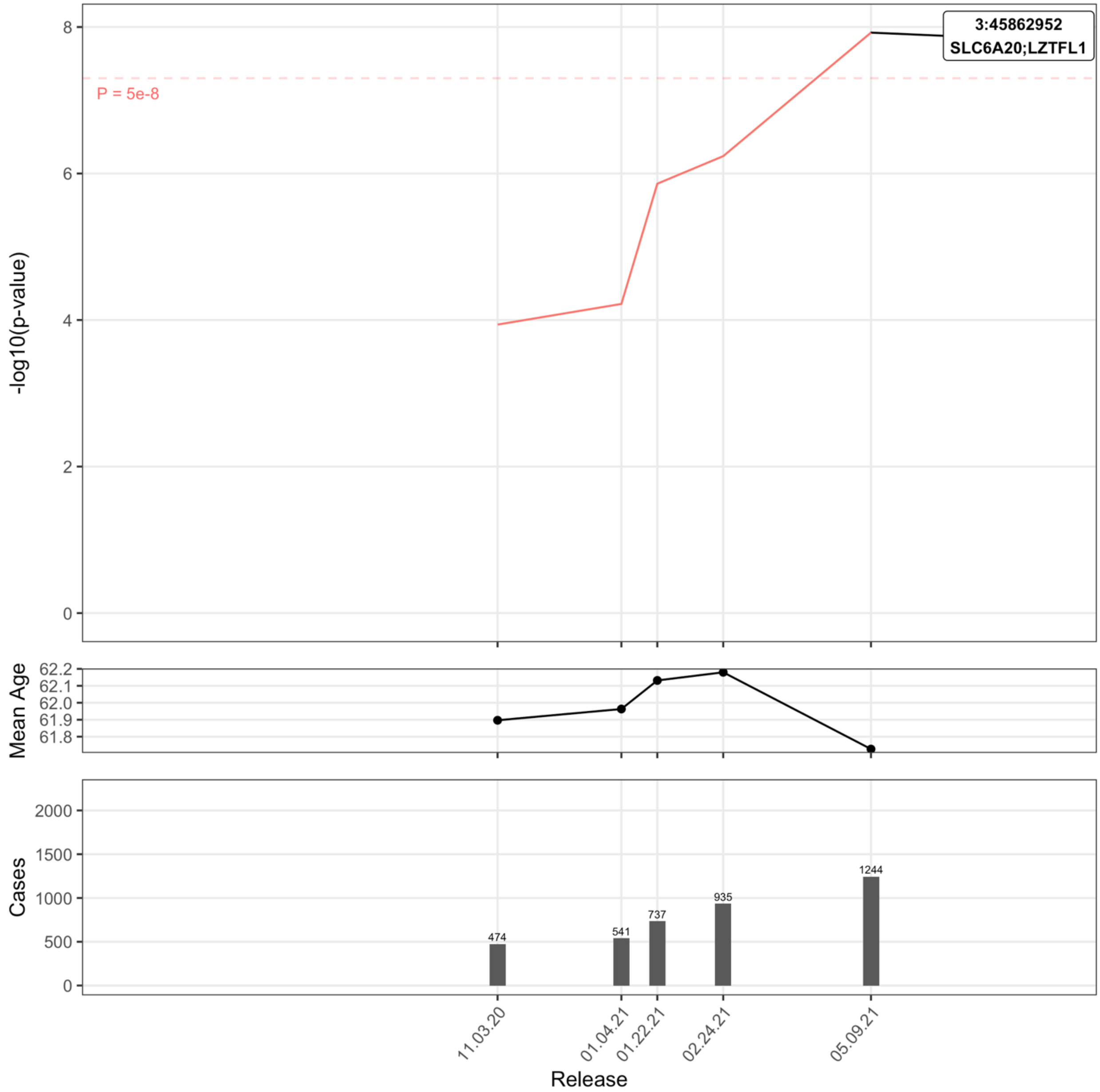

Supplemental Figure 37: Severe Covid-19 in EUR:Pop

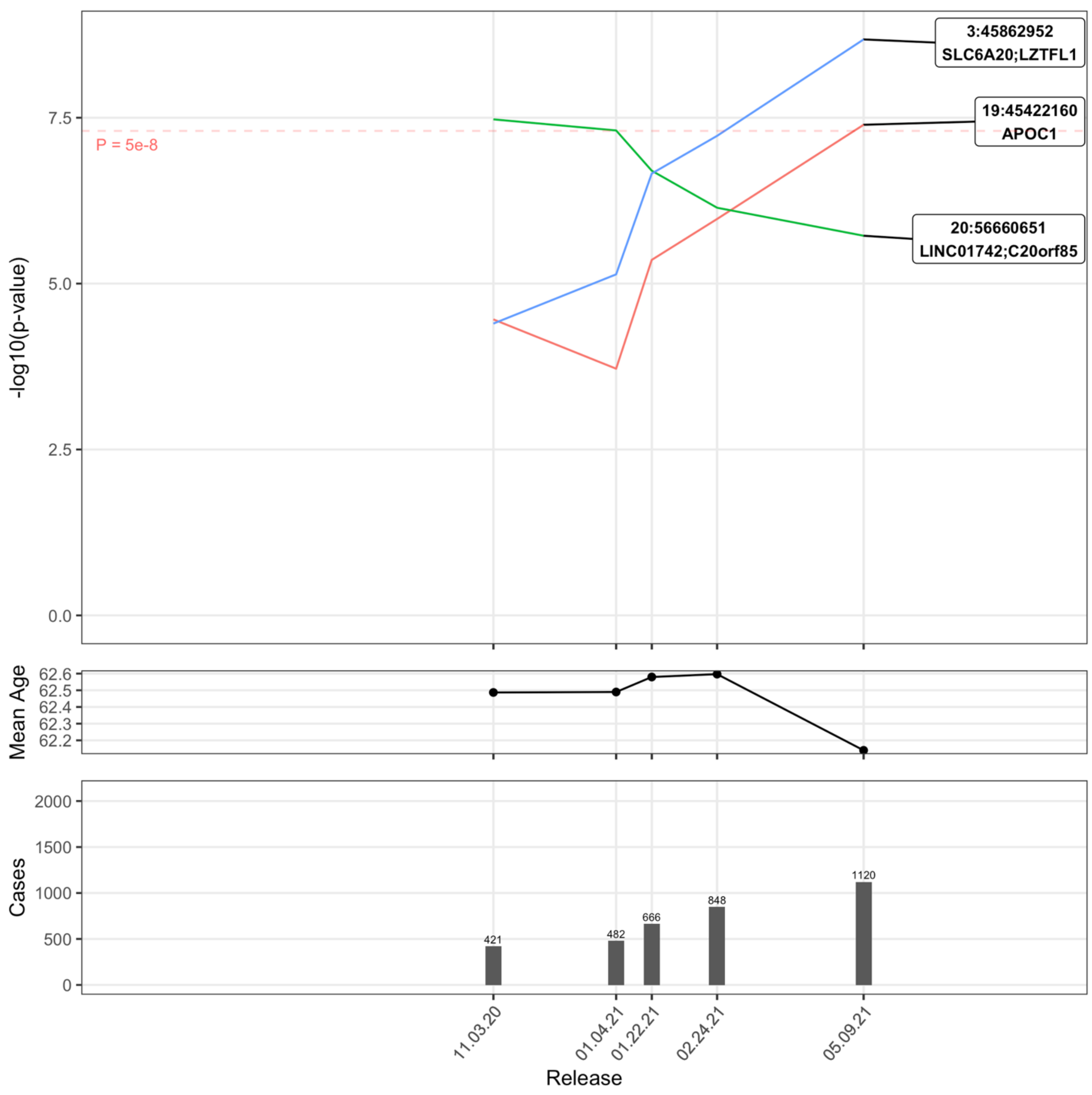

Supplemental Figure 38: Severe Covid-19 in EUR:Pop:F

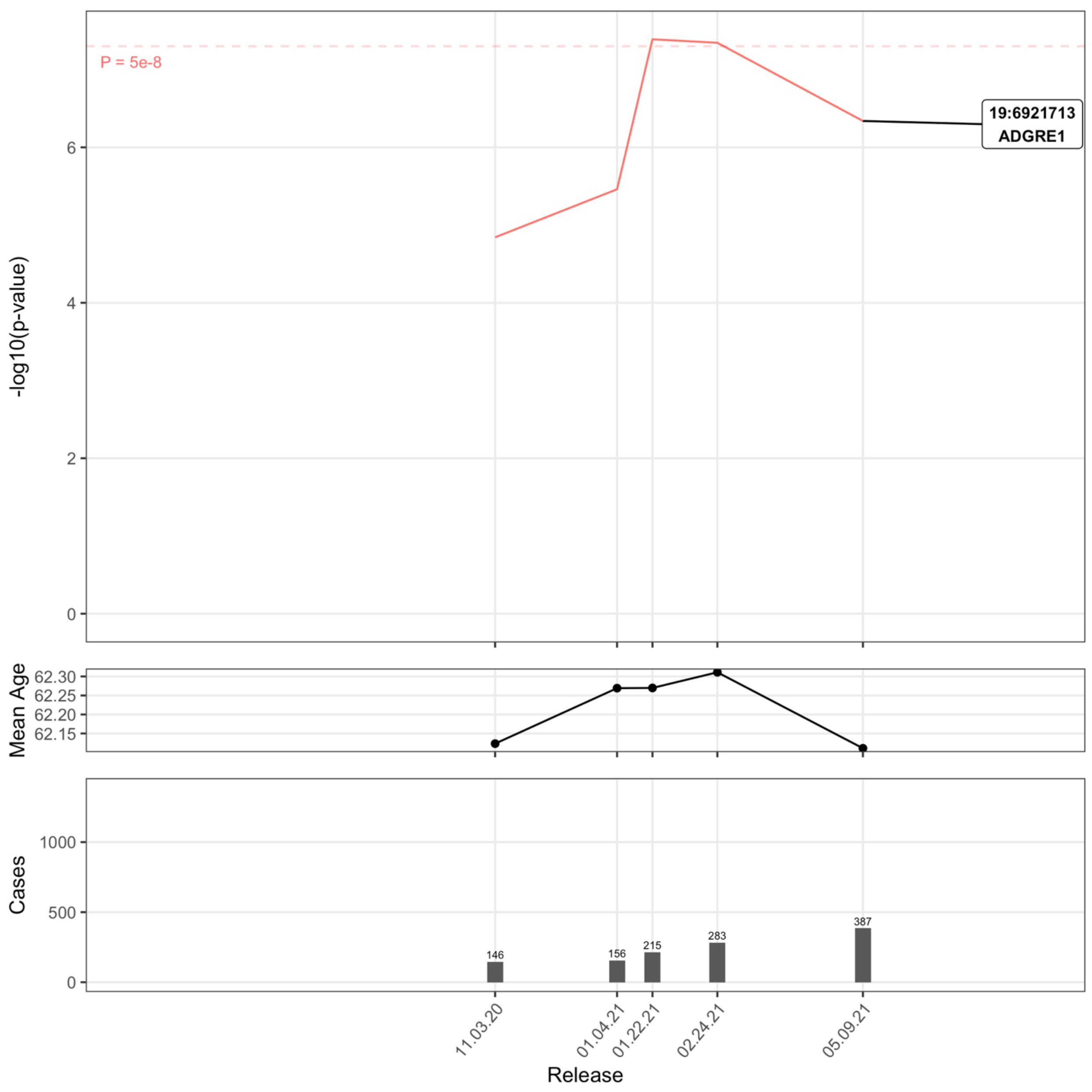

Supplemental Figure 39: Severe Covid-19 in EUR:Tested

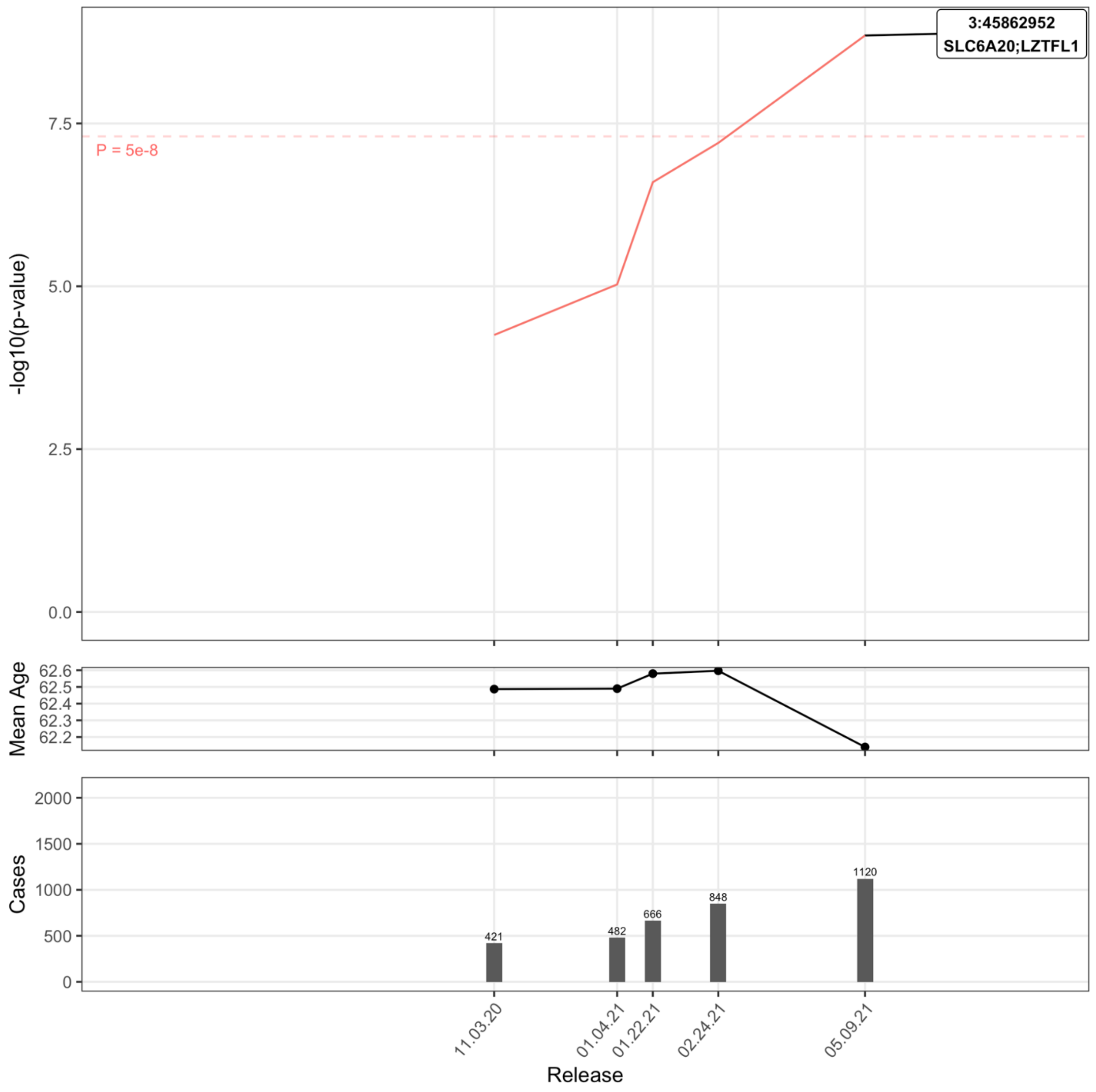

Supplemental Figure 40: Severe Covid-19 in EUR:Tested:F

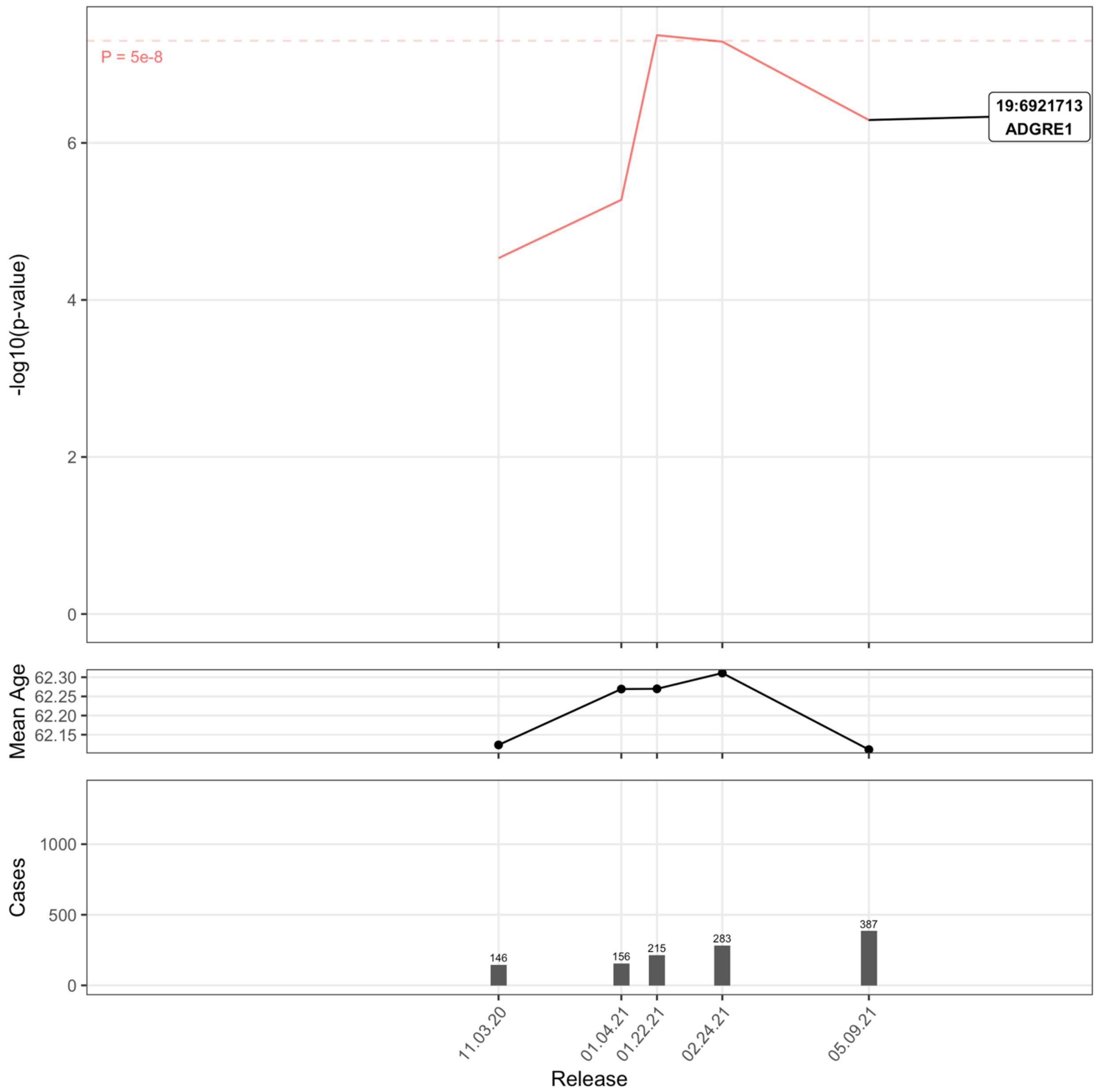

Supplemental Figure 41: Severe Covid-19 in EUR:Positive

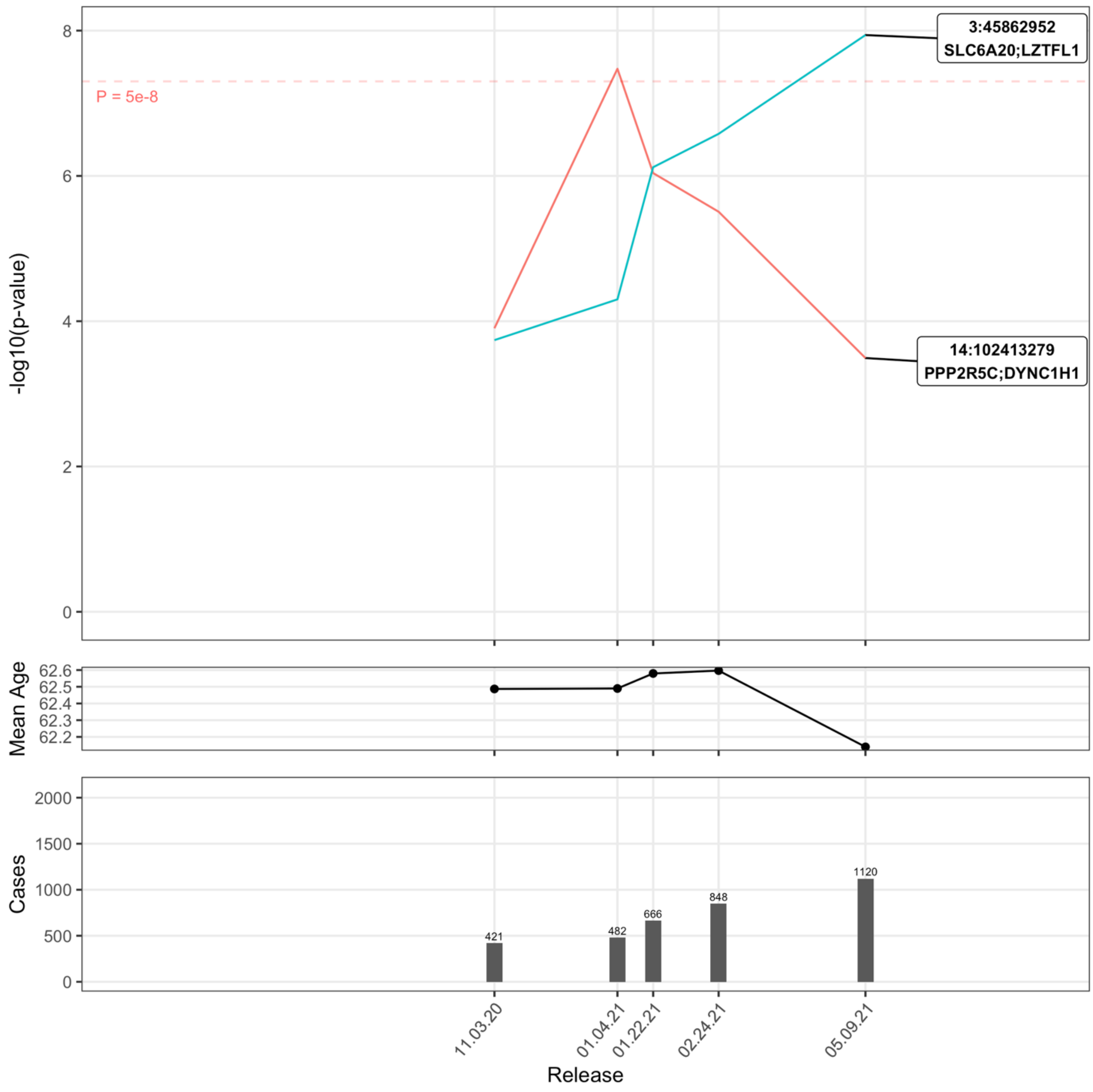

Supplemental Figure 42: Severe Covid-19 in EUR:Positive:M

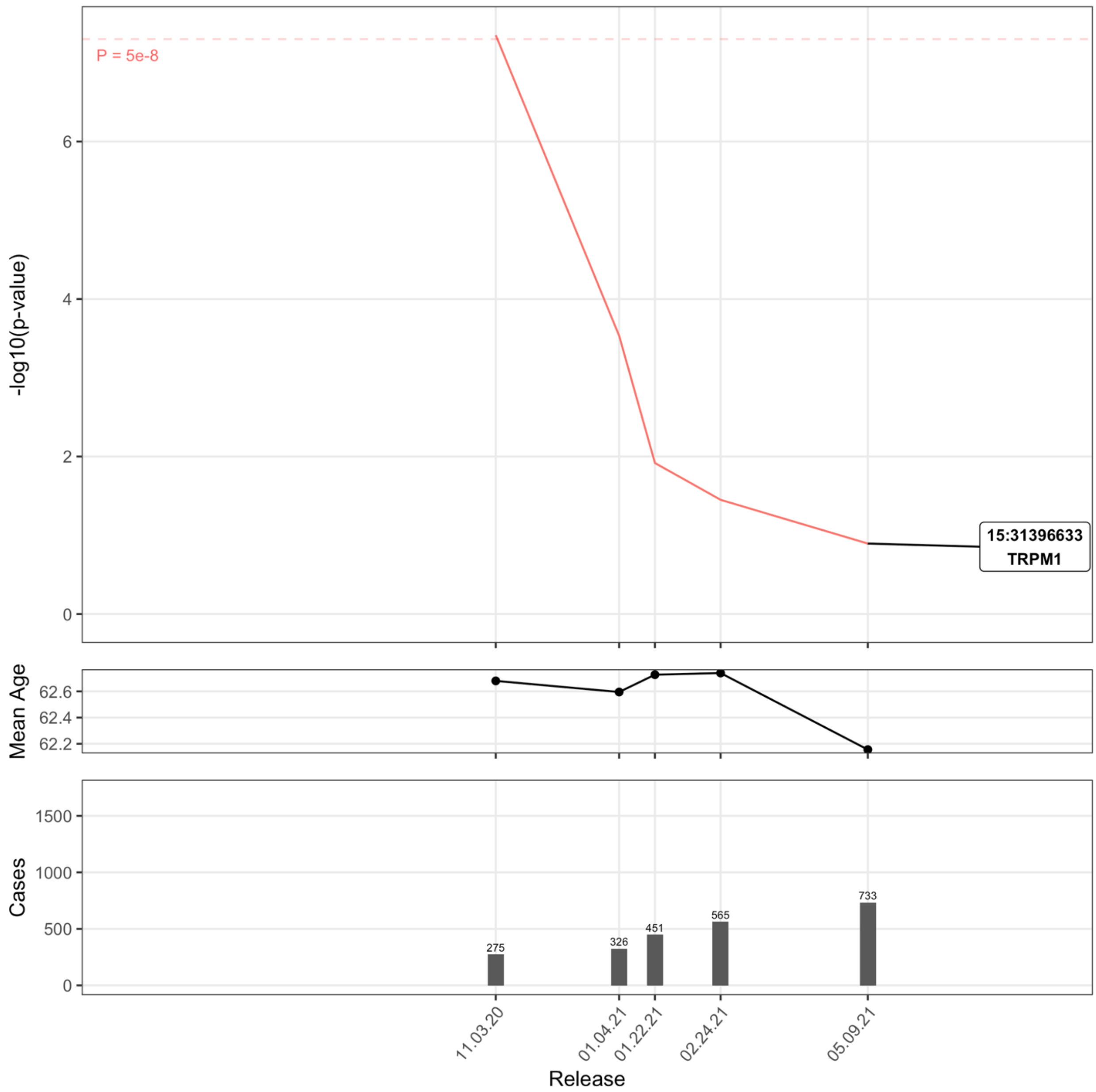

Supplemental Figure 43: Covid-19 Death in ALL:Pop

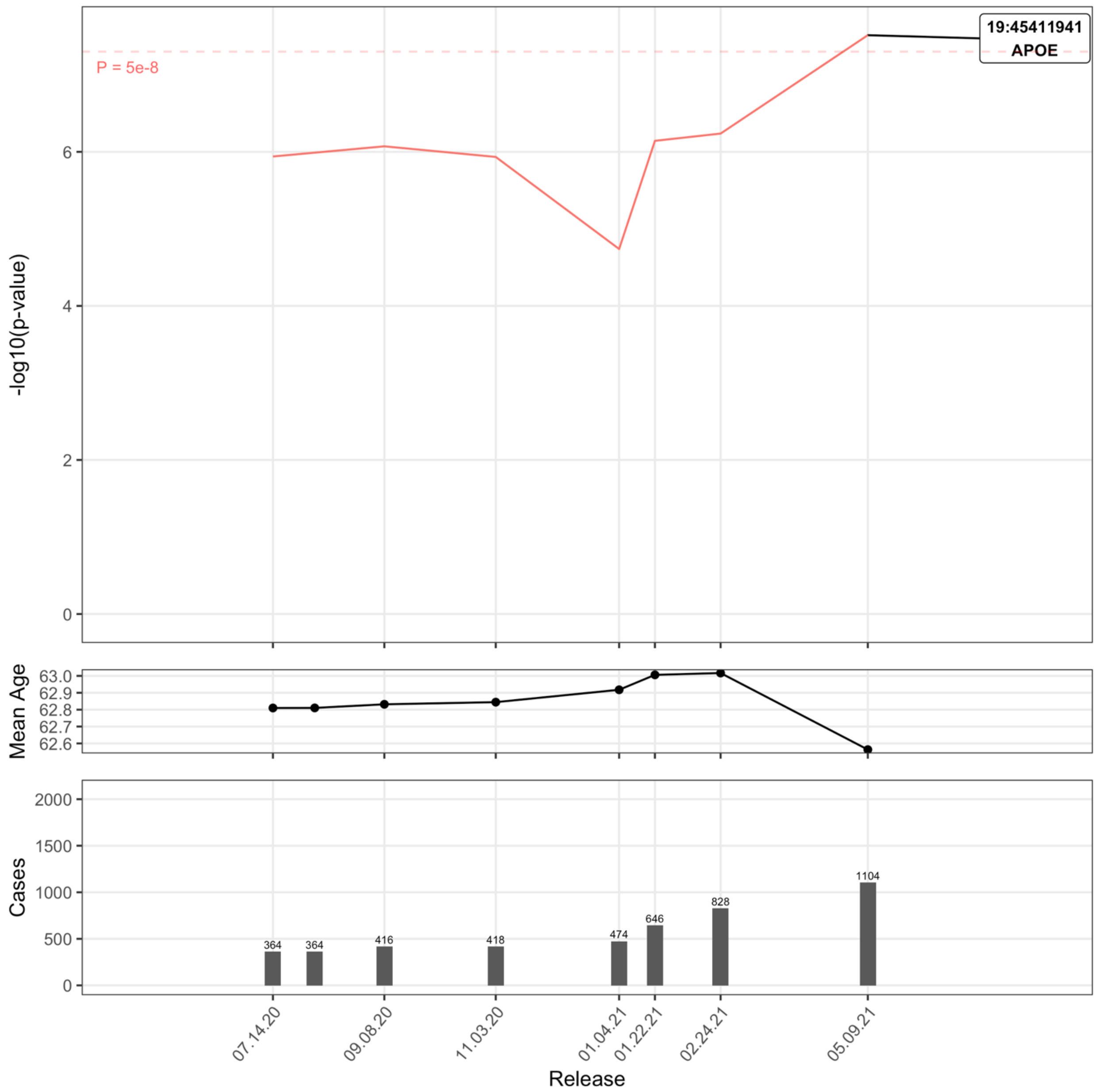

Supplemental Figure 44: Covid-19 Death in ALL:Tested

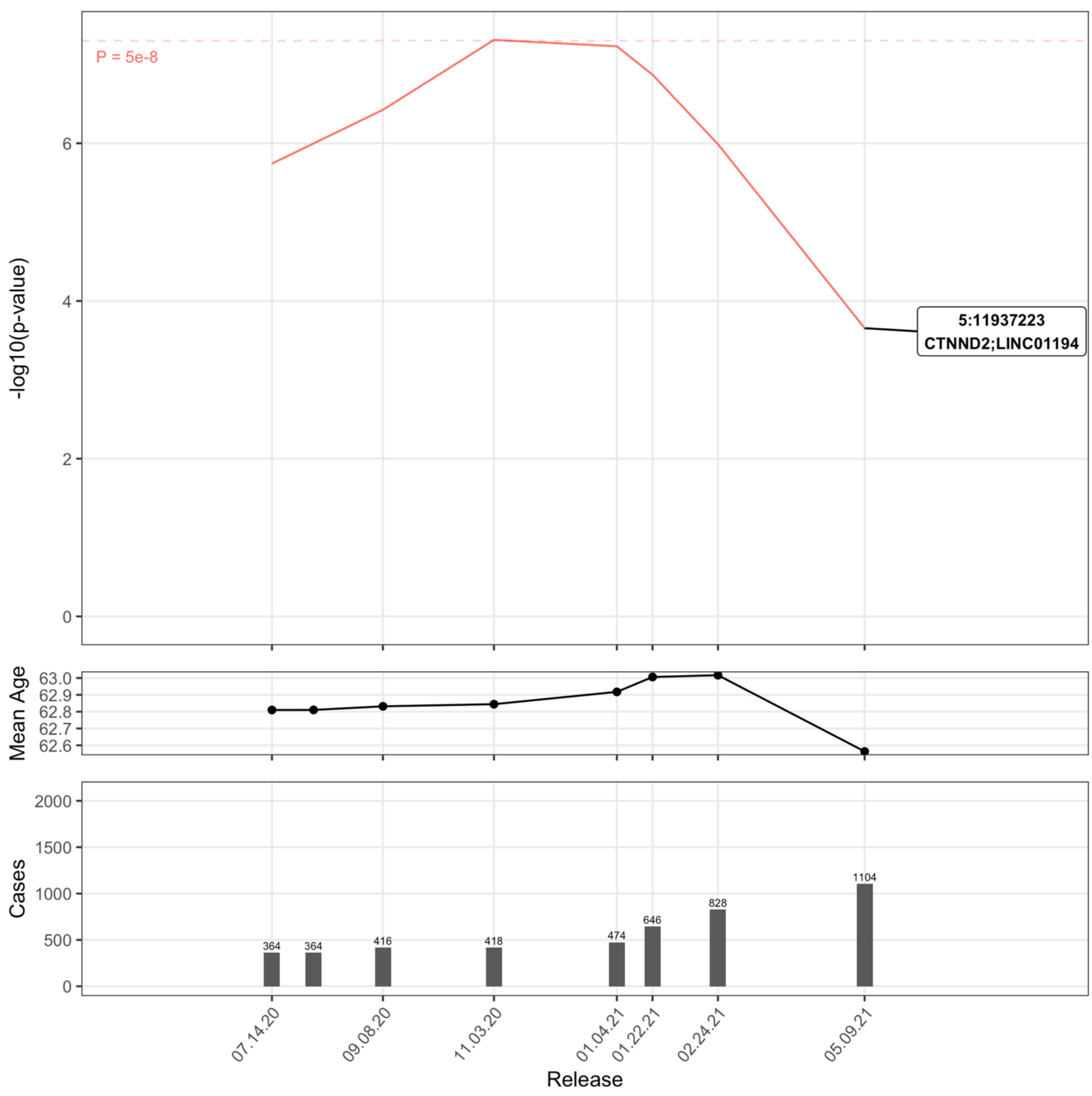

Supplemental Figure 45: Covid-19 Death in EUR:Pop

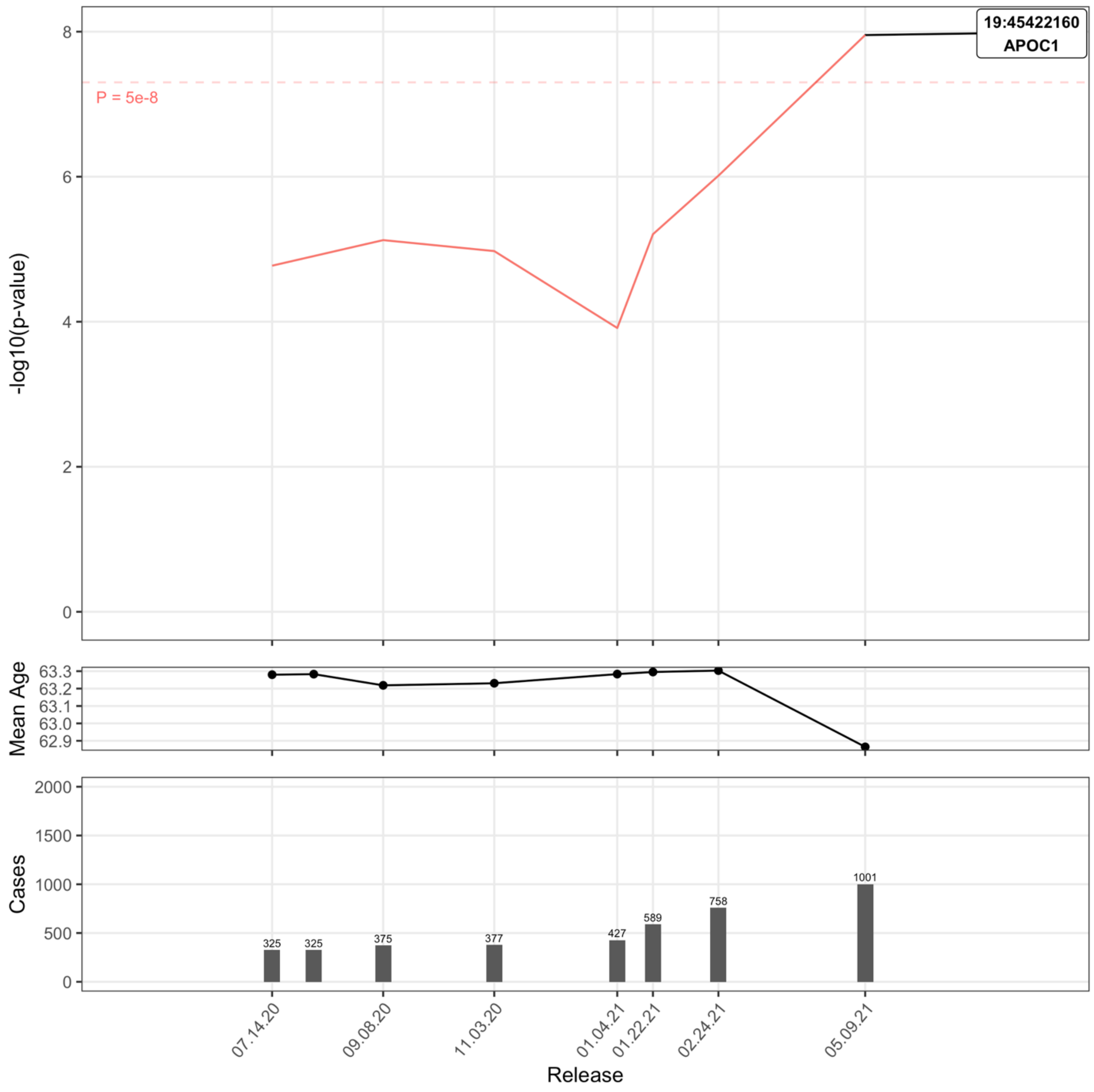

Supplemental Figure 46: Covid-19 Death in EUR:Positive

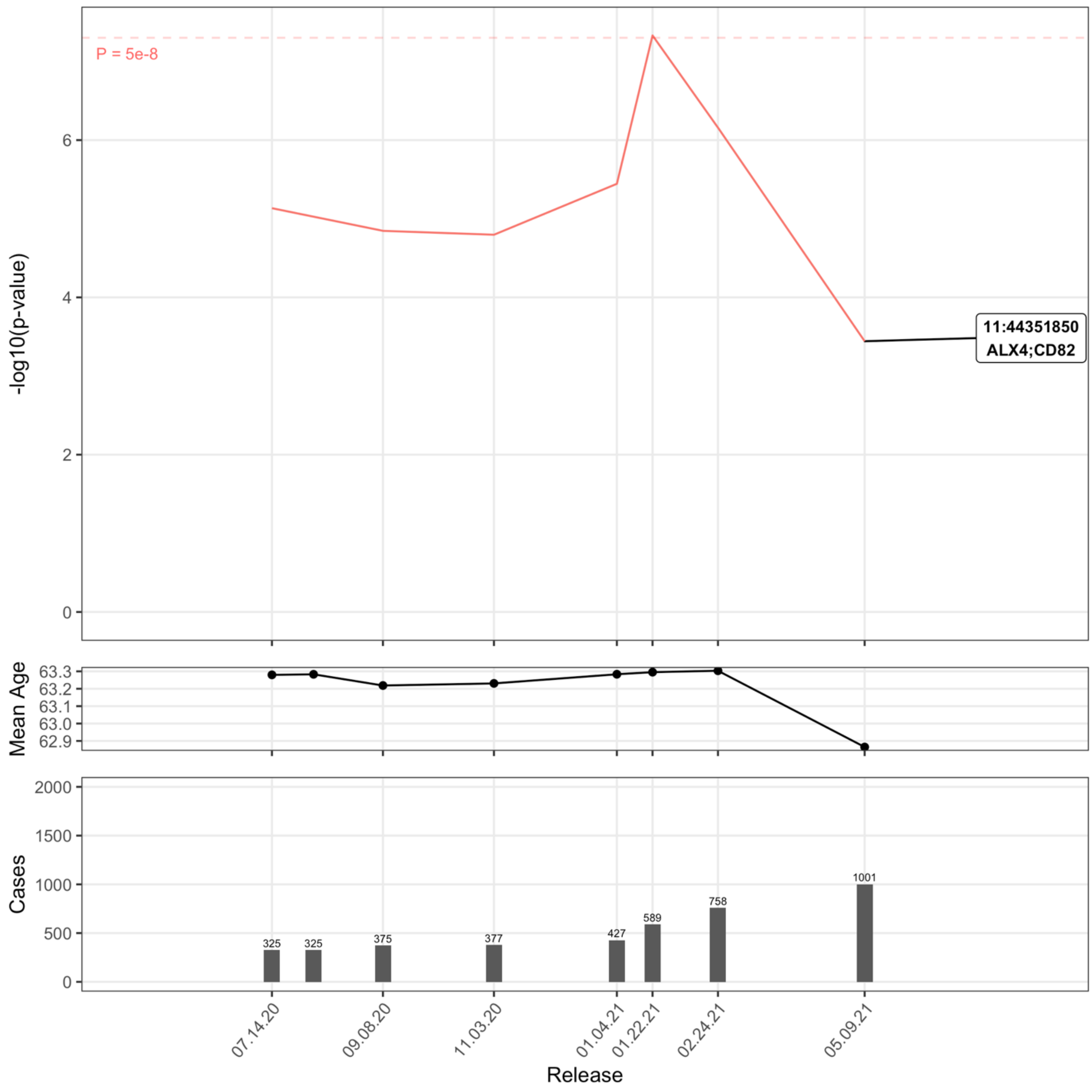

Supplemental Figure 47: Comparison of genome-wide significant signals in analyses using Population and Tested controls

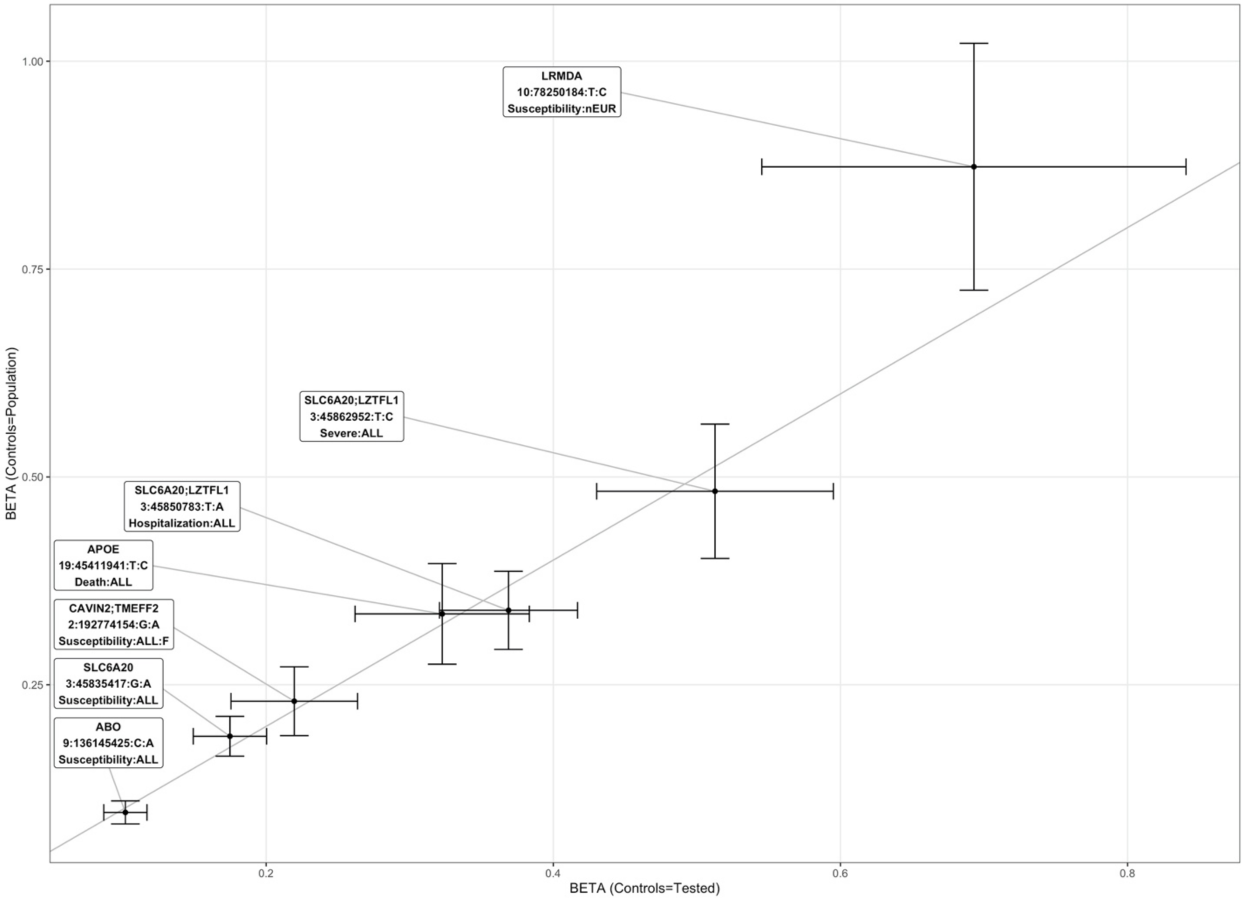


This figure presents the effect of each genome-wide significant signals and associated standard errors, when using the Population control set (on the Y-axis), or the Tested control set (on the X-axis).

Supplemental Figure 48 : Regional association plots of Covid-19 susceptibility and severity in Europeans (Population as controls), at the chr3p21.31 locus

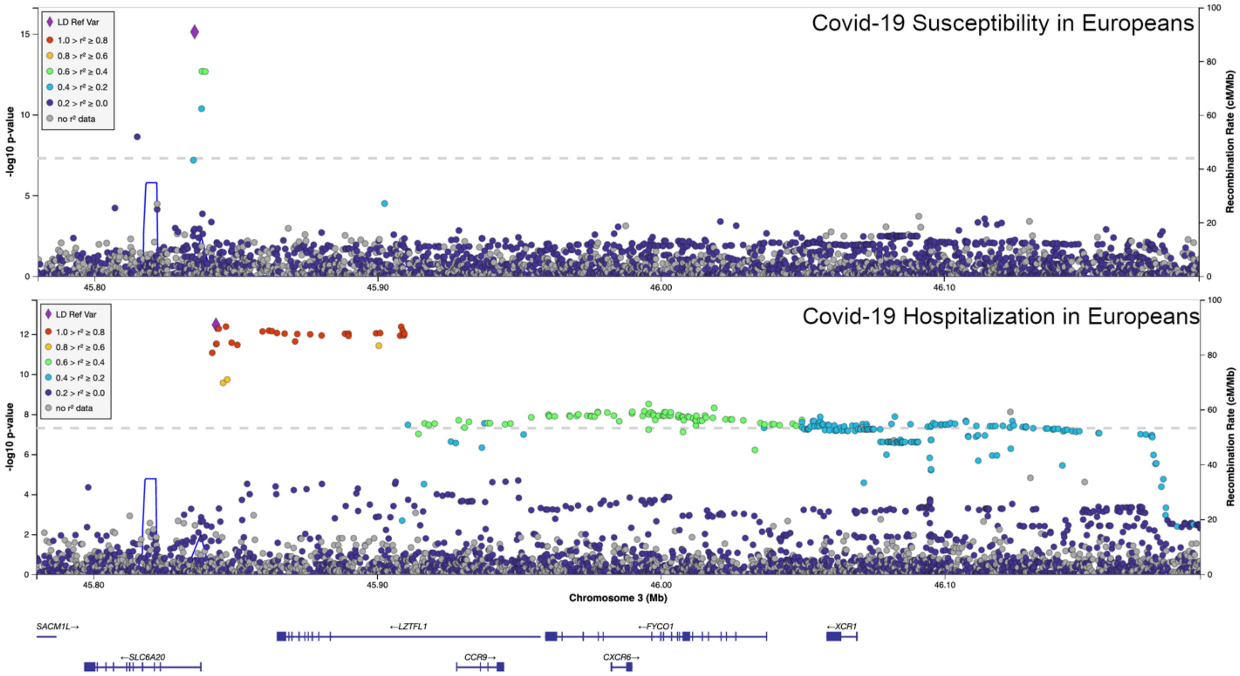


The top panel represent the Covid-19 susceptibility GWAS results in Europeans, at the chr3p21.31 locus, while the bottom panel represent the Covid-19 severity GWAS in Europeans at the same locus. The y-axis represents the -log10(P-value). The linkage disequilibrium between the lead SNP of both analyses (represented as a purple diamond) and each variant follows a color scheme presented on the left side of each panel.

Supplemental Figure 49: Haplotype analysis of ABO blood groups, including the variant associated with Covid-19 susceptibility (from the LDlink tool LDhap)

| | | | | Haplotypes and corresponding blood groups: | | | | | |
|---|---|---|---|---|---|---|---|---|---|
| | RS Number | Position (GRCh37) | Allele Frequencies | O1 | A1 | A2 | B | O2 | B |
| Variant tagging for O2 group : | rs41302905 | chr9:136131316 | C=0.971, T=0.029 | C | C | C | C | T | C |
| Variant tagging for B group : | rs8176743 | chr9:136131415 | C=0.915, T=0.085 | C | C | C | T | C | T |
| Variant tagging for A2 group : | rs1053878 | chr9:136131651 | G=0.901, A=0.099 | G | G | A | G | G | G |
| Variant tagging for O1 group : | rs8176719 | chr9:136132908 | -=0.605, C=0.395 | - | C | C | C | C | C |
| Variant tagging for A1 group : | rs2519093 | chr9:136141870 | C=0.815, T=0.185 | C | T | C | C | C | C |
| Variant associated with Covid susceptibility : | rs9411378 | chr9:136145425 | C=0.701, A=0.299 | C | A | A | C | C | A |
| | | | **Haplotype Count** | 603 | 183 | 94 | 68 | 29 | 17 |
| | | | **Haplotype Frequency** | 0.5994 | 0.1819 | 0.0934 | 0.0676 | 0.0288 | 0.0169 |

The 5 SNPs tagging for the 5 most common blood groups, as well as the variant associated with Covid-19 susceptibility were included in this analysis. This analysis relies on the 1000 genome dataset. The resulting haplotypes, and corresponding blood groups (tagged by their associated variants) observed in this population are indicated on the right. The bottom part indicates the frequency of each haplotype observed. All A1 and A2 haplotypes contain the Covid-19 variant. Out of the 85 B haplotypes present in this population, 17 (20%) also contain the Covid-19 variant.

Supplemental Figure 50: Evolution of the signals from Figure 1 (Susceptibility signals in EUR:Pop, from UKB) in Covid-19hgi C2 analyses

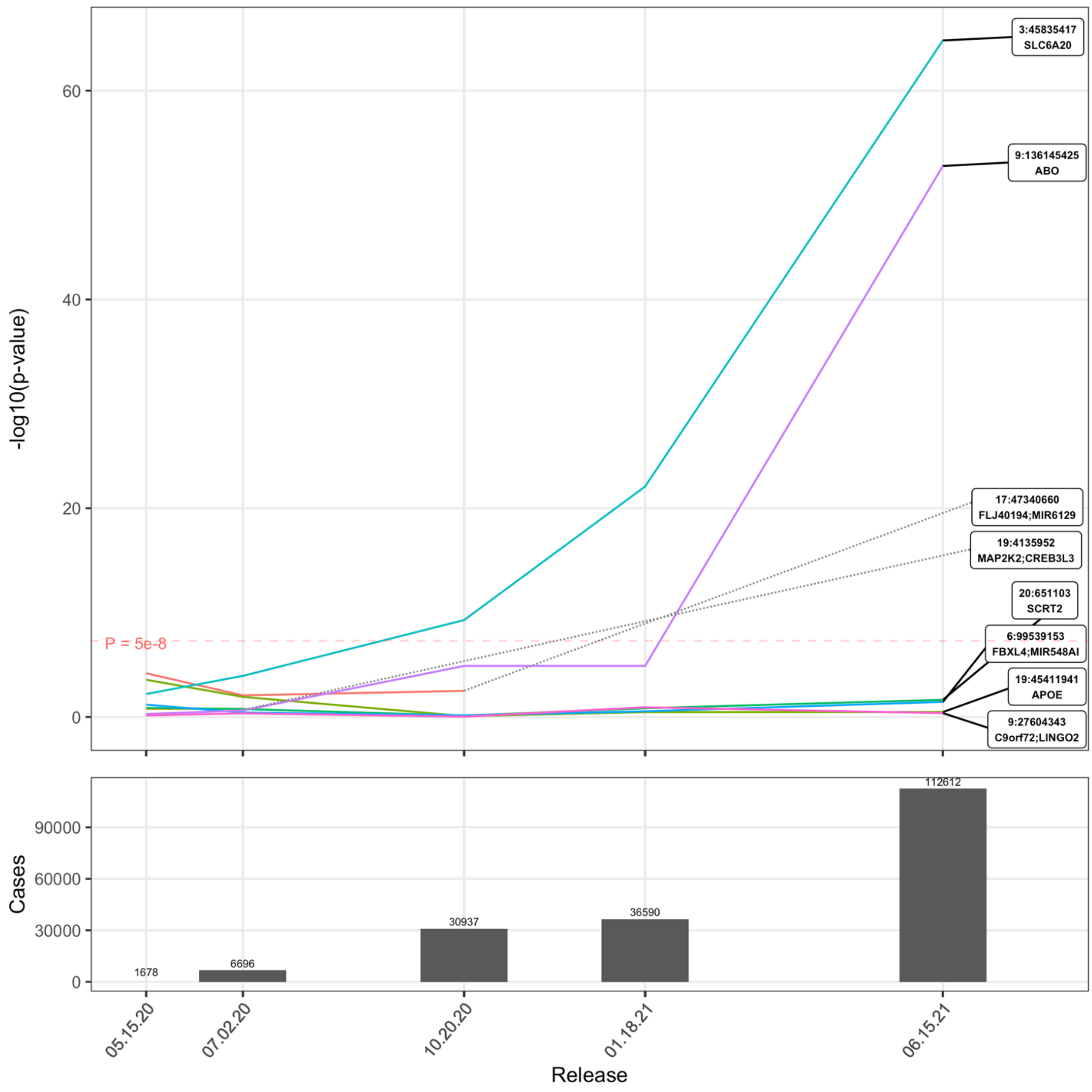


This figure represents the 8 signals from Figure 1 across the 5 C2 freezes from Covid-19hgi. 2 signals (19:4135952 and 17:47340660) were only available in the first 2 and first 3 freezes, respectively. The ABO signal was absent in freeze 5 (released on 01.18.21).
